# Inherited Susceptibility to Urinary Tract Infections from Kidney Papilla to Bladder

**DOI:** 10.64898/2026.07.07.26357417

**Authors:** Katherine Xu, Atlas Khan, Ning Shang, Wenjie Zeng, Chen Wang, Cecilia Berrouet, Tian Huai Shen, Padma Narayanan, Jenny Deng, Chiara DiPerna, Charlotte Williams, Sarah Cuacuas, Timothy R. Olsen, Jeffrey Arace, Aryan Ghotra, Tai Wei Guo, Wayne Monical, Oleg Borisov, Stefan Haug, Hongbo Liu, Eunji Ha, Andrew Yaeh, Run Banlengchit, Abraham Levitman, Bartlomiej Halibart, Diya Patel, Claire Chou, Shawn Simmons, Sanya Goswami, Kivanc Nesanir, Masashi Fujita, Iftikhar J. Kullo, Gail P. Jarvik, Wei-Qi Wei, QiPing Feng, Lan Jiang, C. Michael Stein, Chunhua Weng, George Hripcsak, Ali G. Gharavi, Katalin Susztak, Philip L. De Jager, Anna Köttgen, Jonathan Barasch, Peter A. Sims, Krzysztof Kiryluk

## Abstract

Urinary tract infections (UTIs) are traditionally viewed as environmentally driven, yet their inherited susceptibility remains largely unexplored. We conducted a cross-biobank genome-wide association study of recurrent UTIs in 1,860,836 individuals (213,869 cases and 1,646,967 controls). We identified 36 genetic susceptibility loci and performed tissue-based multi-omic mapping to prioritize candidate causal genes. UTI risk alleles preferentially modulated epithelial gene expression in kidney and bladder, converging on urinary epithelia structure and function. *PSCA*, encoding a secreted epithelial surface protein, emerged as the strongest candidate under genetic control; the gene product is constitutively secreted into the urine from kidney papilla and bladder epithelia, binds uropathogenic *E. coli*, and inhibits bacterial growth *in vitro*. Our findings define the polygenic architecture of UTIs and highlight the critical role of uroepithelial surface defenses, providing a new framework for host-directed, non-antibiotic interventions.

## INTRODUCTION

Urinary tract infections (UTIs) are among the most common bacterial infections worldwide, affecting more than 400 million individuals annually (*1*). Nearly half of all women will experience at least one UTI in their lifetime, and up to 30% will develop recurrent disease (*2–5*); infants, pregnant individuals, the elderly, and patients with diabetes, neurogenic bladder, urinary catheters, or anatomical abnormalities face additional risk (*6*). UTI has traditionally been treated as a largely environmental or acquired condition, a consequence of anatomy, hygiene, sexual activity, and bacterial exposure, rather than inherited biology. Yet even among individuals who share these same exposures and risk factors, susceptibility varies enormously (*7*). Some remain infection-free for life, while others suffer recurrent episodes for years. This variability makes the traditional view increasingly untenable, implicating a genetic component to UTI risk.

It is of growing clinical importance to define host pathways that govern inherited susceptibility to infection. While most infections are treated successfully with antibiotics, increasing rates of antibiotic resistance, particularly among uropathogenic *Escherichia coli* (UPEC; ∼80% of outpatient cases (*8*)), threaten the effectiveness of conventional therapies and highlight the need for alternative prevention and treatment strategies. Identification of inherent risk factors may contribute to these new approaches.

Ascending infection typically begins when intestinal bacteria migrate across the perineum to colonize the urinary tract. The infection must overcome an integrated and multilayered mucosal defense network that extends beyond simple mechanical clearance. Urine flow, acidic pH, and secreted glycoproteins limit bacterial attachment and colonization, while specialized epithelial cells actively participate in antimicrobial defense. Uromodulin, the most abundant urinary protein, binds type 1 pili of UPEC and prevents bacterial adhesion to the urothelium (*9*). Intercalated cells of the kidney collecting duct acidify urine and secrete antimicrobial proteins (*10–12*). In parallel, urothelial and renal epithelial cells sense invading pathogens through pattern recognition receptors and initiate inflammatory responses that recruit innate and adaptive immune cells (*12*). Much of what is known about these defenses relies on mouse models of experimentally induced infection, typically by inoculating the bladder with UPEC. These studies, however, best characterize tissue responses to infection rather than the underlying physiologic risk of infection.

Despite these defenses, a heritable component to UTI risk is already evident in several rare genetic conditions. Congenital urinary tract anomalies such as primary vesicoureteral reflux, seen in one in three children with UTIs, exhibit familial aggregation (*13*). Inherited disorders of urine chemistry, such as some forms of distal renal tubular acidosis (*14*), also increase risk of recurrent UTIs (*15*). Indeed, prior studies have shown that UTIs cluster in families, and that first-degree relatives of women with recurrent UTIs are at increased risk (*16*). However, these are individually uncommon conditions driven by rare variants. Whether common genetic variation confers risk of recurrent urinary tract infections independently of environmental or anatomical factors has not been systematically investigated at the population level. We reasoned that individuals with recurrent infections but no established risk factors, such as catheterization or anatomical abnormalities, would be enriched for inherited susceptibility.

Here, we performed a genome-wide association meta-analysis of 1,860,836 individuals across multiple biobanks to define the genetic architecture of susceptibility to UTIs. We identified 36 genome-wide significant loci and integrated genetic associations with regulatory annotations, urinary tract single-cell atlases, and expression and protein quantitative trait loci to prioritize candidate causal genes and pathways. Our findings reveal a convergent genetic landscape in which epithelial barrier function, innate immune regulation, developmental patterning of the urinary tract, and epithelial nutritional immunity collectively shape susceptibility to UTI. Rather than being induced in response to infection, these inherited programs appear to be constitutively active, forming a physiologic defense that is already in place before bacteria arrive. These results establish epithelial and immune programs of host defense as major, hereditable determinants of infection risk and provide new avenues for treatment and prevention of UTIs in the era of rising antibiotic resistance.

## RESULTS

### Study design

We developed an electronic UTI phenotyping algorithm using diagnostic codes and available urine studies to define UTI cases and non-UTI controls in electronic health record (EHR)-linked biobanks including Electronic Medical Records and Genomics (eMERGE) (*17*), All-of-Us (AoU) (*18*), UK Biobank (UKBB) (*19*), and BioVU (*20*) (fig. S1, Methods). Combining these cohorts with GWAS summary statistics from the Million Veteran Program (*21*), FinnGen (*22*), and 23andMe (*23*), we performed the largest multi-ancestry GWAS meta-analysis for UTIs, comprising a total of 1,860,836 individuals (213,869 cases and 1,646,967 controls). Across all biobanks, UTI cases were defined as individuals with ≥2 ICD-9/10 diagnostic codes for primary UTI/cystitis or pyelonephritis, regardless of bacterial species. Controls were defined by the absence of any UTI-related ICD codes. The final counts of cases and controls by cohort and their characteristics are summarized in table S1.

### Discovery of genetic susceptibility loci

Our primary GWAS meta-analysis defined 28 independently genome-wide significant loci (*P* < 5×10^−8^, HLA locus and 27 non-HLA loci; Fig. 1A, Table 1, data S1), including the previously reported *UMOD* locus on chr.16p12.3 (rs12917707, *P* = 6.8×10^−17^) and a *PSCA* locus on chr.8q24, (rs2920286, *P* = 4.5×10^−12^) (*23*). We also defined 59 independent suggestive loci at *P*<1×10^−6^ (data S2), some of which reached genome-wide significance in secondary analyses stratified by ancestry and sex (figs. S2 and S3, data S3 and S4). For example, suggestive loci, *CASP7* on chr.10q25 and *LINC00899* on chr.22q13 were genome-wide significant in the European ancestry (*CASP7*: rs471447, *P*=3.2×10^−8^; *LINC00899*: rs7285579, *P* = 3.7×10^−9^), while the *BMP7* locus on chr.20q13.31 was genome-wide significant in the male-specific analysis (rs6099264, *P*=1.4×10^−10^). In total, the European- and male-only analyses identified 30 and 7 genome-wide significant loci, respectively, including 9 novel associations not captured in the overall meta-analysis (figs. S2 and S3, data S3 and S4). We did not detect any new genome-wide significant associations in other ancestral groups or female-only analyses. Analysis of sex-stratified effect sizes for genome-wide significant loci by Cochran’s heterogeneity statistic (data S5) highlighted a few notable differences between males and females, with *CLPTM1L* (I^2^=0.83, Cochran’s P=0.02, Fig. 1B) and *FGFR2* (I^2^=0.76, Cochran’s P=0.04, fig. S3), emerging as male-driven loci and *PSCA* (I^2^=0.62, Cochran’s P=0.10, Fig. 1B) trending as a predominantly female-driven locus.

**Fig. 1.**
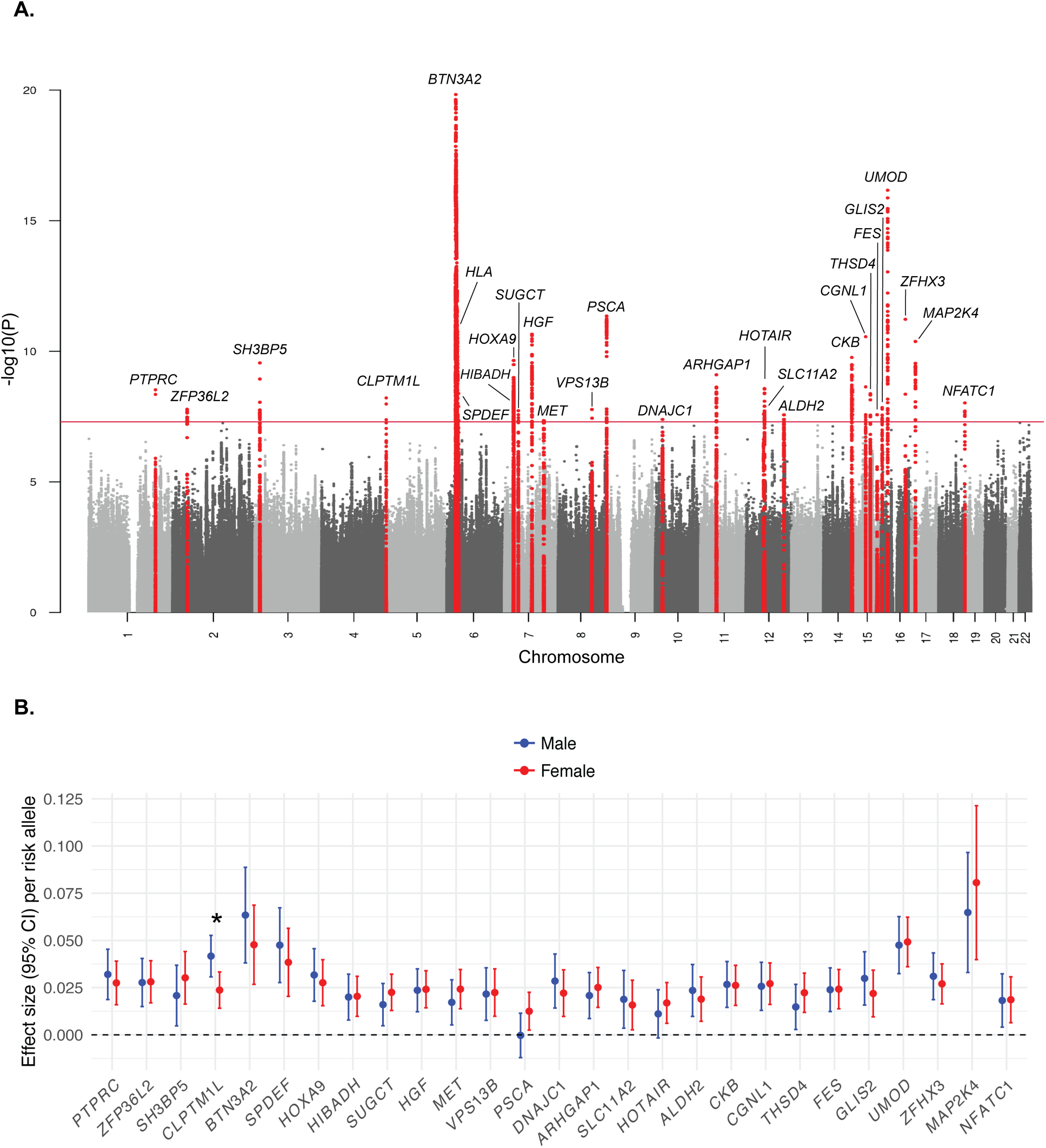
GWAS meta-analysis reveals 28 independent genome-wide significant loci (27 non-HLA and 1 HLA locus) associated with UTI. **(A)** Manhattan plot of GWAS meta-analysis results displaying -log10(P) values across all chromosomes. The red horizontal line denotes the genome-wide significance threshold (P<5×10⁻⁸). Genome-wide significant loci are highlighted in red, with prioritized candidate genes annotated above each locus. **(B)** Sex-stratified effect sizes (95% CI) per risk allele for genome-wide significant loci in males (blue) and females (red). Analysis of sex-stratified effect sizes using Cochran’s heterogeneity test statistic highlighted notable differences between males and females at select loci, with statistically significant heterogeneity (P<0.05) denoted by an asterisk (*).

**Table 1:**
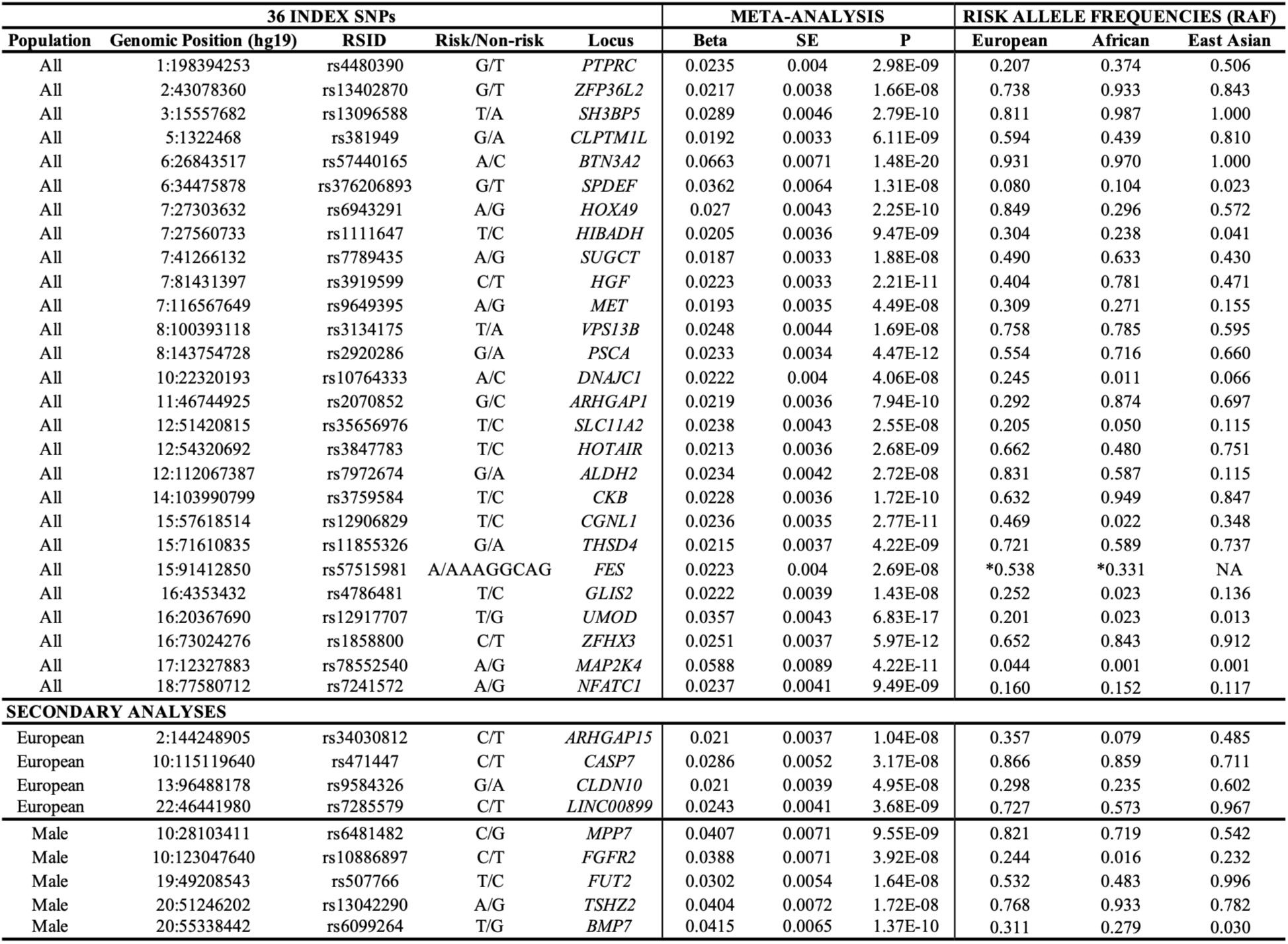
Thirty-six genome-wide significant index SNPs (hg19) associated with UTI across both primary and secondary GWAS meta-analyses. Risk allele frequencies (RAF) were obtained from the 1000 Genomes Project Phase 3 (*24*) reference panel. *Frequency obtained from NCBI Allele Frequency Aggregator (ALFA) (*96*).

In our primary and secondary analyses, we defined independent genome-wide significant associations at individual loci using the following criteria: lead SNP with *P* < 5×10^−8^, pairwise linkage disequilibrium (LD) r^2^ < 0.01 (1000G Phase 3 reference) (*24*) and a distance of more than ±250 kb from another lead SNP. Applying this approach across both analyses, with duplicate loci removed, we curated a final list of 36 non-HLA index SNPs (27 loci in primary analysis and 9 loci in secondary analysis) for downstream analyses (Table 1, fig. S4).

### Genetic annotations of susceptibility loci

We annotated all variants in strong LD with each of the 36 index SNPs based on external reference (r^2^ ≥ 0.7 in 1000G) using ANNOVAR (*25*) and VEP (*26*). Most SNPs mapped to intronic or intergenic regions, but proxy SNPs at two loci intersected coding sequence of *FUT2* and *LRP4* genes (data S6), including a premature stop variant (p.Trp154Ter) in *FUT2* in high LD with the index SNP at this locus (r^2^=0.85). We also interrogated all 36 lead SNPs and their proxies with PromoterAI (*27*) for variants predicted to alter promoter activity and identified candidate variants in promoter sequences of *PSCA* and *DDB2* (data S7). Next, we intersected our loci with *cis*-regulatory elements (cCREs) catalogued by the ENCODE4 project (*28*, *29*), and identified overlaps at 33 loci. This comprised of 174 unique locus-cCRE intersections, including 117 (67%) intersections of distal enhancer-like signatures, 21 (12%) proximal enhancer-like signatures, 12 (7%) chromatin accessibility regions without enhancer or promoter signatures, 8 (5%) transcription factor-associated sites, 5 (3%) promoter-like signatures, 5 (3%) CTCF-associated chromatin accessibility elements, 4 (2%) combined chromatin accessibility-TF signatures, and 2 (1%) H3K4me3-associated chromatin accessibility elements (data S8). While most loci intersected only one or two candidate regulatory regions (fig. S5), several loci intersected larger clusters of multiple regulatory elements, such as *CLPTM1L* (N=30 distinct elements), *CKB* (N=19), *PSCA* (N=18), *ALDH2* (N=14), and *SLC11A2* (N=12). Collectively, these findings suggest that UTI risk loci exhibit their effects mostly through disrupting gene regulatory elements, particularly distal enhancers. Because of the observed enrichment of distal enhancers across the risk loci, we next applied Activity-by-Contact (ABC) enhancer-gene mapping, a method that links distal enhancers to their target genes (*30*). Using the Variant-to-Gene (V2G) pipeline (*31*), we intersected our loci with the ABC maps for 131 human ENCODE samples spanning 74 primary cell types, tissues, and cell lines. We identified 169 unique variant-enhancer-gene mappings with ABC score ≥ 0.02 involving 49 enhancers across 17 loci (data S9), including *CLPTM1L* (rs381949; ABC score=0.45), *DDB2* and *ARHGAP1* (rs2070852; ABC scores=0.41 and 0.38), and *FURIN* (rs57515981; ABC score = 0.34) among the highest scoring genes.

Because kidney and lower urinary tract cell types were not included in the ABC maps, we next refined our analysis using single-nucleus RNA/ATAC-seq multiome data from 25 healthy participants of the Kidney Precision Medicine Project (KPMP) (*32*, *33*) to generate novel kidney cell type specific enhancer-to-gene maps using the scE2G framework (*34*). In total, we detected 14 GWAS loci intersecting with predicted kidney enhancers that had E2G score ≥ 0.177 for genes expressed across 18 kidney cell types (fig. S6). This included 259 unique enhancers and 348 enhancer-gene pairs, prioritizing 39 target genes across 14 loci (fig. S7, data S10). The greatest regulatory activity was observed at *CLPTM1L* (rs381949; n=51), *UMOD* (rs12917707; n=38), and *ALDH2* (rs7972674; n=26). The UTI variant-enhancer-gene links were distributed broadly across tubular, stromal, and immune cell populations, although partitioned heritability enrichment of UTI risk loci across kidney scE2G maps via linkage disequilibrium score regression partitioning demonstrated enhanced enrichment within principal cells (20-fold, P=0.04) and vascular smooth muscle cells/pericytes (41-fold, P=0.04, data S11). Together, our kidney V2E2G mapping demonstrates widespread kidney cell-type-specific regulatory activity of the UTI risk loci, with most prominent enrichment in the collecting duct and vasculature.

We additionally intersected our 36 GWAS loci with an independent dataset of previously published human kidney Open4Gene (O4G) regulatory maps based on snATAC-seq of 30 human kidneys (*35*). At 7 loci, we observed 26 significant variant-peak-gene links involving 10 kidney genes (data S12). Of note, the scE2G and O4G regulatory mapping provided convergent evidence for 48 genes across 15 loci, including for *UMOD*, *PSCA*, *CLPTM1L*, *FES*, *SLC11A2*, *ALDH2*, *CKB*, *GLIS2*, and *LINC00899*.

### Gene expression in the kidney and urinary tract

We next examined gene expression of the GWAS prioritized candidate genes across relevant tissues. We used the Kidney Precision Medicine Project (KPMP) single nucleus RNA-sequencing (snRNA-seq) kidney atlas version V19 (*36*) and single cell atlases of the human ureter (*37*), bladder (*38*), urine cells (*39*), and peripheral blood mononuclear cells (PBMCs) (*40*). Because current datasets do not provide high resolution maps of the kidney papilla, the point of bacterial entry into the kidney, we micro-dissected papilla tissue from healthy mouse kidneys, performed snRNA-seq (Fig. 2A), and integrated these data with the existing atlases. Within the papilla, we defined specific papillary cell types, including intermedullary collecting duct principal cells (*Aqp2*+), intercalated cells (*Atp6v1b1*+), and cells of the ascending (*Clcnka*+) and descending (*Aqp1*+) limbs of the loop of Henle (fig. S8, data S13). Using these data, we mapped our candidate GWAS genes onto the cell types along the entire urinary tract (data S14-20), revealing cell-type specificity for many of the genes. Within the kidney, *UMOD* expression was prominently localized to the thick ascending limb (TAL) of the loop of Henle, while *SLC11A2* showed enrichment in the collecting duct and urothelial cells (Fig. 2B). Lower in the urinary tract, *PSCA* localized to kidney papillary ascending thin limbs (ATL), as well as umbrella cells of the bladder and ureter, with high expression also detected in the cells shed in urine (Fig. 2B). *MPP7* showed relatively specific expression in papillary IMCD-CD cells and urothelial layers of the ureter and bladder. In contrast, *PTPRC*, *ARHGAP15*, and *BTN3A2* were broadly expressed across myeloid and lymphoid populations (Fig. 2B), suggesting immune-mediated effects of these genes. Together, these analyses implicate kidney papilla cells, urothelial cells, and immune cells across the urinary tract as potential mediators of genetic susceptibility to UTI.

**Fig. 2.**
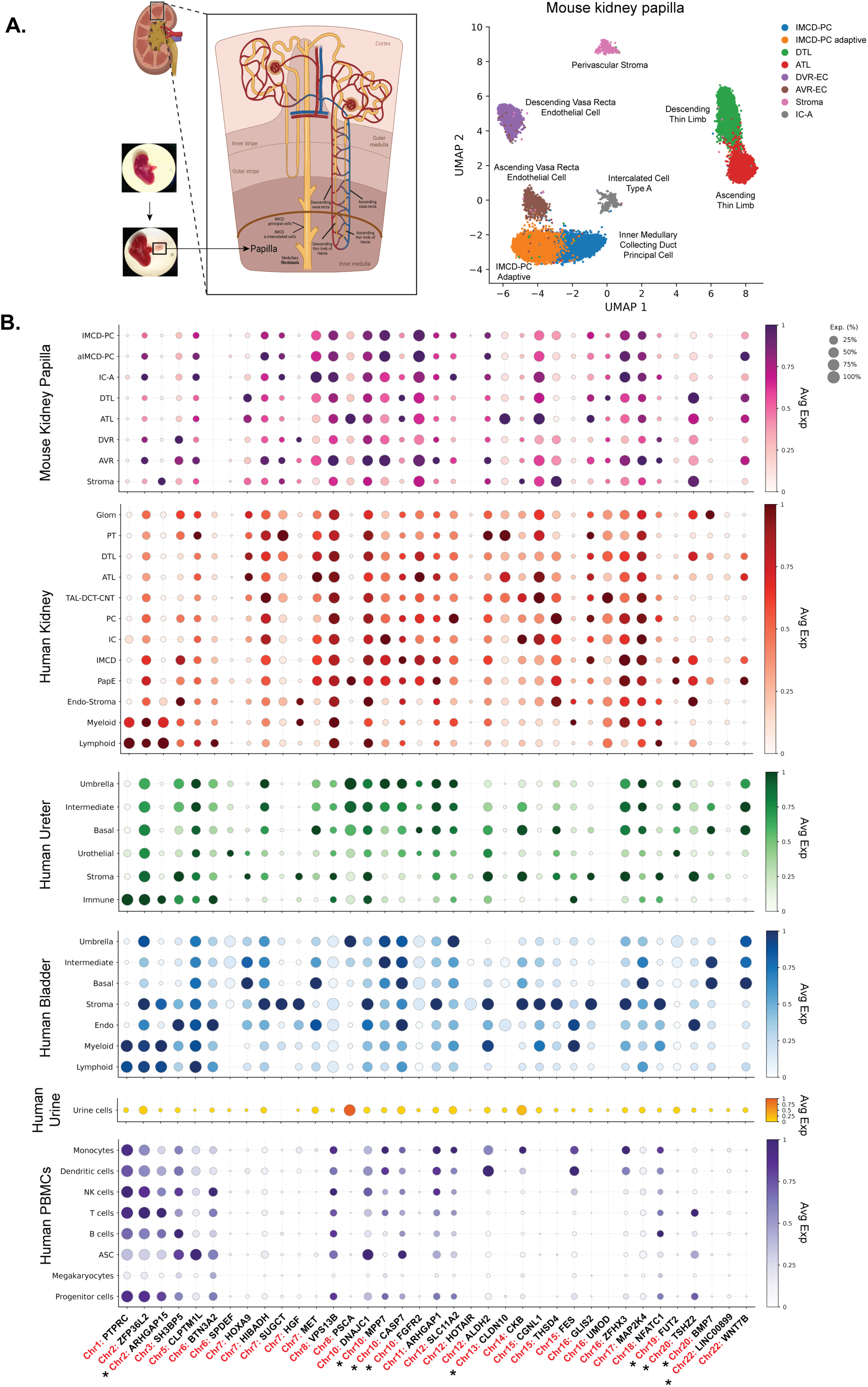
Mapping genome-wide significant GWAS signals to genes expressed in single-cell transcriptomic profiles of urinary tract tissues. **(A)** Schematic of the kidney (created with BioRender.com) highlighting the papilla region with a representative image of a microdissected mouse kidney papilla (left), with a corresponding UMAP of single-cell transcriptomic data (right) showing distinct cell type clusters: IMCD-PC (Inner Medullary Collecting Duct Principal Cell), IMCD-PC Adaptive, DTL (Descending Thin Limb), ATL (Ascending Thin Limb), DVR-EC (Descending Vasa Recta Endothelial Cell), AVR-EC (Ascending Vasa Recta Endothelial Cell), Stroma, and IC-A (Intercalated Cell Type A). **(B)** Dot plots displaying row normalized average expression (Avg Exp) of prioritized candidate genes at each of the 36 UTI GWAS loci across cell types from four urinary tract tissue datasets: mouse kidney papilla, human kidney, human ureter, and human bladder, as well as from human urinary cell and peripheral blood mononuclear cell (PBMCs) datasets. For human bladder, dot size represents relative expression. For all other tissues, dot size represents the percentage of cells expressing each gene (Exp. (%): 25%, 50%, 75%, 100%). See data S14-20 for full list of genes within the GWAS loci and their expressions across the urinary tract. Asterisks (*) represent prioritized genes from GWAS loci that were identified from secondary (European- or Male-specific) meta-analyses.

### Genetic co-localizations with urinary tract transcriptome and urinary proteome

To identify associations between GWAS variants and gene expression or splicing usage in the kidney and urinary tract, we first queried publicly available maps of urinary tract and immune cell expression and splicing quantitative trait loci (e/sQTLs) (*41–43*) (data S21). We then tested the 36 GWAS loci for evidence of colocalization (*44*) with tissue-specific e/sQTLs across bladder (*41*), kidney cortex(*41*), kidney tubules (*42*, *43*), prostate (*41*), blood (*45*), and immune cells (*46*). Nineteen of 36 (53%) loci demonstrated strong evidence of colocalization (PP4 ≥ 0.80) in at least one tissue or QTL modality (fig. S9, data S22). Across all eQTL analyses, the strongest and most consistent evidence was observed for *PSCA*, which colocalized across kidney, bladder, and prostate (PP4 0.81-0.96), with the risk allele consistently associated with lower expression of *PSCA* across all datasets. *ARHGAP1* showed similarly concordant colocalization across kidney, prostate, and immune/blood cells (PP4 0.80-0.98), as well as *HOTAIR* (kidney and prostate, PP4 0.87-0.91) with risk alleles at both loci associated with lower gene expression, suggesting their protective role in urinary antimicrobial defenses. In contrast, the colocalized risk alleles at the *FES* (prostate and monocytes, PP4 0.89-0.98) and *FUT2* (prostate, PP4=0.91) loci were associated with increased expression of these genes, suggesting potential detrimental effects of the gene products. Colocalizations of sQTLs were also observed at several of the same loci, including *UMOD* and *PSCA* (kidney cortex) and *FES* (prostate and immune/blood), suggesting that these loci may simultaneously affect splicing and gene expression levels.

We next interrogated our loci against the first map of urinary protein QTLs (pQTLs) based on 1,246 participants of the German Chronic Kidney Disease study (fig. S9, data S22) (*47*). We observed strong co-localization of the *PSCA* locus with urinary PSCA protein levels (PP4=0.96). Consistent with the eQTL effect, the UTI risk allele strongly associated with reduced urinary PSCA levels (β=-1.1, P<2.2×10^−308^), and this pQTL represented the single strongest urinary cis-pQTL across the entire urinary proteome (*47*). Lastly, we performed plasma proteome-wide association study for UTI based on our GWAS summary statistics and plasma Olink 3K data for the UK Biobank (fig. S10, data S23). The proteome-wide significant signals provided further support for *UMOD* (P=1.9×10^−14^), *BTN3A2* (P=1×10^−13^), and *FES* (P=2.8×10^−6^).

In summary, integration of transcriptomic and proteogenomic datasets revealed that some UTI risk alleles were associated with reduced expression of epithelial surface genes, such as *PSCA, ARHGAP1*, and *HOTAIR,* while others with increased expression of immune and inflammatory regulators (*FES, FUT2, BTN3A2*).

### Prioritization of candidate genes across 36 UTI risk loci

To systematically prioritize candidate causal genes within the 36 GWAS loci defined by all proxy variants in LD (r^2^≥0.7) and within ±500kb of the lead SNP, we scored our various annotation modalities for convergence by assigning one point for each distinct prioritization method. The **genomic annotation criteria** included: (1) gene most proximal to the lead SNP (nearest gene), (2) gene harboring missense or loss-of-function proxy variants, (3) genes with promoter regulatory proxy variants (PromoterAI score > 0.1), (4) genes with regulatory proxy variants linked through ABC maps (*30*) (ABC score > 0.2), (5) genes with proxy variants intersecting kidney-specific scE2G links (*34*) (scE2G score > 0.177), and (6) genes that were mapped using Open4Gene (O4G) (*35*) kidney-specific enhancer-gene maps at FDR<0.05. The **tissue-specific annotation criteria** included genes within ±500kb of index SNP with cell type specific gene expression enriched in (7) kidney cells (*36*), including mouse papillary cells, (8) lower urinary tract cell types, including ureter (*37*) and bladder (*38*), (9) cells shed in urine (*39*), and (10) urinary tract resident and circulating immune cells. The **genetic evidence criteria** included (11–13) genes within ±500kb of index SNP that have urinary tract or immune cell e/sQTLs based on lookups in publicly available datasets; genes with eQTL co-localizations (PP4>0.8) in (14) kidney tissue (GTEx (*41*), Neptune (*42*), or Susztak (*43*)), (15) lower urinary tract (GTEx bladder and prostate (*41*)), and (16) immune cells (*46*) or whole blood (*45*); genes with sQTL colocalizations (PP4>0.8) in (17) the kidney tissue, (18) lower urinary tract, or (19) whole blood/immune cells (*41*); (20) genes with urine pQTL co-localizations (PP4>0.8); and (21) genes associated with UTI risk by PWAS (*48*) (FDR>0.05). The **functional criteria** included: (22) genes differentially expressed in uroepithelial cells in mouse models of UTI (data S24), (23) genes with mouse knockout phenotypes involving either the upper or lower urinary tract or innate immunity based on Mouse Genome Informatics (*49*) database (data S25), (24) genes functionally related to UTI risk prioritized by manual PubMed, Google Scholar, and Harmonizome (*50*) review (data S26), and (25) top genes prioritized per locus by the majority vote of three large language models (LLMs): ChatGPT, Claude, and Gemini (data S27). Notably, as LLMs function in part as literature search tools, ChatGPT, Claude, and Gemini, showed high concordance with manual literature review (94.4%, 83.3%, and 88.9% respectively) but markedly lower concordance with the final prioritized gene at each locus (61.1%, 50.0%, and 55.6% respectively), suggesting LLMs alone are insufficient without integration of genetic and tissue-specific expression evidence.

Our integrative scoring framework identified a single top scoring biological candidate gene per each locus, each supported by convergent regulatory, genetic, and functional evidence (Fig. 3, data S28). More than half of the prioritized candidate genes (20 of 36) were also the most proximal genes to the lead SNP (Fig. 3). All 36 prioritized candidate genes were supported by at least six independent types of evidence. The genes with highest scores across all loci included *PSCA*, *FES*, *UMOD, CKB, ALDH2*, *ARHGAP1*, *ZFHX3*, *SLC11A2*, and *FUT2*, each accumulating strong evidence from multiple annotation methods. While *UMOD* served as a positive control, given its known role in host-urinary pathogen defense, *PSCA* was the single gene with the highest score across all 229 analyzed genes within 36 loci, with a strong support by urinary epithelial expression, V2E2G maps, eQTL and pQTL colocalizations, and functional signatures. The prioritized genes were involved in epithelial mucosal defense (*PSCA, FUT2, UMOD, CLPTM1L*), inflammation and programmed cell death (*HOTAIR, CASP7, STK3*), immune cell regulation (*PTPRC, BTN3A2, ARHGAP1, ARHGAP15*), and urinary tract developmental and repair programs (*FGFR2, BMP7, MPP7*, *WNT7B*, and *HOX* family genes), demonstrating mechanistic convergence at several distinct susceptibility pathways.

**Fig. 3.**
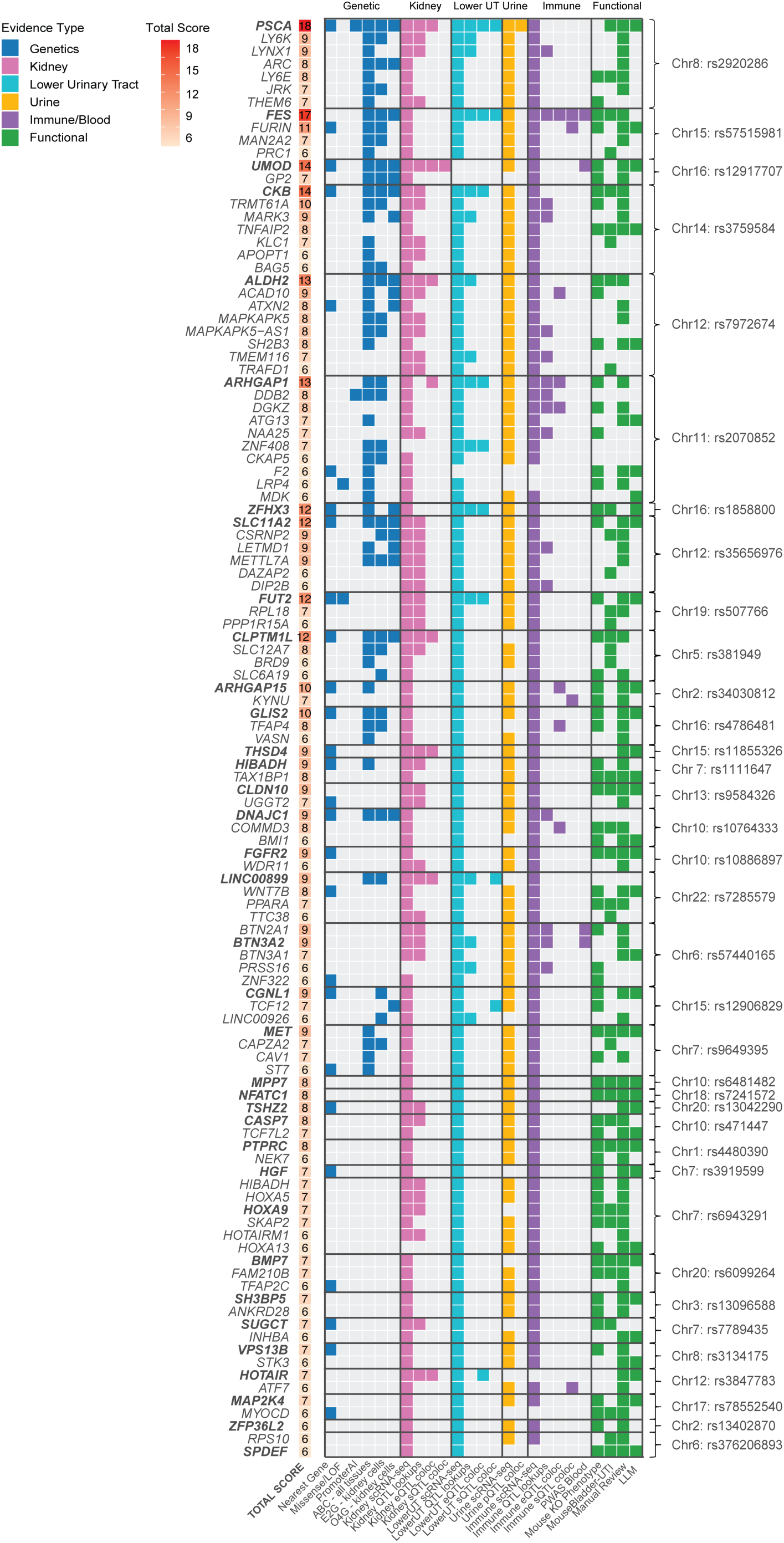
Prioritization of biological candidate genes across 36 UTI risk loci. Framework of multi-evidence gene prioritization scores for candidate genes across 36 UTI GWAS loci. Each row represents a candidate gene within a given locus and each column represents one of 25 binary evidence features (1 point per feature), grouped into genomic, kidney, lower urinary tract (which includes ureter, bladder, and prostate), urine, immune/blood, and functional evidence. The total score for each gene is tabulated and represented by a color gradient. Bolded gene names denote the top-prioritized candidate gene per locus, determined by the highest cumulative score, with ties broken by proximity to the lead GWAS variant. Genes are ordered by total score within each locus. See data S28 for a full list of genes within the GWAS loci and their prioritizations across 25 features.

### Shared susceptibility with other traits and pleiotropic associations of UTI loci

To interrogate shared susceptibility between UTI and other diseases, we examined genome-wide genetic correlations (*rg*) between UTI and 70 complex traits previously studied by GWAS and spanning infections, cardiometabolic disease, autoimmune disease, and cancer (Fig. 4A, data S29). UTI showed broad, predominantly positive genetic correlation with disease across all four domains, most strongly with other infections, such as yeast infections (*rg*=0.45, P=9.7×10^−13^) and pneumonia (*rg*=0.41, P=6.1×10^−9^), cardiometabolic disease, including venous thromboembolism (*rg*=0.37, P=8.6×10^−24^), essential hypertension (*rg*=0.32, P=1.8×10^−21^), coronary artery disease (*rg*=0.30, P=6.9×10^−15^), and type 2 diabetes (*rg*=0.26, P=8.7×10^−10^), autoimmune diseases, such as IgA nephropathy (*rg*=0.30, P=6.4×10^−7^), as well as rheumatoid arthritis and inflammatory bowel disease (*rg*=0.12-0.13, both P<5×10^−5^), and kidney cancer (*rg*=0.28, P=1.8×10⁻¹¹), the strongest correlation among cancers. These associations were highly consistent when we repeated the analysis excluding the HLA region (fig. S11, data S29), indicating that the shared genetic architecture between UTI and these other diseases is largely independent of the HLA locus and instead reflects a broader susceptibility to infection, inflammation, cardiometabolic and malignant disease.

**Fig. 4.**
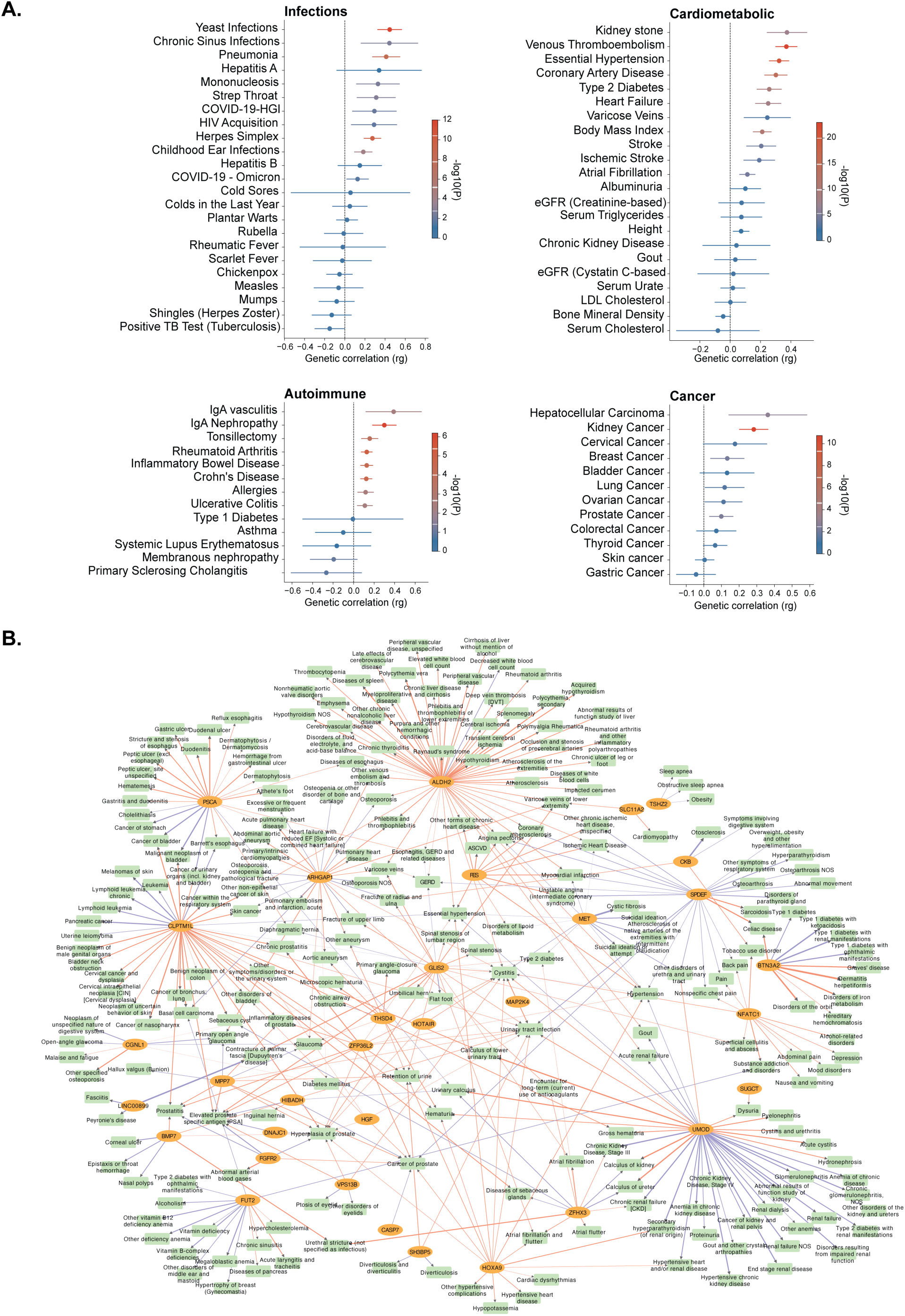
Genetic Correlation and Meta-PheWAS network of UTI GWAS meta-analysis with other GWAS traits. (A) Genetic correlations between UTI and complex traits across four disease domains. Forest plots show genetic correlations (*rg*) between UTI and (A) infections, (B) cardiovascular, renal, and metabolic traits, (C) autoimmune and inflammatory traits, and (D) malignancies, estimated by bivariate LD score regression (LDSC). Points represent rg point estimates; horizontal lines represent 95% confidence intervals. The dashed vertical line marks rg=0. Point color reflects statistical significance (−log10 P-value; see legend, scaled independently within each panel). Full summary statistics, including standard errors and exact P-values, are provided in data S29. (B) Meta-PheWAS network of UTI GWAS meta-analysis. Pleiotropic network of phenome-wide significant associations (P < 2.76×10^−5^) between 33 UTI risk loci and 247 phenotypes, identified by meta-PheWAS of 36 UTI risk loci across 2.9 million participants from seven multiethnic biobanks. Nodes represent either SNPs (gold) or phenotypes (green), with edges indicating concordance (red) or discordance (blue) of association as well as strength of association (width scaled by log-transformed effect size). Full association statistics are in data S30. An interactive version of the Meta-PheWAS and network map are viewable via a web browser at https://uti-phewas-browser-497008612026.us-central1.run.app/.

To further explore pleiotropic associations of individual UTI loci, we performed a meta-phenome-wide association study (Meta-PheWAS) for 36 index SNPs aggregated across 2.9 million participants of major biobanks (fig. S12, data S30). Loci associated with the broadest phenotypic spectra, spanning cardiometabolic, renal, neoplastic, vascular, and gastrointestinal domains, included *ALDH2* (58 phenotypes), *UMOD* (40 phenotypes), *CLPTM1L* (38 phenotypes), *ARHGAP1* (27 phenotypes), and *PSCA* (24 phenotypes). Of these, the *ALDH2* risk locus showed the broadest reach, with concordant associations spanning hypertension, atherosclerosis, and chronic liver disease, consistent with its known role in susceptibility to cardiometabolic disease (*51*). For multiple loci, we also detected significant pleiotropic associations with discordant effects relative to the UTI risk alleles. For example, the *PSCA* risk allele was associated with increased risk of duodenitis, peptic ulcer, and reflux esophagitis, but decreased risk of bladder and stomach cancers, consistent with the established associations of rs2294008 functional variant at this locus (*52–55*) in LD with the lead UTI SNP. Similarly, the *UMOD* risk allele was associated with increased risk of pyelonephritis and nephrolithiasis, but decreased risk of chronic kidney disease, hypertension, and kidney cancer. The *CLPTM1L* risk allele was associated with increased risk of basal cell carcinoma and lung cancer, but decreased risk of melanoma and pancreatic cancer. The *ARHGAP1* risk allele was associated with increased risk of aortic aneurysm and venous thromboembolism, but decreased risk of osteoporosis and skin cancer. Because many pleiotropic associations were shared across UTI risk loci, we built a pleiotropy map for all phenome-wide significant associations (Fig. 4B). An interactive version of the Meta-PheWAS and network map are available via a web browser at https://uti-phewas-browser-497008612026.us-central1.run.app/.

### PSCA as a constitutive epithelial defense gene against UTIs

Our integrative gene prioritization framework provided the strongest evidence for *PSCA* as the most likely causal gene with a protective effect against urinary pathogens (Fig. 5). *PSCA* encodes Prostate Stem Cell Antigen, a 123-amino-acid GPI-anchored protein expressed in the bladder, prostate, esophagus, and stomach (*56*, *57*). Although PSCA overexpression has been reported in prostate cancer (*56*, *58*), its physiological role in normal urinary tract remains poorly understood. Notably, *PSCA* is highly expressed in specialized prostate epithelial progenitor cells with immunomodulatory potential (*59*), and in a population of stomach pit cells where *H. pylori* is preferentially bound (*60*), hinting at broader roles in epithelial defense. These findings were further corroborated by our meta-PheWAS, demonstrating concordant effect of the *PSCA* locus on the risk of duodenitis and peptic ulcer disease.

**Fig. 5.**
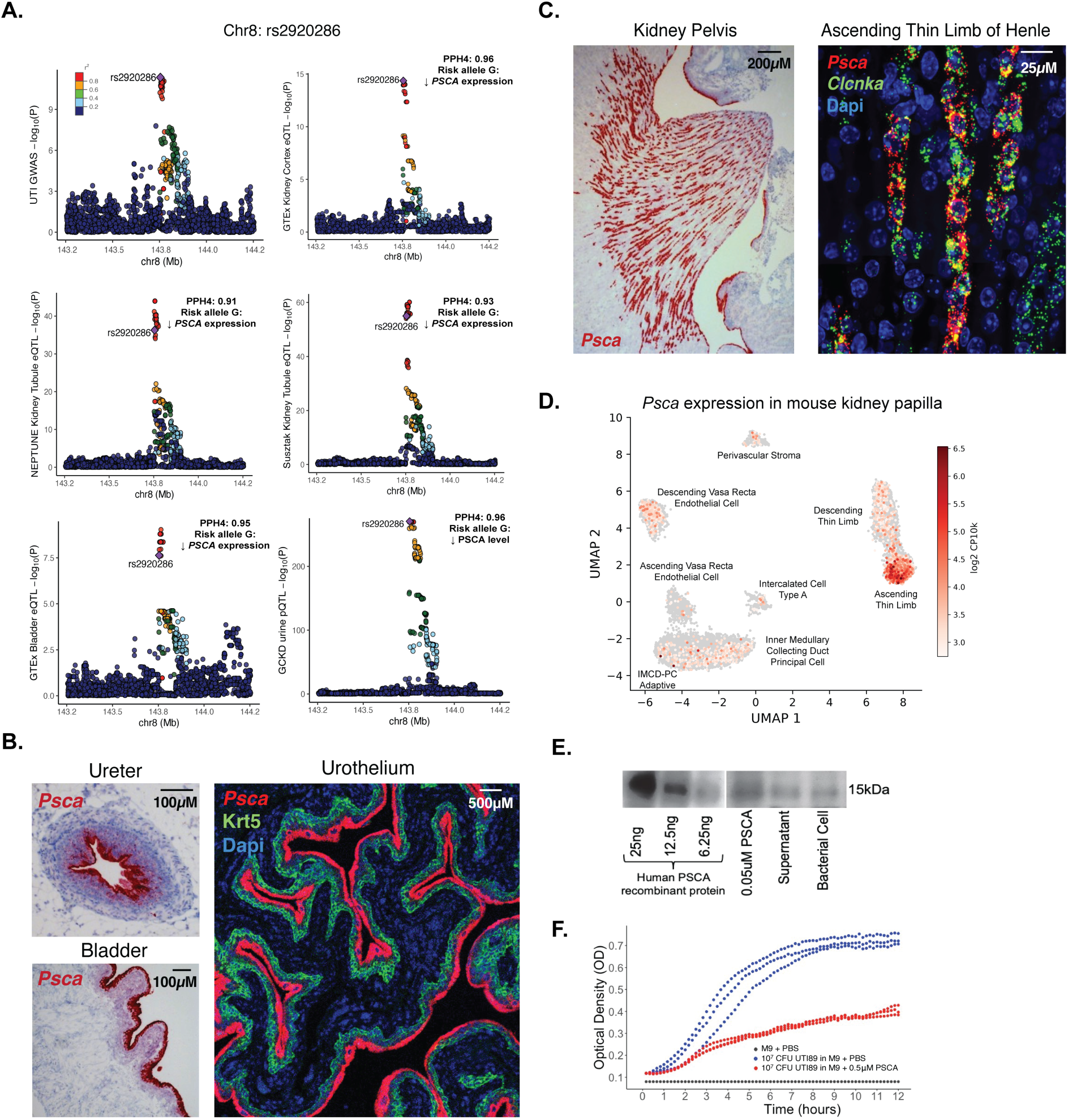
PSCA as a constitutive epithelial defense gene against UTI. **(A)** Regional plots show colocalization of the UTI GWAS signal at the PSCA locus with GTEx kidney cortex eQTL (PP4=0.96), NEPTUNE kidney tubule eQTL (PP4=0.91), Susztak kidney tubule eQTL (PP4=0.93), GTEx bladder eQTL (PP4=0.95), and GCKD urine pQTL (PP4=0.96), all demonstrating that the risk allele rs2920286-G is associated with decreased *PSCA* expression and reduced urinary PSCA levels. **(B)** RNA in situ hybridization (RNAscope) showing *Psca* expression (red) in mouse ureter (scale bar 100μM) and bladder (scale bar 100μM), and immunofluorescence of mouse urothelium co-stained with Psca (red), Krt5 (green), and DAPI (blue, scale bar 500μM). **(C)** RNAscope of mouse kidney pelvis showing *Psca* expression (red) in the kidney papilla (scale bar 200μM, left), and co-staining of *Psca* (red) with *Clcnka* (green) and DAPI (blue) confirming *Psca* expression is confined to ascending thin limb (ATL) cells of the loop of Henle (scale bar 25μM, right). **(D)** UMAP of mouse kidney papilla single-cell RNA sequencing data showing *Psca* expression enriched most prominently in the ascending thin limb (ATL) cell cluster with diffuse expression in other clusters. **(E)** Western blot demonstrating direct binding of human recombinant PSCA protein to *E. coli in vitro*, with human recombinant PSCA protein (0.05μM) at ∼15kDa present in bacterial supernatant, as well as the bacterial cell fraction after multiple washes (5x). **(F)** *In vitro* bacterial growth assay showing optical density (OD) over 12 hours for *E. coli* UTI89 grown in M9 minimal media with PBS (black), 10^7^ CFU UTI89 in M9 with PBS (blue), and 10^7^ CFU UTI89 in M9 with 0.5μM recombinant PSCA (red), demonstrating overall inhibition of bacterial growth by PSCA (estimated mean difference=-0.237±0.023, P=5.0×10^−4^, generalized additive mixed model).

Building on these observations, we investigated PSCA in the context of UTI to define its regulatory mechanisms, expression patterns, and potential functions in urothelial host defenses. We demonstrated that PSCA locus intersected kidney tubule open chromatin peaks (fig. S7, data S10 and S12) and co-localized with kidney tubule, bladder, and prostate eQTLs, as well as one of the strongest pQTLs in human urine (Fig. 5A, fig. S13). These findings support the hypothesis that the UTI risk allele suppresses both PSCA gene expression and urinary protein excretion.

We confirmed that in the mouse urinary tract, *Psca* was expressed in superficial bladder cells and ureteral epithelia (Fig. 5B). However, we also observed striking enrichment in the kidney papilla (Fig. 5C). Within the papilla, we demonstrated that the expression was confined to *Clcnka*⁺ ascending thin limb (ATL) of the loop of Henle (Fig. 5C, fig. S14). This is also consistent with our papillary scRNA-seq data confirming *Psca* expression exclusively in the ATL cell cluster (Fig. 5D). Because *E. coli* preferentially adheres to papillary epithelia (fig. S15A), PSCA occupies an ideal position at the host-pathogen interface. The papilla is also where the kidney injury and antimicrobial marker, *Lcn2* (Ngal) (*61*), is rapidly induced 24 hours after transurethral *E. coli* inoculation in a UTI mouse model (fig. S15B). We demonstrated that *Psca* expression in the papilla remained stable during the course of UTI and the expression pattern did not overlap with *Lcn2*, which was restricted to collecting ducts. These findings suggest that *Psca* expression is constitutive rather than part of an acute antimicrobial response to UTI and likely distinct in regulation.

Similar to UMOD, PSCA protein is anchored to the apical cell surface by a GPI linkage and can be cleaved and released into the urine. By Western blotting, we demonstrated that PSCA (∼12 kDa natively) was readily detected as a range of multiple molecular weight glycoforms (∼20-55kDa) in both UTI-positive and UTI-negative urine samples from human cohorts (*62*, *63*) (fig. S16A). When we removed N-linked glycans enzymatically, these bands collapsed into lower-molecular-weight forms (fig. S16B), confirming that urinary PSCA is heavily N-glycosylated. We then demonstrated that human recombinant PSCA protein directly bound *E. coli in vitro*, even after extensive washing (Fig. 5E). Lastly, we performed *in vitro* bacterial growth suppression assays and demonstrated that recombinant PSCA significantly inhibited bacterial growth relative to the PBS control (estimated mean difference=-0.237±0.023 OD600, P=5.0×10^−4^, Fig. 5F). Together, these findings position PSCA as a constitutive papillary GPI-linked glycoprotein that resides precisely where bacteria attach, can be shed into urine, and directly impedes bacterial proliferation. This convergence of genetic, regulatory, and functional evidence highlights PSCA as a novel epithelial defense factor in the urinary tract.

## DISCUSSION

In this study, we define the genetic architecture of urinary tract infection through the GWAS meta-analysis encompassing 1.86 million individuals. We identified 36 novel independent genome-wide significant loci, including *PSCA*, *UMOD*, and *FUT2*, that collectively illuminate epithelial and immune mechanisms of host defense. These genes are distributed throughout the length of the urinary tract, from the kidney papilla to the bladder, and rather than being induced in response to infection, most appear to act constitutively, at steady state, providing an inherited defense that is already in place before bacteria arrive. By integrating variants and their proxies with regulatory annotations, cell-type-specific transcriptomes, and urinary tract eQTL and pQTL colocalizations, we describe a convergent landscape in which epithelial barrier programs and innate immune regulation shape susceptibility to UTIs.

UTI risk alleles appear to have predominantly regulatory effects, with lead variants enriched in distal enhancer-like elements, consistent with the growing recognition that common disease variants frequently influence gene regulation through enhancer activity rather than coding changes. Loci mapping to epithelial surface and barrier proteins (*PSCA*, *UMOD*, *CLPTM1L*, *FUT2*), innate immune or inflammatory regulators (*BTN3A2*, *FES*, *HOTAIR, PTPRC, NFATC1*), and developmental determinants of mucosal architecture (*FGFR2*, *BMP7*, *GLIS2*, *ZFHX3*) suggest that infection risk arises from a convergence of epithelial defense, immune dysregulation, and heritable variation in urinary tract development (table S2). Colocalization analyses indicate that approximately one-third of loci modulate gene expression in kidney tubules, which are directly exposed to uropathogens, with directional effects showing risk alleles that often downregulate epithelial effectors and upregulate inflammatory mediators.

Among all signals, *PSCA* (Chr8: rs2920286) emerged as the most likely novel causal gene. *PSCA* encodes a 123-amino-acid GPI-anchored glycoprotein expressed in the prostate, bladder, esophagus, and stomach. We demonstrate that in the urinary tract, *PSCA* is strikingly enriched in the ascending thin limb cells of the kidney papilla and in umbrella cells of the bladder and ureteral epithelia, all interfaces for ascending urinary infection. We find that similar to uromodulin, PSCA is shed into the urine as heavily glycosylated forms that can bind *E. coli* and suppress bacterial growth *in vitro*, and risk alleles are associated with reduced kidney and bladder gene expression and reduced urinary protein abundance. This association can be explained in part by rs2294008, a variant in high LD with our index SNP rs2920286 at the *PSCA* locus: the risk-associated C allele lacks an upstream start codon present in the protective T allele, defaulting to a cytoplasmic form that is rapidly degraded, while the protective T allele introduces this upstream A<u>T</u>G, adding a signal peptide that directs *PSCA* to the cell surface (*54*). These data nominate *PSCA* as a constitutive epithelial effector with antibacterial function in the urinary tract.

The *PSCA* locus further exhibits pleiotropic architecture, conferring protection against urinary tract infection and peptic ulcer disease, duodenitis, gastritis, and esophagitis, with *E. coli* and *H. pylori* serving as the dominant pathogens at each respective mucosal site. At the same time, this locus predisposes to bladder, prostate, and gastric cancers. This pattern of antagonistic pleiotropy may reflect an evolutionary trade-off in which the allele conferring maximal mucosal defense carries oncogenic risk. This divergence may reflect *PSCA*’s identity as a marker of terminal epithelial differentiation across mucosal surfaces, a state that can itself become a bacterial target. In the stomach, *H. pylori* preferentially colonize *PSCA*-high pit cells (*60*) and actively suppresses *PSCA* expression during infection (*64*), suggesting a cycle in which colonization erodes the differentiated barrier state marked by *PSCA*. The same logic may extend to the urothelium, where *PSCA* marks umbrella cells (the terminally differentiated apical layer constituting the primary barrier to UPEC adhesion) such that lower *PSCA* abundance may reflect a less intact barrier prior to or during infection. Notably, recurrent UTI is associated with reduced urothelial proliferation markers even after clinical recovery (*65*), consistent with impaired epithelial regeneration as a contributor to susceptibility. PSCA’s established role in regulating cell proliferation across mucosal tissues may thus bridge its antimicrobial and oncogenic associations, providing a plausible explanation for the divergent phenotypes at this locus.

The GPI-anchored glycoprotein nature of *PSCA*, enabling its apical surface localization and release into urine as a soluble decoy, connects it to a broader mucosal glycocalyx defense network. Analogous to *UMOD* (Chr16: rs12917707), another GPI-anchored protein which aggregates piliated bacteria (*9*), this network may provide a dual mechanism for epithelial bacterial capture and urinary clearance. *CLPTM1L* (Chr5: rs381949) facilitates ER processing and trafficking of GPI-linked proteins to the cell surface (*66*), potentially regulating insertion of PSCA and UMOD into cholesterol-rich lipid raft domains. Though GPI-anchored proteins lack transmembrane domains, they can transduce signals through raft-resident receptors. For example, PSCA has been shown to induce IL-6 via p38/NF-κB signaling (*67*) suggesting it may couple bacterial capture to downstream inflammatory responses in addition to its decoy function.

The glycan dimension of this mucosal glycocalyx defense extends further to *FUT2* (Chr19: rs507766), whose fucosylation of uroepithelial glycoproteins (*68*) adds a second layer of surface glycan remodeling with direct implications for pathogen attachment. *FUT2* encodes α-1,2-fucosyltransferase, which adds fucose to the terminal galactose of precursor glycans on mucins and other secreted and cell-surface glycoproteins in epithelial and secretory tissues, generating ABO and H blood group antigens outside the erythroid lineage (where the paralog *FUT1* predominates instead). The canonical loss-of-function variant rs601338 (G461A, p.W154*, data S6) abolishes enzyme activity and produces the “non-secretor” phenotype, characterized by absence of ABO/H antigens in saliva and other mucosal secretions. The lead UTI risk variant at this locus, rs507766-T, is in high LD with rs601338-G, such that the UTI risk tracks with functional FUT2 and the secretor phenotype. This direction of effect contrasts with earlier clinical studies that associated non-secretor status with increased UTI susceptibility in women (*69*, *70*). However, those studies were limited by small sample sizes and, in some reports, significant associations were only observed in specific subsets (*69*, *70*). Nevertheless, the direction of effect observed in our study is consistent with evidence supporting secretor status as a risk factor for a range of mucosal infections that exploit fucosylated antigens as attachment receptors, including rotavirus (*71*), norovirus (*71*, *72*), SARS-CoV-2 (*73*), *H. pylori* (*74*), and pathogens that cause otitis media (*23*, *75*). Mechanistically, *H. pylori* infection induces rapid upregulation of *FUT1*/*FUT2* and increased gastric epithelial display of fucosylated glycans within hours of exposure (*76*). By analogy, FUT2 activity in the urinary tract could generate fucosylated glycoconjugates on uroepithelial mucins that serve as attachment sites for UPEC adhesins. Together, these genes suggest that CLPTM1L-dependent GPI anchoring, PSCA surface display, and FUT2-mediated glycosylation form a coherent axis of mucosal protection. We propose that this GPI-anchored glycoprotein complex mediates (i) bacterial capture at the epithelial surface, (ii) downstream inflammatory signaling, and (iii) decoy secretion via shed glycoproteins in urine.

Beyond these lead loci, several other prioritized genes illuminate further layers of host defense that span five additional mechanistic themes: innate immune activation, epithelial cell death and infection resolution, epithelial identity and barrier integrity, kidney and urinary tract developmental programs, and nutritional immunity (summarized in table S2). Beyond these themes, pleiotropic signals across epithelial and immune loci further linked UTI susceptibility to gastrointestinal, gynecologic, and respiratory phenotypes. These shared associations highlight conserved genetic programs governing mucosal integrity across organ systems.

Despite unprecedented power, ancestry-specific and sex-specific resolution remains limited due to lower sample sizes of subgroup analyses. Additionally, intrinsic limitations of the EHR data, such as variable coding practices, unequal follow-up time, and non-random data missingness, likely lead to the dilution of effect size estimates for the detected loci. Integrating environmental factors, including lifestyle factors, urinary microbiome composition, and antibiotic exposures, may further refine understanding of UTI risk. Mechanistic understanding through mouse knockout models of *PSCA* and other mucosal glycocalyx-associated genes will be essential to validate the causal pathways suggested by our GWAS.

By integrating genetic, regulatory, and functional evidence, our study demonstrates that epithelial surface programs, innate immune regulation, developmental patterning, and epithelial nutritional immunity collectively represent inherited determinants of UTI susceptibility, most operating at steady state rather than being induced by infection. *PSCA* emerges as a constitutive epithelial defense factor operating within a GPI-anchored decoy framework that includes *CLPTML1*, *FUT2*, and *UMOD*. The antagonistic pleiotropy at the *PSCA* locus suggests evolutionary trade-offs between mucosal defense and cancer, highlighting the dynamic balance between infection resistance and tissue renewal. Our findings highlight host barrier-centered mechanisms of susceptibility and nominate numerous actionable targets for both prophylaxis in at-risk patients and broader non-antibiotic prevention of urinary tract infections.

## MATERIALS AND METHODS

### Electronic phenotyping for urinary tract infections

We developed an electronic phenotyping algorithm (e-phenotype) to define UTI cases and non-UTI controls using diagnostic codes and available urine tests in electronic health record (EHR)-linked biobank datasets from Electronic Medical Records and Genomics-III (eMERGE-III) (*17*), UK Biobank (UKBB) (*19*), BioVU (*20*), and All-of-Us (AoU) (*18*), (fig. S1). We excluded any cases with suspected secondary causes of UTIs such as urethritis, obstruction, catheter-related and nosocomial infections, congenital/genetic anomalies, transplant, cancer, or immunodeficiency. For the biobanks with available diagnostic timestamps (eMERGE, BioVU, AoU), we required at least two UTI-defining billing codes more than 7 days apart to define cases. If no timestamp was available (e.g. UKBB), two occurrences of a billing code were required to define cases regardless of their temporal relationship. We excluded any cases with a single UTI event/single diagnostic code. The non-UTI controls were those without any qualifying diagnostic codes and without any positive urine cultures or urine bacteria on urinalysis data. We validated the accuracy of our phenotyping through a manual review of randomly selected cases, which confirmed a positive predictive value >82%. Details of the UTI algorithm along with the related code, lists of diagnostic and procedure codes, and validation are available at https://phekb.org/phenotype/urinary-tract-infection-uti-electronic-phenotype-e-phenotype-algorithm.

### Genome wide association study (GWAS)

Detailed descriptions of each cohort are provided in Supplementary Notes. In each biobank, we applied stringent genotype QC filters, including only high confidence imputed markers (R^2^>0.8), removing low-quality samples, ancestry outliers, and cryptically related individuals. We applied a uniform QC pipeline for all cohorts using a combination of VCFtools (*77*), SAIGE (*78*), and PLINK (*79*). Each cohort was further stratified by genetically-inferred sex (male/female) and continental genetic ancestry cluster as defined by co-clustering with 1000G reference populations. Case-control genetic association tests were performed using logistic regression under additive genetic model (dosage genotype coding) within each sex and continental ancestry-defined stratum separately, and with adjustment for significant PCs of ancestry. GWAS summary statistics were then combined using inverse-variance-weighted fixed-effects meta-analysis with METAL(*80*). Subgroup meta-analyses by genetic ancestry and sex were also performed to define ancestry-specific and sex-specific loci. We examined QQ-plots and genomic inflation for each meta-analysis. The genomic inflation was λ = 1.27 for the overall meta-analysis, λ = 1.27 for the European-only analysis, and λ = 1.19 and 1.17 for male and female only analyses, respectively. The intercept from the LDSC regression was 1.1, suggesting that mild genomic inflation is likely due to polygenicity rather than population stratification. Genome-wide significant signals were defined by the generally accepted P < 5.0 × 10^−8^ significance threshold, and signals with P < 1.0 × 10^−6^ were defined as suggestive.

### Genomic and regulatory annotations of UTI GWAS loci

For genetic annotations of our GWAS loci, we curated variants in strong LD with the 36 index SNPs based on external reference panels (r2 ≥ 0.7, 1000G all ancestries), which yielded 730 total SNPs. These SNPs were subsequently annotated using ANNOVAR (*25*) and VEP (*26*) to define any coding, splicing, and 3’UTR and 5’UTR variants and their predicted effects. GENCODE v49 annotations were used to identify the nearest TSS. To assess the regulatory impact of noncoding variants on gene expression, we applied PromoterAI (*27*), a trained model that predicts allele-specific effects of promoter variants on gene expression; variant-gene associations with absolute PromoterAI scores ≥ 0.1 were prioritized for further investigation.

### ENCODE4 registry

Non-coding variants were evaluated for overlap with candidate cis-regulatory elements (cCREs) from the ENCODE4 project (registry v4, GRCh38/hg38) (*29*). A total of 2,348,854 cCREs spanning 1,888 biosamples were assessed from the ENCODE SCREEN portal (https://screen.wenglab.org/; accessed March 6, 2026). Overlap between 36 UTI GWAS index SNPs and their proxies (N=730) and cCRE coordinates was determined by positional intersection across all biosamples, retaining variants whose genomic position fell within a cCRE interval. Intersecting variants were classified across all eight biosample-specific cCRE activity categories: promoter-like signatures (PLS), proximal and distal enhancer-like signatures (pELS and dELS), chromatin accessible with H3K4me3 (CA-H3K4me3), chromatin accessible with CTCF binding (CA-CTCF), chromatin accessible with transcription factor binding (CA-TF), chromatin accessible only (CA), and transcription factor binding only (TF).

### Activity-by-contact (ABC) enhancer-gene maps

Significant GWAS loci index SNPs and their proxies (N=730) were intersected with enhancer regions from Activity-by-Contact (ABC) enhancer-gene maps across 131 human biosamples (accessed via https://github.com/EngreitzLab/V2G). The ABC model predicts enhancer-gene links by integrating chromatin accessibility (ATAC-seq/DNase-seq) and H3K27ac signals as measures of enhancer activity, with Hi-C-derived contact frequency as a measure of enhancer-promoter interaction (*30*). Variants overlapping ABC enhancer regions with ABC scores >0.02 were retained as SNPs with predicted regulatory activity.

### Single cell enhancer-to-gene (scE2G) mapping in human kidney cells

To link non-coding UTI GWAS variants to regulatory targets in kidney tissue, we constructed cell-type-resolved enhancer-to-gene (E2G) maps using 10x Multiome data from 25 healthy kidney samples from the Kidney Precision Medicine Project (KPMP) (*32*). Cell-type annotations were assigned via label transfer using 38 healthy reference samples from the KPMP Atlas V2.0 snRNA-seq (*33*). We defined 12 kidney epithelial cell types and an additional 9 stromal, vascular, and immune populations, and cell clusters were validated using established canonical marker genes (*36*). Enhancer-gene links were predicted for 17 cell clusters with >10 cells in the KPMP 10x Multiome dataset using the scE2G Multiome framework (*34*). The scE2G model is a supervised classifier that scores candidate element-gene pairs, defined as ATAC peaks within 5 Mb of a gene’s TSS and using eight features: (1) a pseudobulk ABC score with 3D contact estimated as the inverse of genomic distance; (2) the Kendall correlation between element accessibility and gene expression across single cells; (3) a measure of whether the gene is ubiquitously expressed; and (4–8) additional features describing genomic distance and chromatin accessibility around the regulatory element. Element-gene pairs with a score >0.171, corresponding to 70% recall against CRISPRi-validated enhancer–gene pairs, were retained as high-confidence regulatory interactions. UTI GWAS SNPs and their proxies (N=730) were subsequently intersected with scE2G peak-gene regions to identify variants overlapping predicted regulatory elements and their associated target genes with kidney cell type resolution.

### Open4Gene (O4G) chromatin accessibility-gene links in human kidney cells

We obtained orthogonal enhancer-gene mappings based on the Open4Gene (O4G) dataset derived from paired single-nucleus ATAC-seq and RNA-seq profiles of 30 human kidney samples (*35*). This dataset links chromatin accessibility peaks to nearby gene expression using a hurdle regression framework applied within matched cell types. Genes located within ±250 kb of each lead SNP were considered. Our 36 GWAS index SNPs and proxies (N=730) were intersected with accessible chromatin peaks, and peak-gene associations with Zero-Inflated Negative Binomial (ZINB) count FDR <0.05 or ZINB zero FDR <0.05 were used for gene prioritization.

### Single nuclei RNA sequencing of mouse kidney papilla

Left and right kidney papillae (2 mg each) were micro-dissected from a healthy 10-week-old female C57Bl/6 mouse. Nuclei were isolated by dounce homogenization in 1 mL of buffer containing 250 mM sucrose, 25 mM KCl, 5 mM MgCl₂, and 10 mM Tris (pH 8.0), 1x cOmplete EDTA-free protease inhibitor cocktail (Roche Cat No: 04693132001), 0.2 U µl^−1^ SUPERase·In, centrifuged at 4 °C for 5 min at 300*g*, followed by resuspension in PBS. Nuclei suspensions were washed using a Curiox Laminar Flow system (5 μL/s flow rate, 20 cycles with PBS) to remove debris and improve purity. Single-nuclei suspensions were loaded into the 10x Genomics Chromium Controller (Single Cell 3′ v3.1 chemistry). Libraries were prepared by cDNA synthesis and Nextera-based library construction, according to manufacturer protocols. Sequencing was performed on the Element Aviti system using paired-end 75 bp reads. Raw snRNA-seq data were processed using a previously reported pipeline (*81*) (DropSeqPipeline8.py; code availability at https://github.com/simslab/DropSeqPipeline8). Sequencing reads were aligned to the mouse reference genome (GRCm38, Gencode vM10). Nuclei were retained if they had ≥1,000 UMIs, ≥650 detected genes, and ≤2% mitochondrial fraction, yielding 20,681 nuclei. Using previously described methods (*82–85*) (cluster_diffex.py; full code and pipeline availability at https://github.com/simslab/cluster_diffex2018), highly variable genes were identified by deviation from the gene dropout curve and used to construct a Spearman’s correlation matrix and *k*-nearest neighbor graph, from which unsupervised clustering was performed via Louvain community detection (Phenograph) and visualized with UMAP. Clusters containing multiplets (identified by marker expression inspection and validated with scDblFinder (*86*)) and clusters consistent with thick ascending limb of Henle (TAL) or proximal tubule populations (PT), representing contamination from microdissection, were removed, excluding 2,289 nuclei in total and yielding a final dataset of 18,392 papillary nuclei. Final clustering of the filtered dataset produced 8 transcriptionally distinct populations: inner medullary collecting duct principal cells (IMCD-PC), adaptive inner medullary collecting duct principal cells (aIMCD-PC), intercalated cells type A (IC-A), descending thin limb of Henle (DTL), ascending thin limb of Henle (ATL), descending vasa recta (DVR), ascending vasa recta (AVR), and mixed fibroblasts and pericytes (Stroma). Cell type identities were assigned based on cluster-enriched marker genes and validated against the KPMP snRNA-seq human kidney reference (*36*).

### Urinary tract gene expression of candidate GWAS genes in mouse and human datasets

Genes within 500kb of the lead GWAS SNPs were interrogated for gene expression in relevant kidney and urinary tract tissues using our mouse kidney papilla snRNA-seq dataset, as well as published transcriptomic atlases of kidney (Kidney Precision Medicine Project; KPMP) (*36*), ureter (*37*), bladder (*38*), urine (*39*), and immune (*40*) cells. Each dataset was processed as follows:

***Mouse kidney papilla:*** snRNA-seq counts comprising 18,392 nuclei from a healthy mouse kidney papilla were normalized to log2(CP10k+1). Cell type labels were assigned from previously described clustering analysis to annotate the nuclei (see above Methods for snRNA-seq).

***Human kidney*:** Human kidney snRNA-seq aggregated clustered data from Kidney Precision Medicine Project (KPMP) comprising 615,169 nuclei from 45 healthy reference donors and 48 patients with kidney disease (*36*). Normalized expression values were extracted directly from the Seurat-normalized h5Seurat file (SingleNucleus_KPMP_Explorer_05182025.h5Seurat; log1p(CP10k) as computed by the KPMP pipeline). Cell type annotations were decoded from the *meta.data/subclass.l2* factor field and subsequently collapsed into harmonized categories to compute average expression.

***Human ureter*:** Human ureter scRNA-seq dataset comprising 30,942 cells from 10 individuals were obtained as count matrices for global, urothelial, stromal, and immune ureter subsets (*37*). These count matrices were normalized to log2(CP10k+1), and cell type labels were obtained from the accompanying metadata files and subsequently collapsed into harmonized categories to compute average expression.

***Human bladder*:** Human bladder scRNA-seq dataset (*38*) comprising 12,423 cells from 3 individuals were obtained as 10x Genomics sparse matrix files without cell type annotations per barcode. These count matrices were merged and normalized to log2(CP10k+1) for tissue-level analysis only. Cell-type-level expression and dot plot visualization used published per-cluster average expression values and summary statistics. Cluster subtypes were collapsed into harmonized categories, and average expression was computed as the mean across constituent clusters.

***Human urine:*** Human urine scRNA-seq dataset (*39*) comprising 23,082 urinary cells from 5 individuals with diabetic kidney disease and a pooled reference sample from 10 healthy individuals were obtained as a raw count matrix and normalized to log2(CP10k+1), analyzed at the tissue level only.

***Human peripheral blood mononuclear cells (PBMCs):*** Reference PBMC scRNA-seq data (*40*) from 163 healthy individuals, comprising 710,654 cells, were obtained as a clustered Seurat object. Normalized expression values (as computed by Seurat’s “LogNormalize” method: log1p(CP10k)) were extracted, and cell type annotations were decoded from the cluster-level metadata field and subsequently collapsed into harmonized categories to compute average expression.

***Gene Expression Detection Threshold:*** For datasets with raw count matrices, a per-gene noise threshold was defined as the 95th percentile of a Gaussian fit to cells with expression below 0.1 (mean + 1.645 SD). A gene was called expressed in a tissue or urine if: (i) ≥1% of all cells exceeded the threshold (tissue-wide), or (ii) ≥10% of cells in any single cluster exceeded the threshold (cluster-specific). For human bladder and human urine datasets, genes present in the published summary statistics (pct.1 > 0.25 in at least one cluster) were used in place of the noise threshold approach and assigned a positive expression call.

***Dot Plot Visualization:*** Dot plots were generated for the 36 top-prioritized GWAS candidate genes (top scoring candidate gene per locus). Dot color represents row-normalized average expression, where each gene’s mean log2 CP10k is scaled to [0, 1] relative to its maximum-expressing cell type, using dataset-specific sequential color scales: magenta-purple (mouse kidney papilla), red (human kidney), green (human ureter), blue (human bladder), gold (human urine), and purple (human PBMCs). Dot size represents the percentage of cells per cell type exceeding a per-gene noise threshold (95th percentile of a Gaussian fit to the low-expressing cell population); for bladder, dot size reflects relative expression scaled to the maximum-expressing cell type, as only cluster-level summary statistics were available.

### Lookups across QTL datasets

Lead UTI GWAS variants were directly queried for QTL effects (at P<0.05) against kidney tubule and glomerular eQTLs from the Susztak lab (*43*), kidney tubulointerstitial and glomerular eQTLs from the NephQTL consortium (*42*, *87*), and GTEx v10 (*41*) eQTLs and sQTLs from kidney cortex, bladder, prostate, and whole blood tissues.

### Colocalization of UTI loci with QTLs

Colocalization analyses between UTI GWAS and specific QTL datasets were performed using the Coloc package in R (v5.2.3). Genomic intervals of ±500 kb were defined for top 36 lead SNPs from our UTI GWAS. For each locus, we tested colocalization with multiple published tissue-specific QTL datasets, including (i) kidney tubule eQTLs (*43*) from the Susztak lab, (ii) kidney tubulointerstitial eQTLs from the NephQTL consortium (*42*, *87*), (iii) GTEx v10 kidney cortex, bladder, prostate, and whole blood eQTLs and sQTLs (*41*), (iv) whole blood eQTL meta-analysis from 31,684 individuals (*45*), (v) immune cell eQTLs from the CEDAR study (*46*), including monocytes (CD14+), neutrophils (CD15+), CD4+ and CD8+ T cells, B cells (CD19+), and (vi) urine pQTL from the German Chronic Kidney Disease (GCKD) study (*47*, *88*). After harmonization of effect alleles, we identified all colocalized QTLs mapping to the ±500 kb region of the index SNP. For each locus-QTL pair, coloc was run under the default single causal variant assumption, and posterior probabilities were estimated for five models: H0 (no association in either trait), H1 (association in GWAS only), H2 (association in QTL only), H3 (independent associations), and H4 (shared causal variant). Colocalization was considered strongly supported if the posterior probability for a shared causal variant (PP4) exceeded 0.80.

### Proteome-wide association study (PWAS)

We tested the effects of genetically predicted protein expressions on UTI using PWAS and employed the *BLISS* framework (*89*), which builds protein prediction models from pQTL summary statistics derived from the UK Biobank, deCODE, and ARIC studies. Specifically, we applied protein prediction models trained on the UK Biobank Pharma Proteomics Project (UKBPPP) (*48*), using data from 49,341 individuals of European ancestry and encompassing 2,808 Olink-measured proteins. We restricted our analysis to 1,407 proteins with estimated cis-heritability >0.01 and evaluated the associations between 1,398 genetically predicted protein expression levels and UTI leveraging the following three summary statistics: (1) GWAS including all 1,819,456 individuals; (2) European-ancestry-specific GWAS of 1,543,656 individuals; and (3) male-specific GWAS of 888,849 individuals. Bonferroni-corrected *P*<0.05 was used to define significant associations.

### Intersection of GWAS loci with mouse bladder transcriptome in UTI

To identify genes with functional relevance to UTI within GWAS loci, we intersected genes within ±500kb of each lead GWAS variant with differentially expressed genes from a nascent RNA transcriptomic dataset of bladder urothelial cells generated in a parallel study. Briefly, the study employed a thiouracil-based labeling approach (*90*) in mice expressing the UPRT enzyme under the Uroplakin 2 (Upk2-Cre) promoter to target RNA labeling to bladder urothelium, with two cohorts of 8-week-old female mice (Cohort 1: UTI n=5, PBS n=5; Cohort 2: UTI n=3, PBS n=5). Genes within GWAS loci that were differentially expressed (FDR<0.05) between UTI and control conditions in that dataset were prioritized as candidate effector genes.

### Gene prioritization via mouse knockout dataset and literature review

Candidate genes within GWAS loci were functionally annotated using the Mouse Genome Informatics (MGI) Resource (*49*) to identify genes with mouse knockout phenotypes relevant to urinary tract or immune mechanisms. Complementary manual literature searches were performed using PubMed, Google Scholar, and Harmonizome 3.0 (*50*) to identify published functional evidence relating to the urinary tract or immune response in the context of UTI. Additional gene annotations including function, disease association, pathway, and protein-level information were retrieved from UniProt.

### Gene prioritization using large language models (LLM)

To systematically prioritize candidate causal genes, we employed an LLM-based approach adapted from Shringarpure et al. (*91*). Genes within ±500 kb of each 36 lead GWAS variants were submitted to three LLMs: ChatGPT (V5.5), Claude (V1.8), and Gemini (V3.1), using a standardized prompt instructing each model to identify the most likely causal gene per locus based on biological and literature evidence for UTI susceptibility. Each LLM was prompted to perform 20 independent runs per locus, returning a causal gene, confidence score (0–1), and brief rationale in JSON format, with the gene receiving the most votes across runs selected as that model’s top candidate. See system prompt below. The final prioritized gene per locus was determined by a majority vote across the three LLMs, with ties resulting in 2 or 3 co-prioritized genes. LLM-prioritized genes were then compared against manual literature curation to evaluate prioritization concordance.

### System prompt

*“You are an expert in biology and genetics. Your task is to identify likely causal genes within a locus for a given GWAS phenotype based on literature evidence. From the list, provide the likely causal gene (matching one of the given genes), confidence (0: very unsure to 1: very confident), and a brief reason (50 words or less) for your choice. Return your response in JSON format, excluding the GWAS phenotype name and gene list in the locus. JSON keys should be ’causal_gene’,’confidence’,’reason’. Your response must start with ’{’ and end with ’}’. Run this 20 times and treat each run as independent from all previous runs. Do not try to maintain consistency with earlier outputs. Base your decision only on general biological knowledge and literature. Report a vote of confidence in the causal gene that is most consistently and stably chosen among all runs. Use the following set of genes within the 36 GWAS loci. Prioritize one gene per locus. The phenotype is Urinary Tract Infections.”*

### Integrative gene prioritization framework

To systematically prioritize candidate causal genes across 36 UTI GWAS loci, we developed an integrative scoring framework encompassing 229 candidate genes within ±500kb of each lead GWAS variant. Each gene was evaluated across 25 binary features (1 point per feature, maximum score of 25) spanning four different categories of evidence: genomic, tissue-specific, genetic, and functional as described in the Results. Each gene received a cumulative score by summing binary scores across all 25 features. Scores were then aggregated into a ranked gene prioritization matrix, with the top-scoring gene per locus designated as the primary candidate. In cases where two or more genes within a locus received equivalent scores, the gene nearest to the lead GWAS variant was designated as the primary candidate.

### Genetic correlation analysis

Genome-wide genetic correlations between UTI and other complex traits and diseases were estimated using linkage disequilibrium score regression (LDSC) (*92*). Genetic correlation coefficients were calculated from GWAS summary statistics using precomputed linkage disequilibrium scores derived from all populations from the 1000 Genomes Project reference panel (*24*). Summary statistics were harmonized and filtered according to LDSC recommendations before analysis.

### Meta-phenome wide association studies (Meta-PheWAS)

Phenome-wide association analyses were conducted across multiple large-scale biobank cohorts, including eMERGE, UK Biobank, and the AoU Research Program, each of which provides linked electronic health record and genetic data for genomic analyses. ICD diagnosis codes were mapped to standardized phecodes to harmonize phenotype definitions across datasets. Case definitions for each phecode required multiple diagnostic codes, and control groups were defined according to the standard PheWAS framework (*93*). Logistic regression models were used to test the association between SNP genotype and each phecode, adjusting for age, sex, study site, and principal components of ancestry. Association results were generated for all available phecodes within each cohort and subsequently combined using meta-analytic approaches across biobanks. We additionally obtained publicly available PheWAS summary statistics from the Million Veteran Program (*21*), Taiwan Precision Medicine Initiative (*94*), FinnGen (*22*), and Mass General Brigham (MGB) (*95*). All cohorts were harmonized to a common phecode mapping framework to ensure consistency of phenotype definitions across studies. Summary statistics were aligned by effect allele and standardized prior to meta-analysis. We performed a meta-phenome-wide association study (Meta-PheWAS) across all available biobanks by combining cohort-specific association results using fixed-effects inverse-variance weighted meta-analysis implemented in the PheWAS R package (metagen function). Statistical significance was defined using a Bonferroni-corrected threshold of P < 2.75 x 10^−5^ (0.05/1,817 phecodes tested). Meta PheWAS network was constructed by importing phenome-wide significant locus-phenotype associations from the meta-PheWAS (data S30) into Cytoscape (v3.10.4) as an edge list, with UTI risk loci (n=33 nodes) and associated phenotypes (n=247 nodes) defined as separate node types. Edge color was mapped to the direction of effect relative to the UTI risk allele (concordant=red, discordant=blue), and edge width was mapped to the log-scaled function of effect size. The network was initialized using the Prefuse Force Directed OpenCL layout in Cytoscape; node layout was then adjusted manually to minimize edge crossing and optimize readability.

### Mouse husbandry

Female and male C57BL/6 and C3H/HeN mice were purchased from Jackson Laboratory (Bar Harbor, ME) and housed and bred at Columbia University Irving Medical Center under protocol AC-AABN7563, approved by the Columbia University Institutional Animal Care and Use Committee.

### Urinary tract infection mouse model

Female and male C3H/HeN mice, 8-10 weeks of age, were used for uropathogenic *E. coli* (UPEC) infection. Briefly, anesthetized mice were transurethrally inoculated with 20-50 μl of UPEC strain UTI89-GFP (1×10⁹ CFU/ml) via a soft polyethylene catheter. Infection was confirmed by serial plating of urine collected directly from mice at 24 hours post-infection, with colony-forming units (CFU) enumerated on LB agar plates.

### Harvesting of mouse tissues

For in situ hybridization, kidney, ureter, and bladder tissues were harvested, fixed in 10% formalin overnight, and transferred to 70% ethanol prior to paraffin embedding. For immunofluorescence staining, tissues were fixed in 4% paraformaldehyde/0.1 M phosphate buffer at 4°C overnight, cryoprotected in 30% sucrose/0.1 M phosphate buffer at 4°C overnight, and embedded in O.C.T. Compound (Tissue-Tek).

### Mouse in situ hybridization (ISH)

ISH on FFPE mouse kidney tissues (5 μm) was performed using the chromogenic RNAscope 2.5 HD reagent kit (ACD, catalog 322350) and RNAscope 2.5 HD duplex reagent kit (ACD, catalog 322430) or the RNAscope Multiplex Fluorescent Reagent Kit v2 (ACD, catalog 323100) according to the manufacturer’s protocols. The following probes and antibodies of interest were used in various combinations on single, dual, or multi-probe channels: Mm-Psca-C1 and C2 (catalog 575301/-C2; 1:50 dilution for C2), Mm-Lcn2-C2 (catalog 313971-C2; 1:500 dilution), Mm-Clcnka-C1 (catalog 536031), Mm-Slc14a2-C1 (catalog 476721), goat anti mouse Aqp2 (Novus Biologicals NBP1-70378), rabbit anti mouse Atp6v1b1 (Proteintech 14780-1-AP), and chicken anti mouse Krt5 (1:400; Biolegend catalog 905904). Briefly, tissue sections were deparaffinized and rehydrated with xylene and ethanol washes, blocked with hydrogen peroxidase, heated in target retrieval buffer, and digested by protease. The tissue sections were then incubated with RNAscope target probe(s) at 40°C in a hybridization oven for 2 hours. RNA signal was amplified using the kit’s amplification system and detected using either alkaline phosphatase- or HRP-conjugated chromogenic substrates (RNAscope 2.5 HD and 2.5 HD Duplex kits) or HRP-mediated tyramide signal amplification with fluorophore-conjugated probes (RNAscope Multiplex Fluorescent v2 kit). For chromogenic assays, tissue sections were counterstained with hematoxylin, dried in a 60°C oven, and mounted for image analysis; for fluorescent assays, sections were counterstained with 4’,6-diamidino-2-phenylindole DAPI (1:1000) and mounted with Permafluor Mountant (Thermo Fisher Scientific). Bright-field images of chromogenic slides were captured using the Olympus IX73 inverted microscope under low (100×) and high (600×) magnifications, and fluorescent images were captured using a Zeiss LSM 710 confocal microscope, Leica SP8-DLSM light sheet microscope, or Nikon Ti2 inverted microscope (AXR).

### Immunofluorescent staining

Frozen sections of 20 μm were used for immunofluorescence staining with rat monoclonal anti mouse Krt8 (1:400; Developmental Studies Hybridoma Bank TROMA-I), goat anti *E. coli* serotype O/K antibody (1:200; Invitrogen PA1-73032), and chicken anti-GFP antibody (1:200; Abcam ab13970). Fluorescent secondary antibodies, including Alexa Fluor 488-AffiniPure F(ab′)2 Fragment IgG Donkey Anti-Rat IgG, Alexa Fluor 594-AffiniPure F(ab′)2 Fragment Donkey Anti-Goat, and Alexa Fluor 647-AffiniPure F(ab′)2 Fragment Donkey Anti-Chicken, (1:1000; Jackson ImmunoResearch Laboratories), were used for Krt8, *E. Coli*, and GFP identification, respectively. All slides were co-stained with DAPI to identify nuclei.

### Urinary PSCA immunoblotting

Urine samples from our previously published Columbia University Emergency Department (ED) cohort were assessed for PSCA expression by immunoblot in individuals with and without UTI, defined as leukocyte esterase-positive urine with >10,000 colony-forming units (CFU). Samples were resolved by 4-15% SDS-PAGE (Bio-Rad Laboratories) alongside serial dilutions of human PSCA recombinant protein (25, 12.5, and 6.25 ng) as a reference standard, and immunoblotted using rabbit anti-mouse PSCA primary antibody (1:1000; Invitrogen, catalog PA5-11463) and HRP-conjugated rabbit polyclonal secondary antibody (1:5000; Jackson ImmunoResearch, catalog 711-035-152).

### PNGase F Deglycosylation of Urinary PSCA

Human urine samples were treated with PNGase F (New England Biolabs, catalog P0704S) according to the manufacturer’s protocol. Briefly, samples were combined with Glycoprotein Denaturing Buffer (10X) and denatured at 100°C for 10 minutes. Denatured samples were chilled on ice, briefly centrifuged, and split into paired aliquots. GlycoBuffer 2 (10X) and 10% NP-40 were added to each aliquot to counteract SDS-mediated inhibition of PNGase F activity. PNGase F was added to one aliquot of each sample pair, while the paired control aliquot received an equivalent volume of H_2_O. Reactions were incubated at 37°C overnight, and deglycosylation was assessed by electrophoretic mobility shift on SDS-PAGE/immunoblot using antibodies and protocols described above

### PSCA-Bacterial Binding Assay

To assess PSCA binding to UPEC, UTI89 strain was used at a final concentration of 5×10^7^ CFU/mL in M9 minimal medium and incubated with human recombinant PSCA protein (OriGene catalog TP790100) at a final concentration of 0.05 µM for 15 minutes at 4°C, followed by five washes with M9 buffer at 5000 rpm for 5 minutes at 4°C to remove unbound protein. Samples were then centrifuged at 5000 rpm for 10 minutes at 4°C to pellet bacteria, and the supernatant was collected into a separate tube. The pellet was air-dried for 5 minutes and resuspended in M9 for downstream immunoblot analysis using antibodies and protocols described above.

### Bacterial Growth Curve Assay

UTI89 UPEC strain (10⁷ CFU/mL in M9 minimal medium) was incubated in the presence of recombinant human PSCA protein (OriGene, catalog TP790100) at 0.5 µM or PBS as a vehicle control; M9 with PBS alone served as a media-only control. Bacterial growth was monitored using a BioTek Synergy HTX multimode plate reader with a protocol consisting of continuous orbital shaking at 37°C for 12 hours, with OD600 readings collected every 10 minutes. OD600 values over time between PBS and PSCA-treated groups were compared using a generalized additive mixed model.

## Data Availability

All data produced in the present study are available upon reasonable request to the authors.

