## Supplementary Materials for "Inherited Susceptibility to Urinary Tract Infections from Kidney Papilla to Bladder"

**This PDF file includes:**

Supplementary table S1: Summary of case and control cohorts across biobanks
Supplementary table S2: Prioritized candidate genes converge on six mechanistic themes of host defense
Supplementary figures S1 to S15
Supplementary note 1: UTI case inclusion and exclusion criteria
Supplementary note 2: Description of each biobank cohort
References

**Other Supplementary Materials for this manuscript include the following:**

Supplementary data S1 to S30 (separate Excel file)

**Table S1:** Summary of case and control cohorts across biobanks with UTI phenotyping algorithm and summary statistics.

| Cohort | N Total | UTI Cases / Controls | EUR | AFR | EAS | AMR | SAS | Female (%) | Mean Age (years) |
|---|---|---|---|---|---|---|---|---|---|
| eMERGE-III | 62,283 | 9,094 / 53,189 | 7,631 / 43,265 | 1,152 / 8,525 | 311 / 1,399 | ---- | ---- | 53 | 62 |
| UK Biobank | 440,598 | 16,326 / 424,272 | 16,326 / 424,272 | ---- | ---- | ---- | ---- | 55 | 57 |
| FinnGen | 464,788 | 61,460 / 403,328 | 61,460 / 403,328 | ---- | ---- | ---- | ---- | 56 | 53 |
| BioVU | 36,665 | 5,785 / 30,880 | 5,785 / 30,880 | ---- | ---- | ---- | ---- | 60 | 34 |
| All of Us | 204,530 | 18,917 / 185,613 | 10,388 / 96,597 | 3,844 / 44,013 | 287 / 6,116 | 4,286 / 36,528 | 112 / 2,359 | 60 | 56 |
| VA Million Veteran Program | 583,494 | 67,287 / 516,207 | 46,012 / 368,445 | 15,423 / 93,003 | ---- | 5,852 / 54,759 | ---- | 9 | 62 |
| 23andMe | 68,478 | 35,000 / 33,478 | 35,000 / 33,478 | ---- | ---- | ---- | ---- | 51 | 45-60 |
| **Total** | **1,860,836** | **213,869 / 1,646,967** | **182,602 / 1,400,265** | **20,419 / 145,541** | **598 / 7,515** | **10,138 / 91,287** | **112 / 2,359** | | |

**Table S2.** Prioritized candidate genes converge on six mechanistic themes of host defense.

| Chromosome, SNP | Prioritized Gene | Functions Potentially Related to UTI Susceptibility |
|---|---|---|
| **Theme 1: GPI-Anchored Glycoprotein Defense / Mucosal Glycocalyx** | | |
| Chr8: rs2920286 | *PSCA** | • GPI-anchored glycoprotein shed into urine as heavily N-glycosylated forms (*1*)<br>• Directly binds *E. coli* and suppresses bacterial growth *in vitro* |
| Chr16: rs12917707 | *UMOD** | • GPI-anchored glycoprotein shed into urine as heavily N-glycosylated forms<br>• Constitutive urinary decoy for uropathogens, aggregating piliated bacteria (*2*) |
| Chr5: rs381949 | *CLPTM1L** | • ER processing and cell-surface trafficking of GPI-linked glycoproteins (*3*) |
| Chr19: rs507766 | *FUT2** | • α-1,2-fucosyltransferase generating ABO/H blood antigens on epithelial glycoproteins (*4*)<br>• May generate UPEC adhesin attachment sites on decoy glycoproteins by regulating fucosylation |
| **Theme 2: Innate Immune Activation** | | |
| Chr15: rs57515981 | *FES** | • Non-receptor tyrosine kinase that modulates TLR4 signaling and macrophage activation (*5*) |
| Chr1: rs4480390 | *PTPRC** | • Encodes CD45 required for lymphocyte/macrophage activation (*6*)<br>• Loss-of-function mutations cause severe combined immunodeficiency (SCID) |
| Chr1: rs4480390 | *NEK7*** | • Binds to NLRP3 to enable inflammasome assembly triggering inflammation (*7*) |
| Chr12: rs3847783 | *HOTAIR** | • LncRNA amplifying NF-κB-mediated cytokine production in macrophages upon LPS stimulation (*8*) |
| Chr18: rs7241572 | *NFATC1** | • TF with established roles in immune cell activation (*9*) |
| Chr6: rs57440165 | *BTN3A2** | • Required for activation of specific γδ T cells (*10*) |
| **Theme 3: Epithelial Cell Death and Infection Resolution** | | |
| Chr8: rs3134175 | *STK3*** | • Hippo pathway kinase with established roles in apoptosis (*11*) |
| Chr10: rs471447 | *CASP7** | • Effector caspase in apoptotic cell death (*12*) |
| Chr7: rs1111647 | *TAX1BP1*** | • Established negative regulator of NF-κB (*13*) and inhibitor of virus-induced apoptosis (*14*).<br>• Autophagy receptor facilitating clearance of bacteria such as Salmonella (*15*) and Mycobacterium (*16*) |
| Chr2: rs13402870 | *ZFP36L2** | • Anti-inflammatory mRNA-binding protein promoting cytokine mRNA decay (*17*) |
| **Theme 4: Epithelial Identity and Barrier Integrity** | | |
| Chr16: rs4786481 | *GLIS2** | • Suppresses inflammation including TLR2 signaling pathway (*18*)<br>• Biallelic mutations cause nephronophthisis (tubular atrophy/interstitial infiltration) (*19*) |
| Chr13: rs9584326 | *CLDN10** | • Kidney papilla-restricted claudin; biallelic mutations cause renal tubular dysfunction (*20*) |
| Chr16: rs1858800 | *ZFHX3** | • Differentiation-associated tumor suppressor in bladder urothelium (*21*) |
| Chr6: rs376206893 | *SPDEF** | • Drives terminal secretory epithelial differentiation and production of mucins (*22*) |
| Chr10: rs6481482 | *MPP7** | • Scaffolding protein that organizes epithelial tight junction complexes and apicobasal polarity (*23*) |
| **Theme 5: Kidney and Urinary Tract Development** | | |
| Chr10: rs10886897 | *FGFR2** | • Required for ureteric bud branching (*24*) and collecting duct patterning (*25*) |
| Chr20: rs6099264 | *BMP7** | • Sustains nephron progenitor pool (*26*), knockout leads to severe renal dysplasia (*27*) |
| Chr7: rs3919599 | *HGF** | • Promotes tubulogenesis and epithelial morphogenesis (*28*) |
| Chr7: rs9649395 | *MET** | • HGF receptor promoting tubulogenesis *in vitro* (*28*, *29*) |
| Chr7: rs6943291 | *HOXA9** | • Urogenital patterning (*30*) |
| Chr20: rs13042290 | *TSHZ2** | • By analogy to paralogue *TSHZ3*, may contribute to ureteral smooth muscle differentiation (*31*) |
| Chr22: rs7285579 | *WNT7B*** / *LINC00899** | • *WNT7B* is required for medullary collecting duct elongation (*32*)<br>• *LINC00899* is the prioritized gene for this locus with *WNT7B* as the nearest protein-coding gene |
| **Theme 6: Nutritional Immunity** | | |
| Chr12: rs35656976 | *SLC11A2** | imports ferrous iron into epithelial cells (*33*) limiting luminal iron as a mucosal defense mechanism (*34*) |
| Chr20: rs6099264 | *FAM210B*** | Mitochondrial protein required for iron import and heme synthesis during erythropoiesis (*35*) |
| Chr6: rs376206893 | *RPS10*** | Ribosomal protein linked to erythropoiesis and Diamond-Blackfan anemia (*36*) |

* Top prioritized gene for the locus; ** Second prioritized gene for the locus.

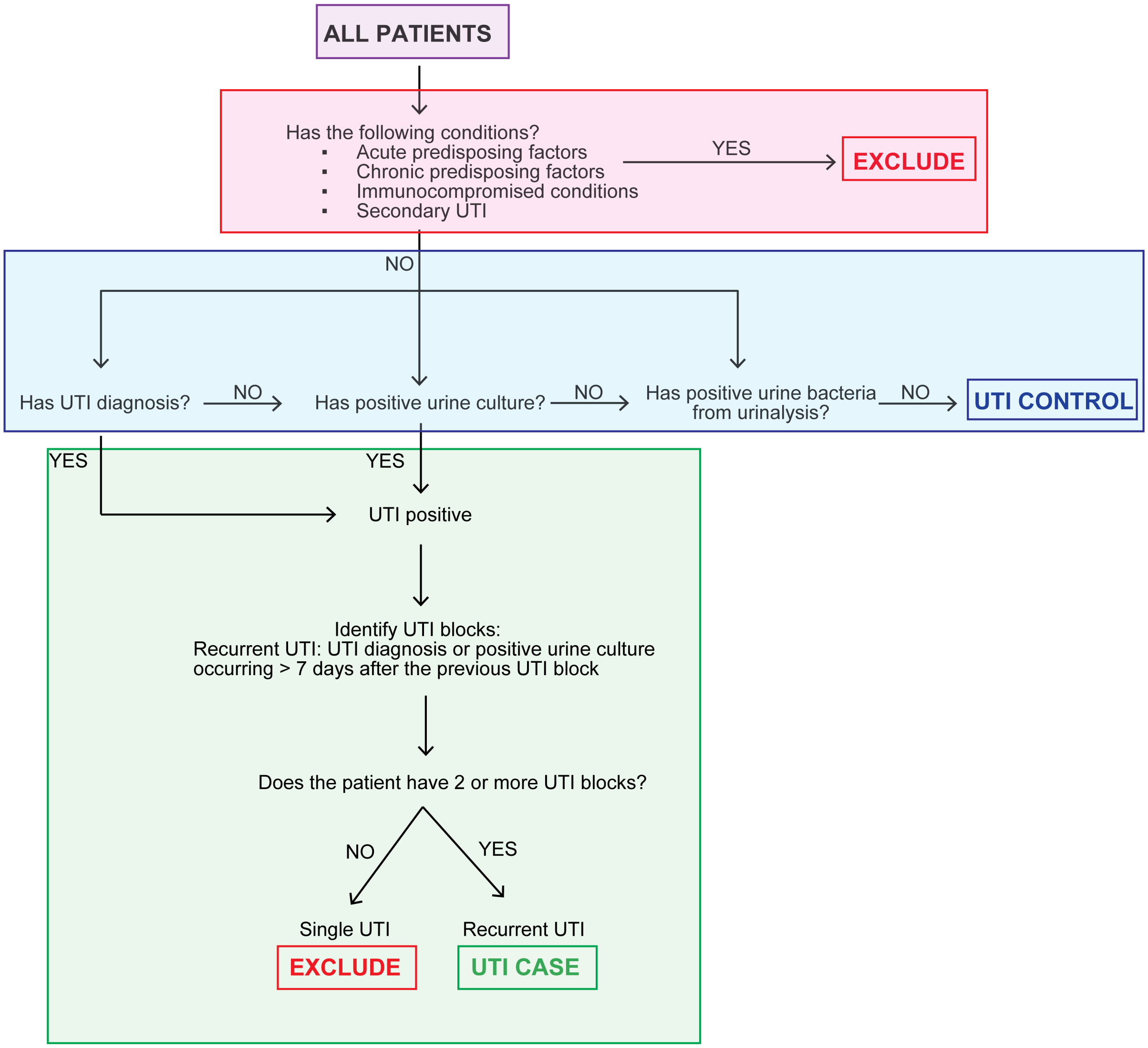


**Fig. S1. Flowchart of electronic UTI phenotyping algorithm.** All individuals with were first screened for exclusion criteria including acute and chronic predisposing factors, immunocompromised conditions, and secondary UTI (full exclusion criteria listed in Supplementary Notes). Remaining patients were classified as UTI controls if they had no UTI diagnosis, positive urine culture, or urine bacteria on urinalysis at any point in their health record history. UTI-positive patients were defined as two instances of UTI diagnosis and/or positive urine culture spaced >7 days apart. Individuals with a single occurrence of a UTI diagnosis or positive urine culture were excluded.

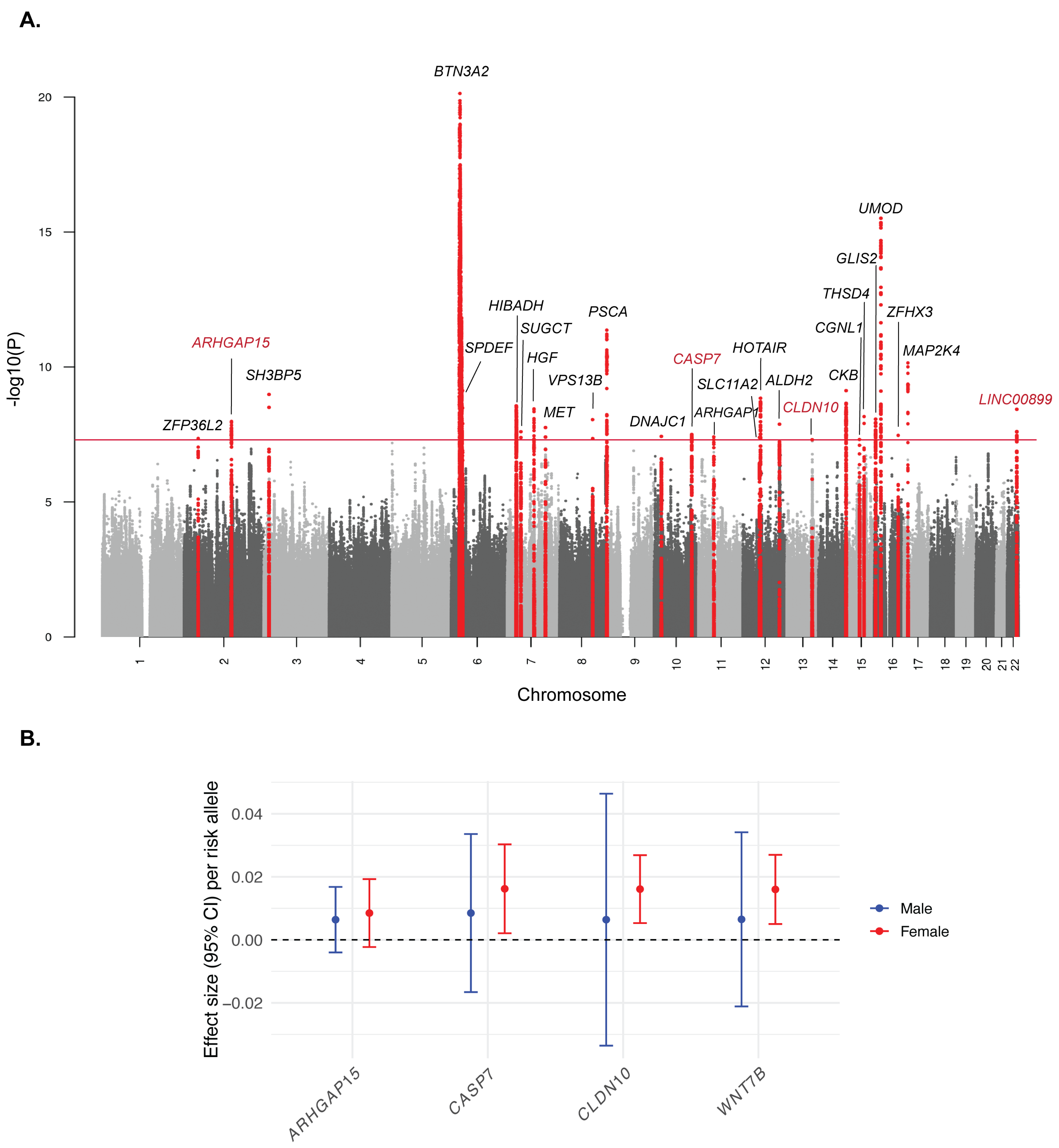


**Fig. S2. GWAS meta-analysis in European ancestry reveals genome-wide significant loci associated with UTI.** (A) Manhattan plot of GWAS meta-analysis results in individuals of European ancestry displaying -log10(P) values across all chromosomes. The red horizontal line denotes the genome-wide significance threshold ($P< 5\times10^{-8}$). Significant loci are highlighted in red, with prioritized candidate gene labels in red denoting loci not identified in the primary meta-analysis. (B) Sex-stratified effect sizes (95% CI) per risk allele for genome-wide significant loci unique to the European ancestry analysis in males (blue) and females (red).

**A.**

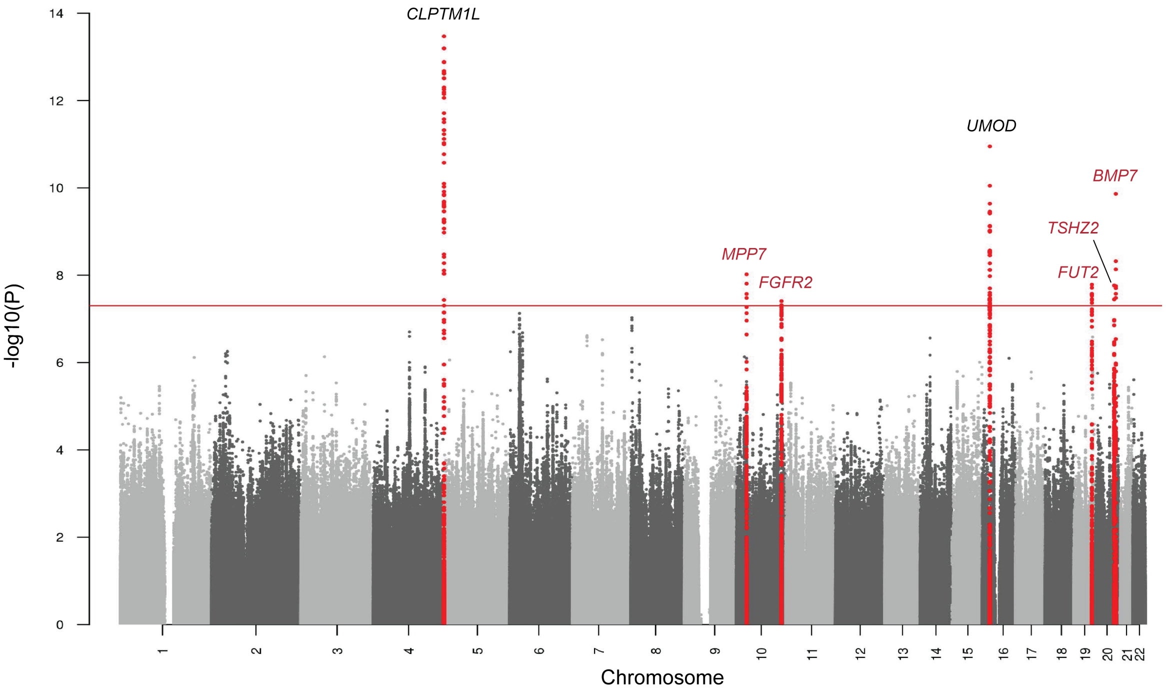


**B.**

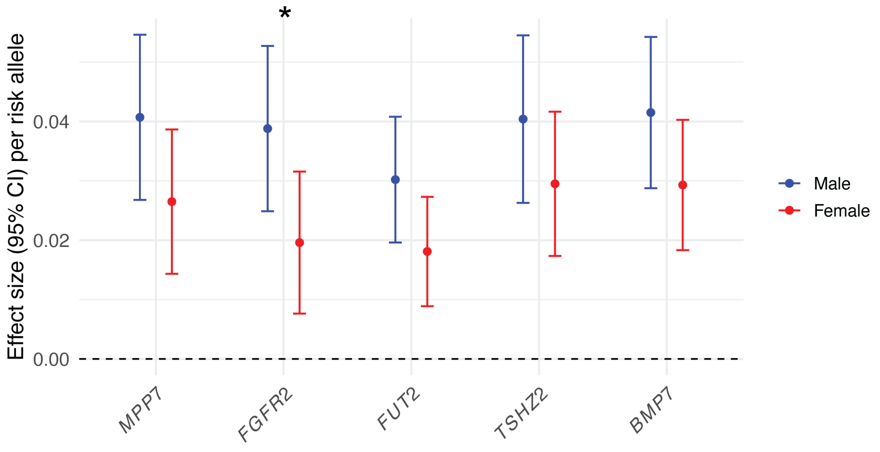


**Fig. S3. GWAS meta-analysis in males reveals genome-wide significant loci associated with UTI.** (A) Manhattan plot of GWAS meta-analysis results in males displaying -log10(P) values across all chromosomes. The red horizontal line denotes the genome-wide significance threshold ($P < 5\times10^{-8}$). Significant loci are highlighted in red, with prioritized candidate gene labels in red denoting loci not identified in the primary meta-analysis. (B) Sex-stratified effect sizes (95% CI) per risk allele for genome-wide significant loci in the male-specific analysis in males (blue) and females (red), with statistically significant Cochran's heterogeneity between sexes denoted by an asterisk (*).

**Fig. S4. Regional association plots of lead UTI GWAS loci from (A) the primary meta-analysis and (B) the secondary meta-analyses.**

**A.**

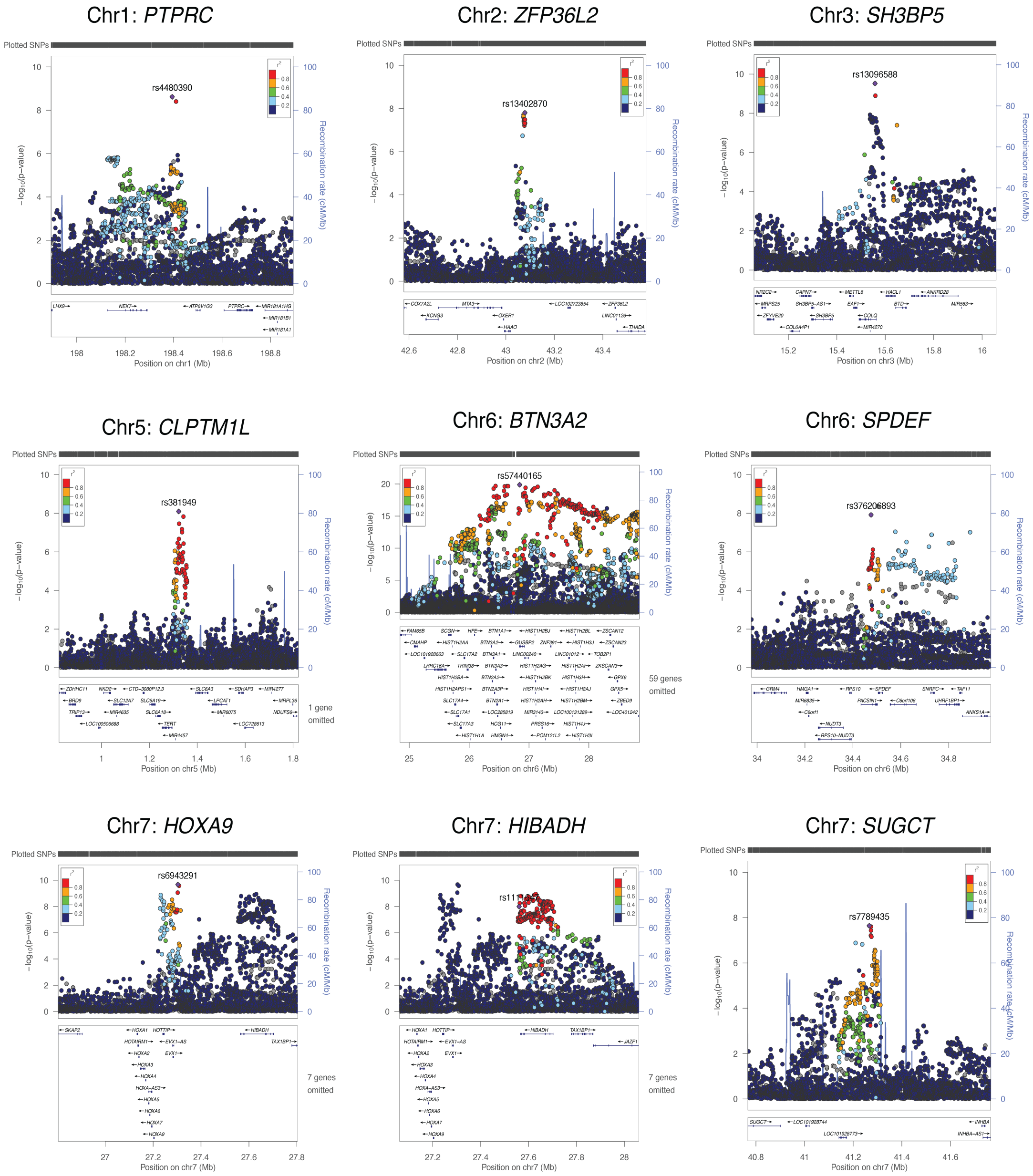

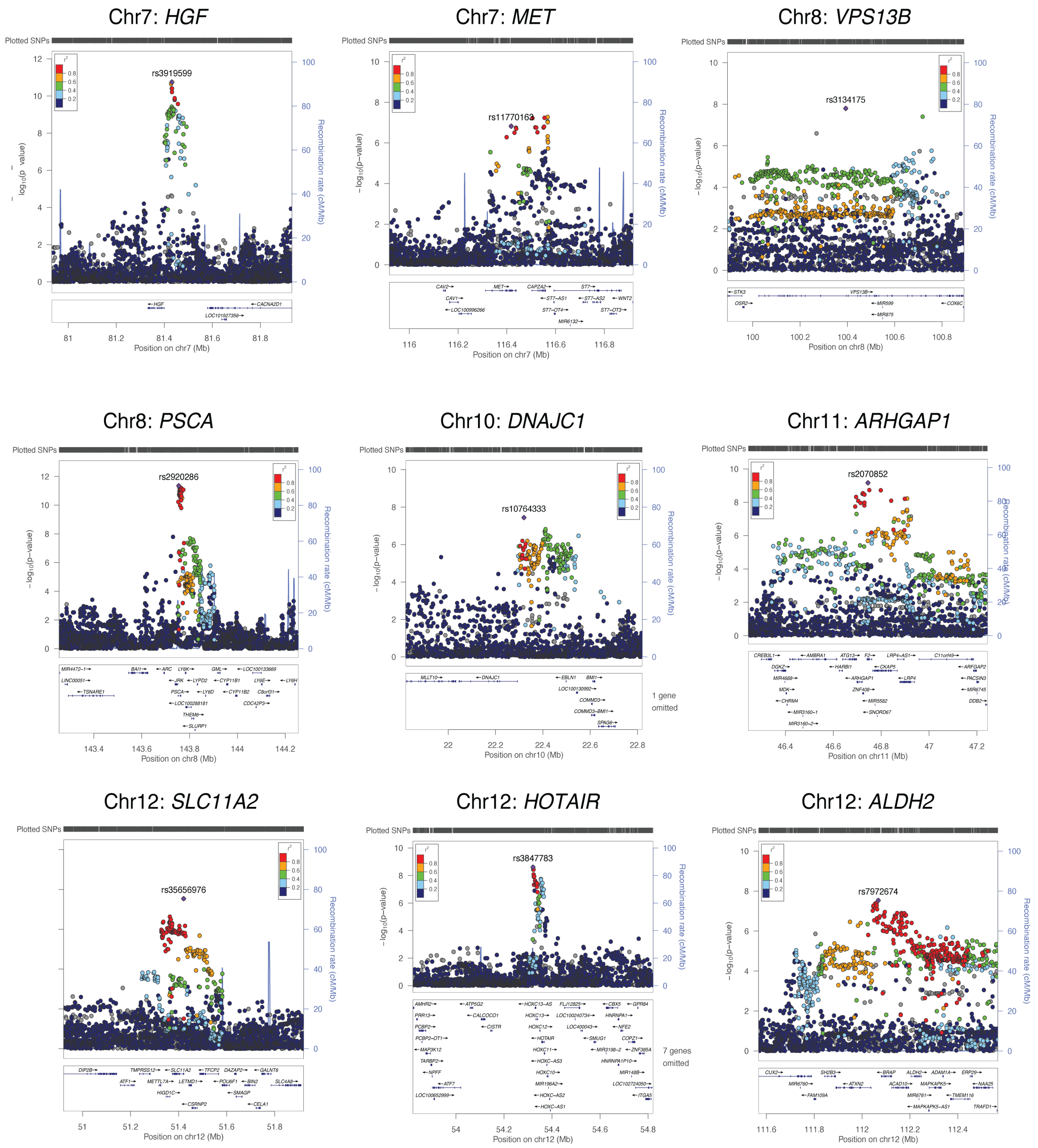
Chr7: HGF
rs3919599
Position on chr7 (Mb)
Chr7: MET
rs11770163
Position on chr7 (Mb)
Chr8: VPS13B
rs3134175
Position on chr8 (Mb)
Chr8: PSCA
rs2920286
Position on chr8 (Mb)
Chr10: DNAJC1
rs10764333
1 gene omitted
Position on chr10 (Mb)
Chr11: ARHGAP1
rs2070852
Position on chr11 (Mb)
Chr12: SLC11A2
rs35656976
Position on chr12 (Mb)
Chr12: HOTAIR
rs3847783
7 genes omitted
Position on chr12 (Mb)
Chr12: ALDH2
rs7972674
Position on chr12 (Mb)
Plotted SNPs
−log10(p−value)
Recombination rate (cM/Mb)

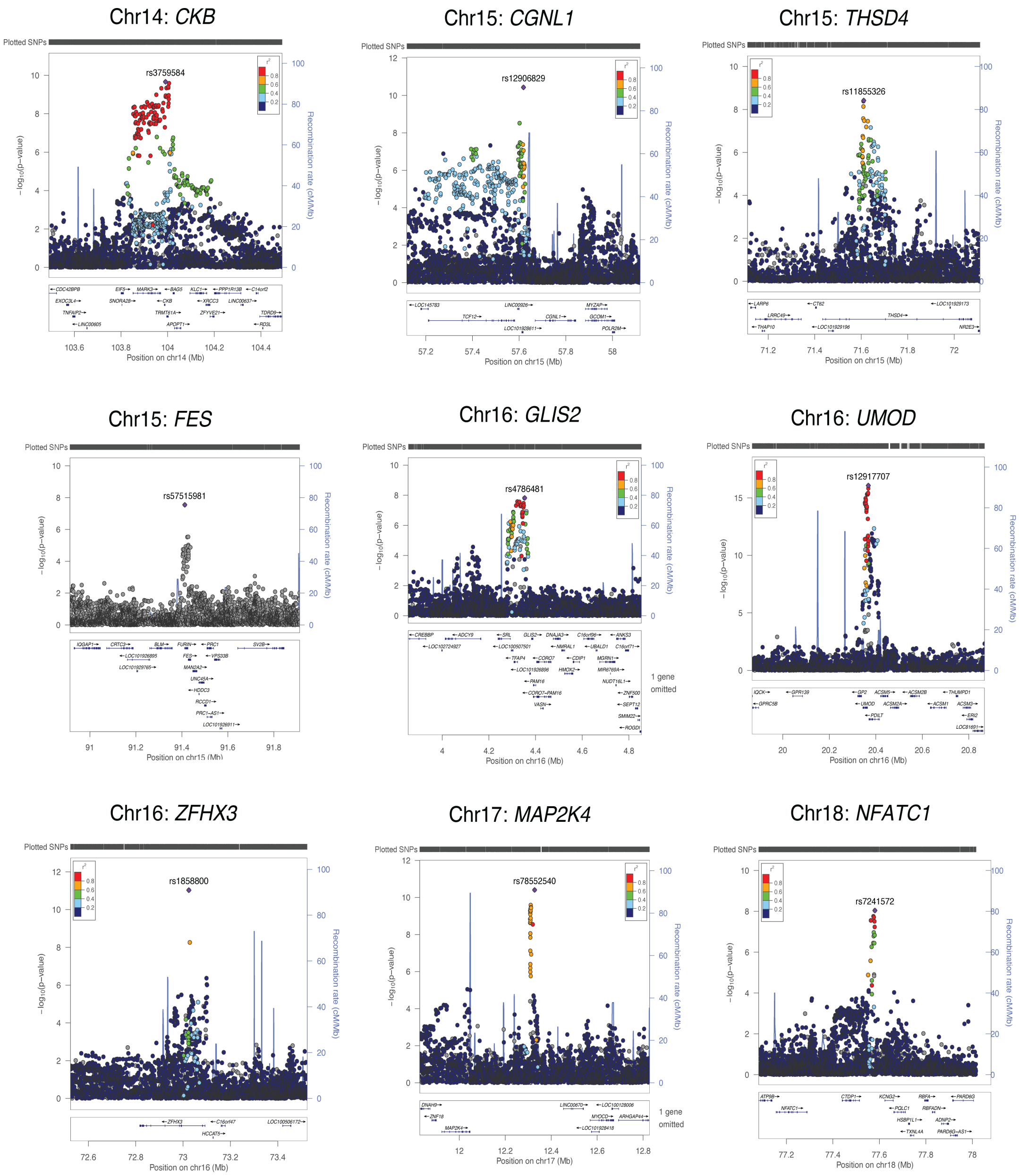

Chr14: CKB
rs3759584
Position on chr14 (Mb)
Chr15: CGNL1
rs12906829
Position on chr15 (Mb)
Chr15: THSD4
rs11855326
Position on chr15 (Mb)
Chr15: FES
rs57515981
Position on chr15 (Mb)
Chr16: GLIS2
rs4786481
Position on chr16 (Mb)
Chr16: UMOD
rs12917707
Position on chr16 (Mb)
Chr16: ZFHX3
rs1858800
Position on chr16 (Mb)
Chr17: MAP2K4
rs78552540
Position on chr17 (Mb)
Chr18: NFATC1
rs7241572
Position on chr18 (Mb)
Plotted SNPs
Recombination rate (cM/Mb)

**B.**

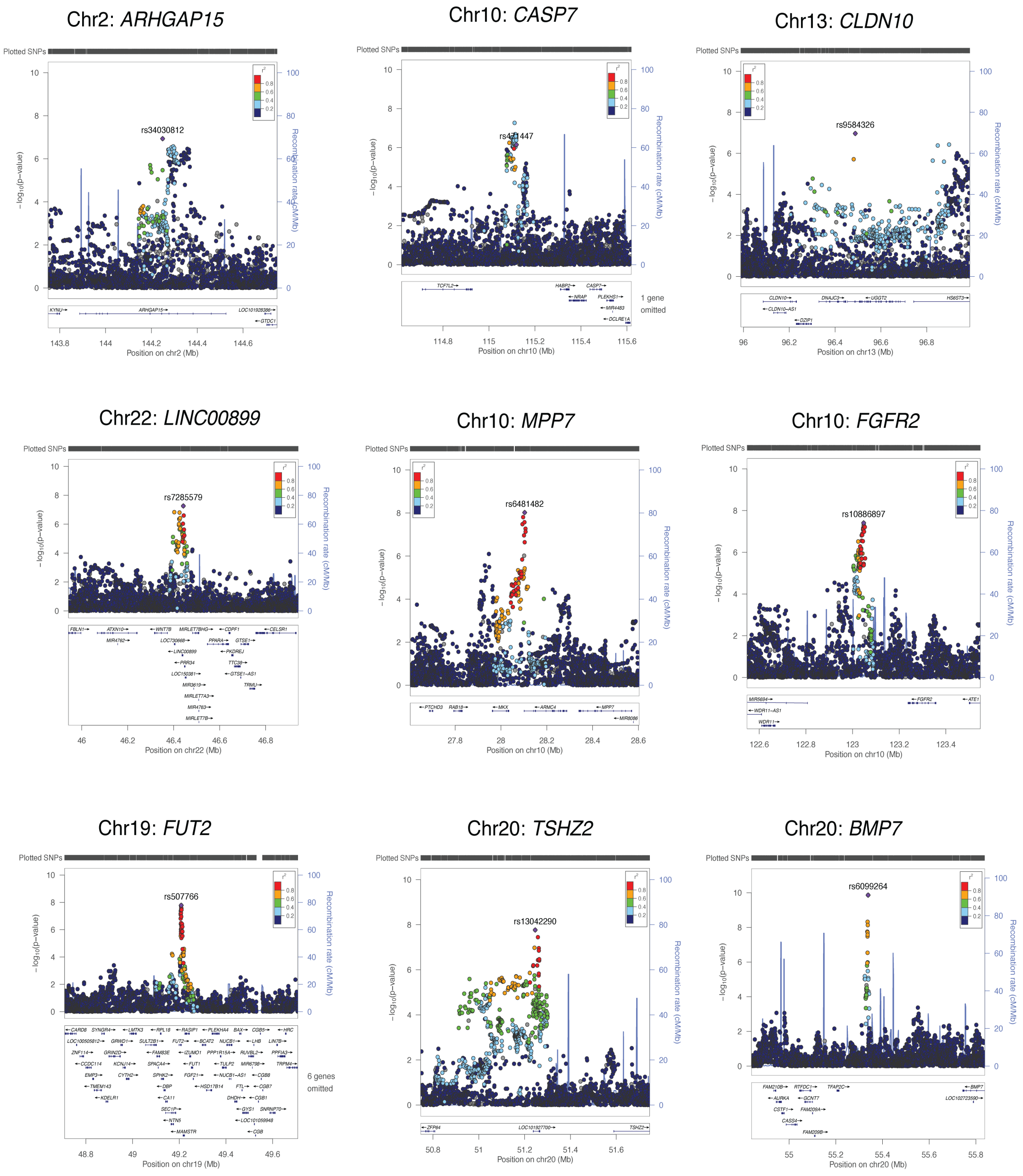


**Fig. S4. Regional association plots of lead UTI GWAS loci from (A) the primary meta-analysis and (B) the secondary meta-analyses.** Regional association plots are shown for each of the 36 genome-wide significant loci, with the genomic window defined as ±500kb of the index SNP, with the exception of the *BTN3A2* locus on chromosome 6, for which the window is expanded to ±2Mb. Each plot displays -log10(P) values (left y-axis) and recombination rate (right y-axis, blue line) across the region. Variants are colored by linkage disequilibrium ($r^2$) with the lead variant. Gene annotations are shown below each plot; where gene density is high, the number of omitted genes is indicated.

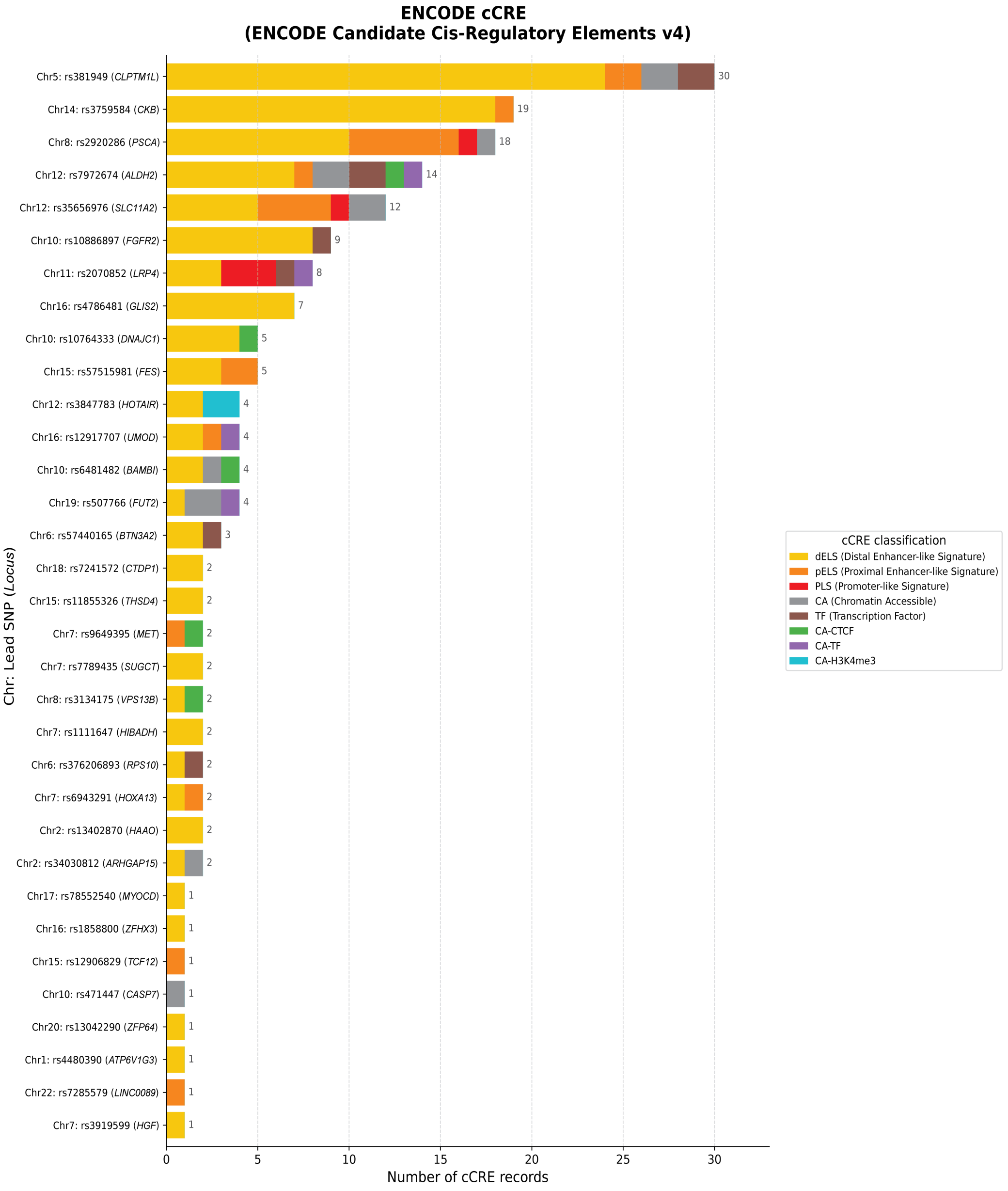


**Fig. S5. ENCODE candidate cis-regulatory element (cCRE) composition across 36 UTI GWAS loci.** A total of 174 unique cCREs overlapping variants in linkage disequilibrium ($r^2 > 0.7$) with the 36 lead SNPs were identified and classified according to ENCODE regulatory signatures (ENCODE Candidate Cis-Regulatory Elements v4). Stacked bar plots display the number of cCRE records per locus and colored by cCRE classification: distal enhancer-like signature (dELS), proximal enhancer-like signature (pELS), promoter-like signature (PLS), chromatin accessible (CA), transcription factor binding (TF), CA-CTCF (chromatin accessible-CTCF), CA-TF (chromatin accessible-transcription factor), and CA-H3K4me3 (chromatin accessible-H3K4me3).

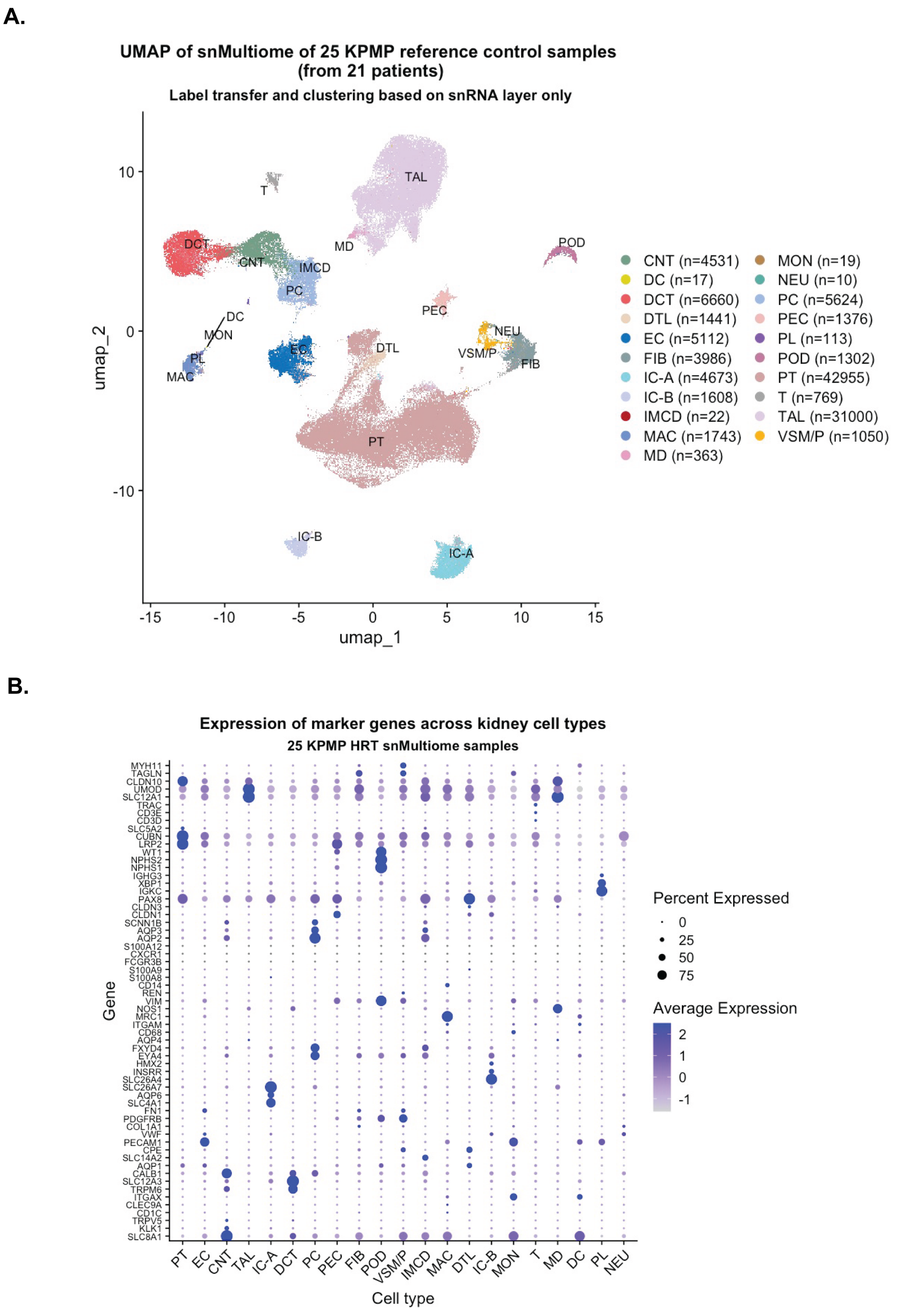


**Fig. S6. Cell-type annotation of KPMP 10x Multiome data.** (A) UMAP of 25 healthy KPMP kidney samples annotated across 21 cell types: Proximal Tubule (PT), Proximal Convoluted Tubule (PC), Thick Ascending Limb of the Loop of Henle (TAL), Distal Convoluted Tubule (DCT), Connecting Tubule (CNT), Descending Thin Limb of the Loop of Henle (DTL), Medullary Collecting Duct (MCD), Parietal Epithelial Cells (PEC), Intercalated Cell Type A (IC-A), Intercalated Cell Type B (IC-B), Podocytes (POD), Macula Densa (MD), Endothelial Cells (EC),

Vascular Smooth Muscle/Pericytes (VSM/P), Fibroblasts (FIB), T Cells (T), Monocytes (MON), Macrophages (MAC), Dendritic Cells (DC), Neurons (NEU), and Plasma Cells (PL). Cell-type annotations were assigned via label transfer using KPMP Atlas V2.0 snRNA-seq as reference. (B) Dot plot showing expression of canonical marker genes used to validate each cell cluster.

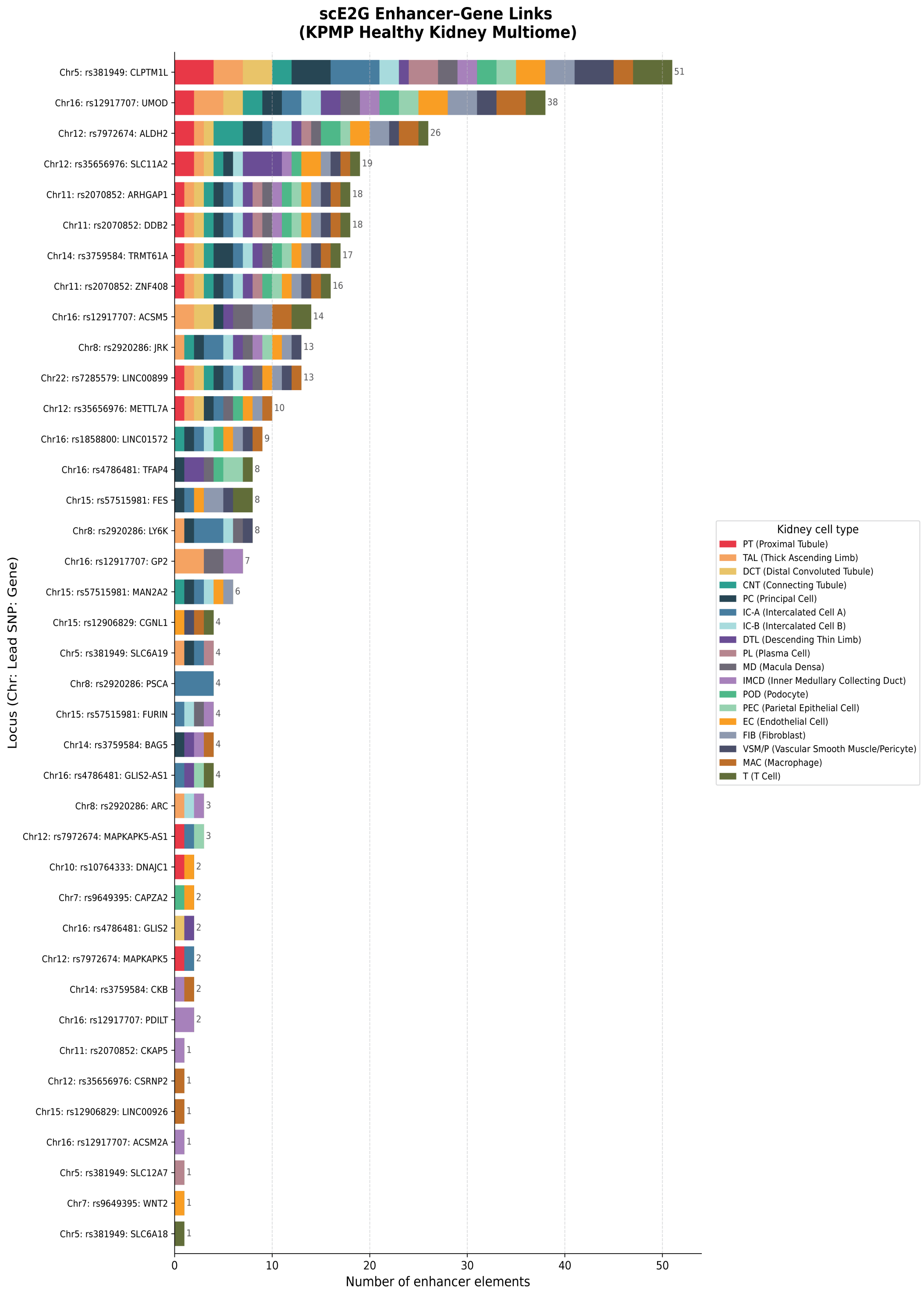


**Fig. S7. scE2G single cell enhancer-gene links per GWAS locus across KPMP healthy kidney cell types.** Fourteen UTI lead SNPs mapped to 39 genes based on 348 enhancer-gene links spanning 18 kidney cell types.

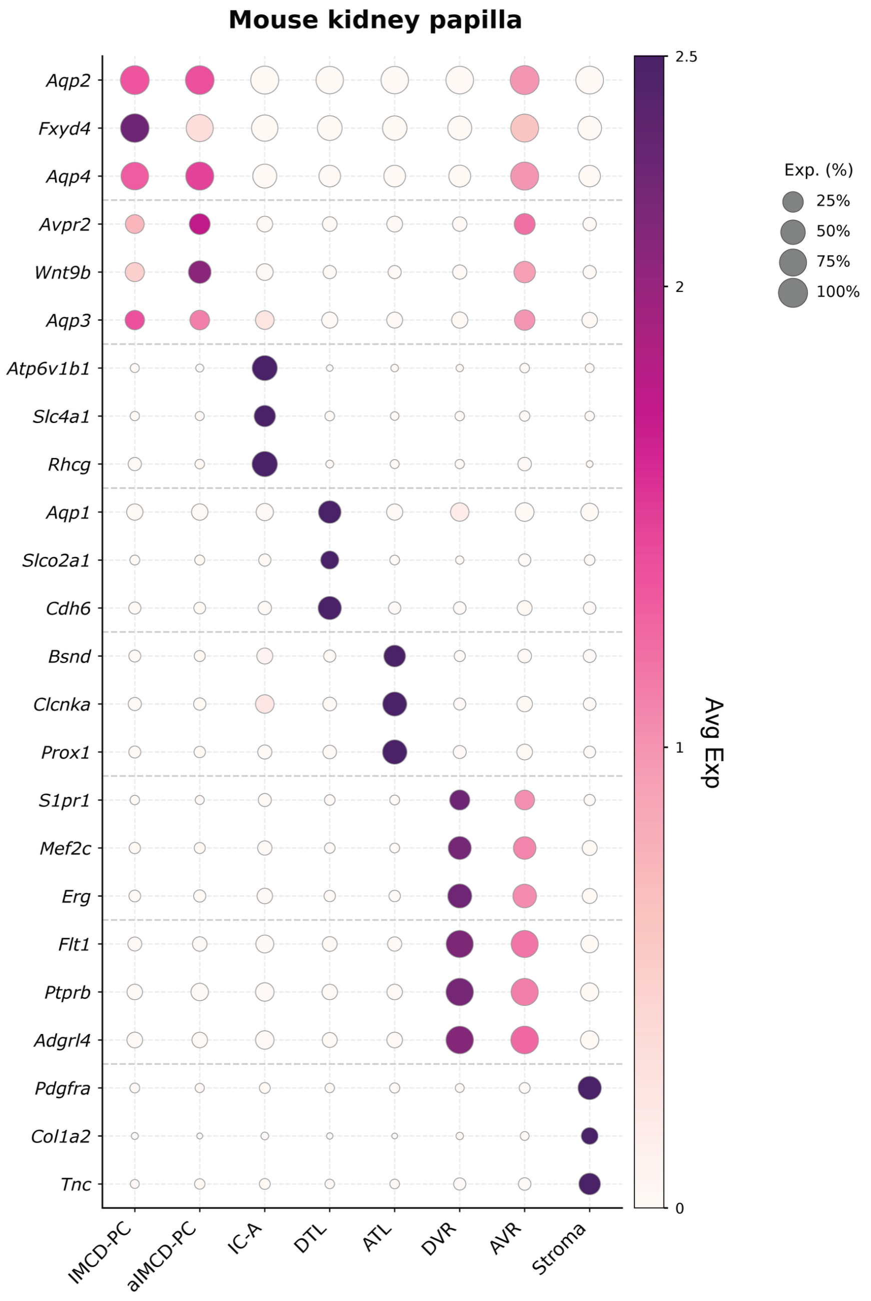


**Fig. S8. Single-cell gene expression across mouse kidney papilla cell types**. Expression of marker genes across 8 kidney papilla cell types in mouse. Dot size represents the percentage of cells expressing each gene (Exp. (%): 25%, 50%, 75%, 100%) and dot color indicates z-score normalized average expression per gene across cell types (Avg Exp), highlighting cell type-specific enrichment of each marker. Cell type abbreviations: IMCD-PC (Inner Medullary Collecting Duct - Principal Cell), aIMCD-PC (adaptive Inner Medullary Collecting Duct - Principal Cell), IC-A (Intercalated Cell Type A), DTL (Descending Thin Limb), ATL (Ascending Thin Limb), DVR (Descending Vasa Recta), AVR (Ascending Vasa Recta), Stroma.

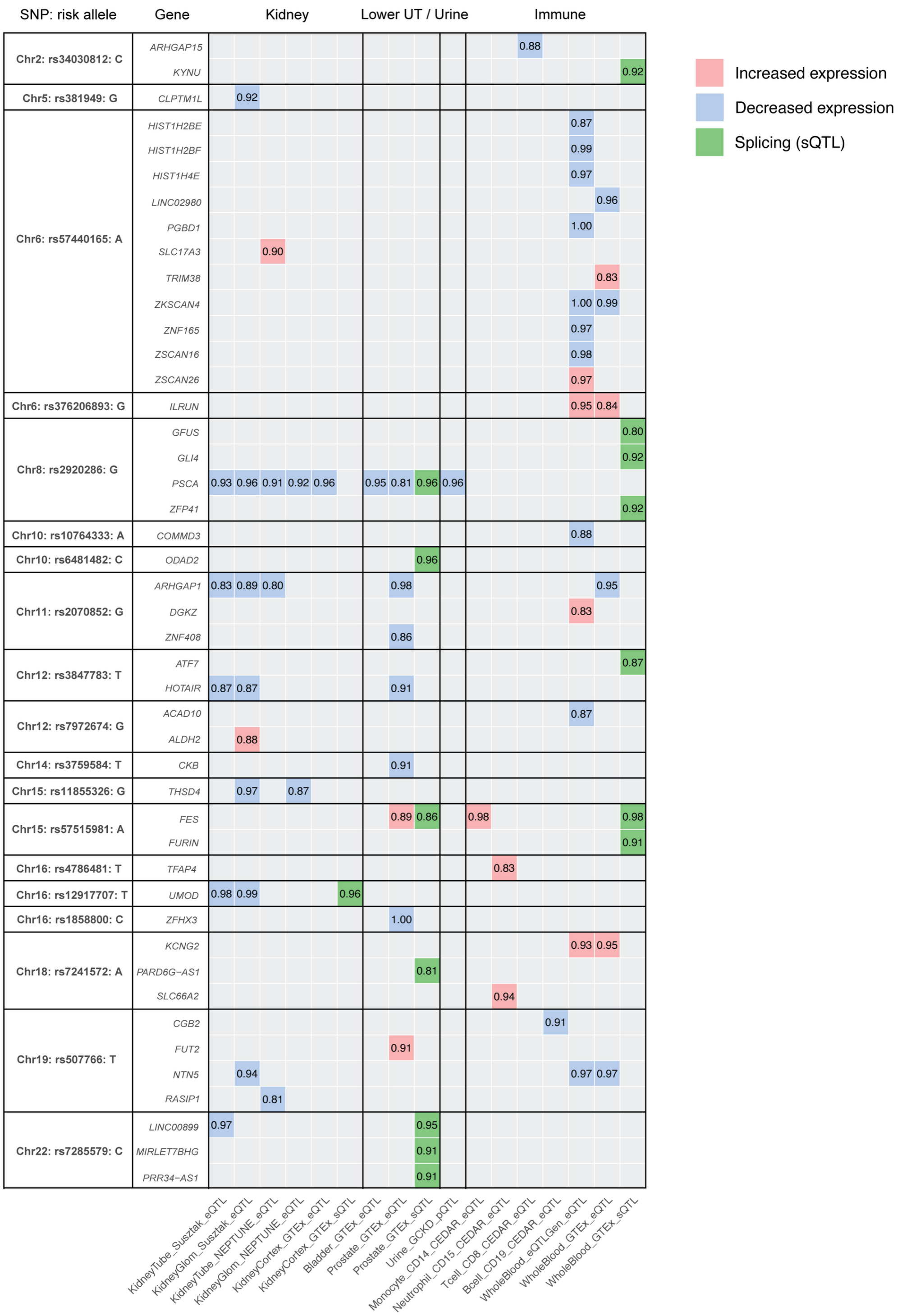


**Fig. S9. Colocalization of UTI GWAS and tissue-specific QTL signals.** Posterior probabilities of colocalization (PPH4) between UTI GWAS signals and eQTL, sQTL, or pQTL signals across kidney, lower urinary tract (bladder, prostate), urine, immune cells, and whole blood datasets for each lead GWAS variant and candidate gene (within ±500kb of the lead SNP) are represented in the heatmap. Only locus-gene pairs with PPH4 ≥ 0.80 are shown. Cell color indicates the direction of effect of the risk allele on gene expression: pink (increased expression), blue (decreased expression), and green (splicing, sQTL).

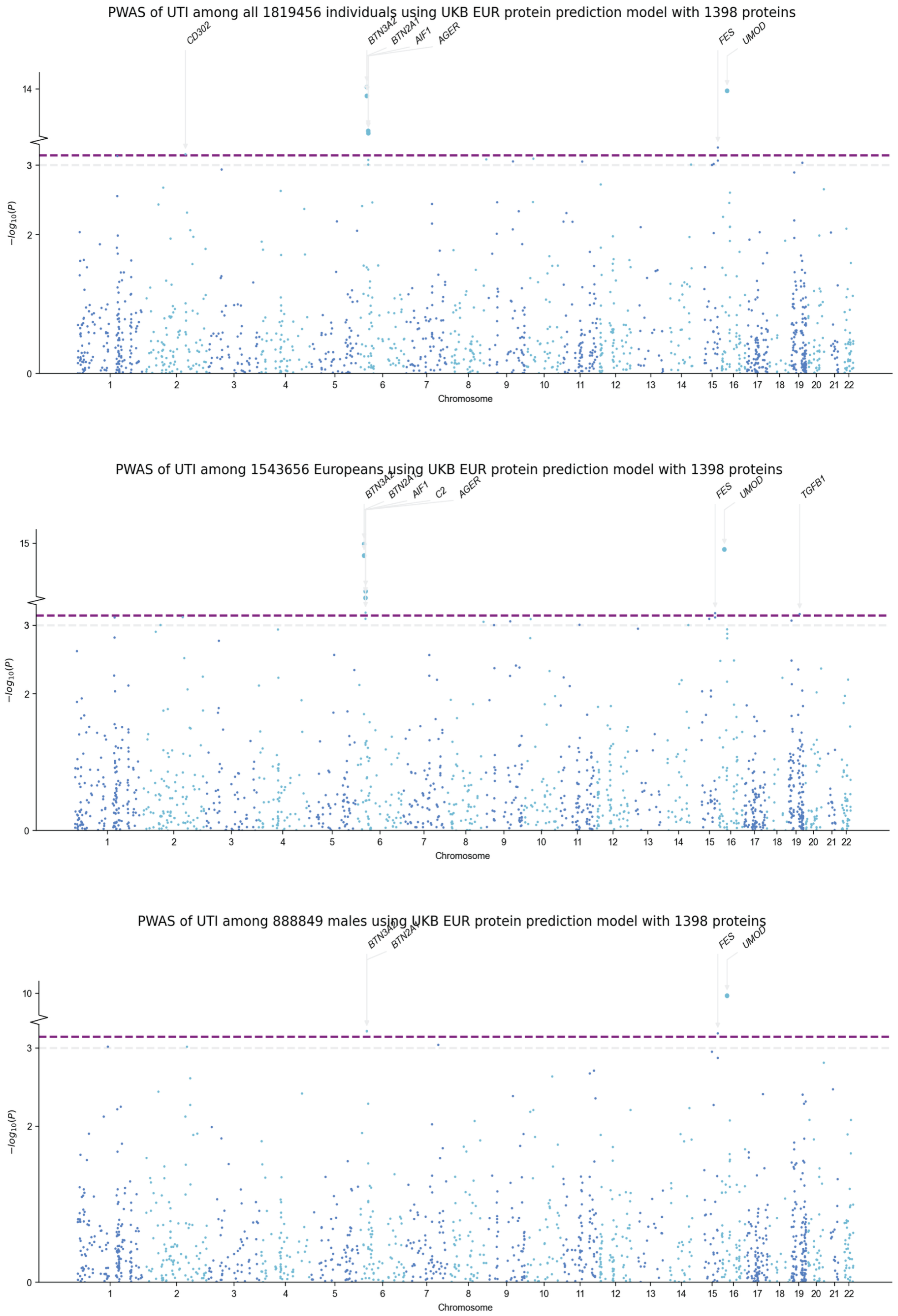


**Fig. S10. UKBB-BLISS ancestry-specific blood PWAS model for UTI risk loci.** Manhattan plots of proteome-wide association study (PWAS) results for UTI using the UKBB European (EUR) ancestry protein prediction model with 1,398 proteins, shown for our primary meta-analysis: all individuals (n=1,819,456, top), and our secondary analyses: European-ancestry (n=1,543,656, middle), and Males (n=888,849, bottom). The dashed purple horizontal line denotes the proteome-wide significance threshold. Significant proteins are labeled above each peak.

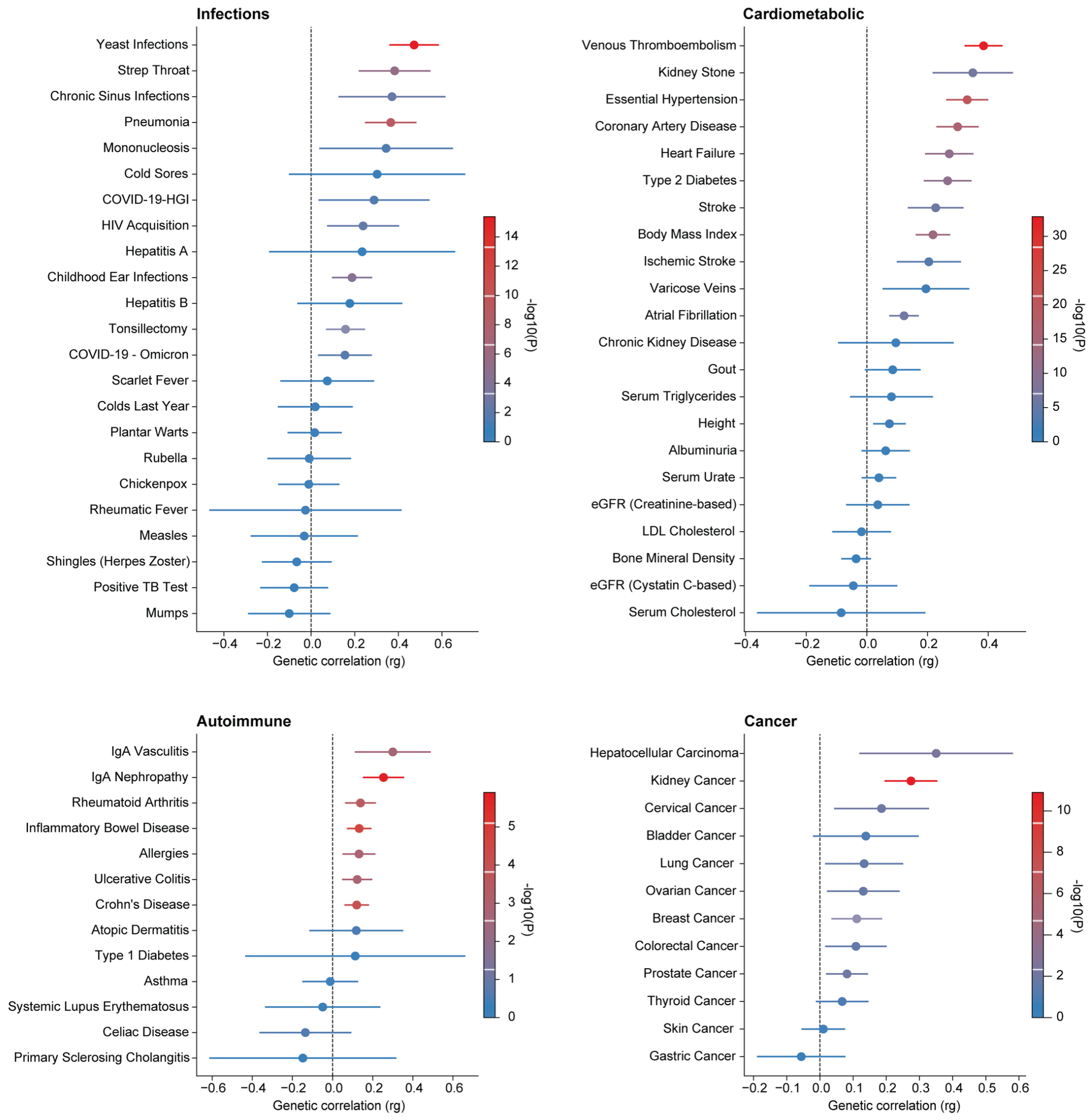


**Fig. S11. Genetic correlation of UTI GWAS meta-analysis with other GWAS traits excluding the HLA region.** (A) Genetic correlations between UTI and complex traits across four disease domains, excluding the HLA region. Forest plots show genetic correlations (*rg*) between UTI and (A) infections, (B) cardiovascular, renal, and metabolic traits, (C) autoimmune and inflammatory traits, and (D) malignancies, estimated by bivariate LD score regression (LDSC). Points represent rg point estimates; horizontal lines represent 95% confidence intervals. The dashed vertical line marks rg=0. Point color reflects statistical significance (−log10 P-value; see legend, scaled independently within each panel). Full summary statistics, including standard errors and exact P-values, are provided in Table S29.

**Fig. S12. Meta PheWAS plots of each UTI GWAS lead SNP.**

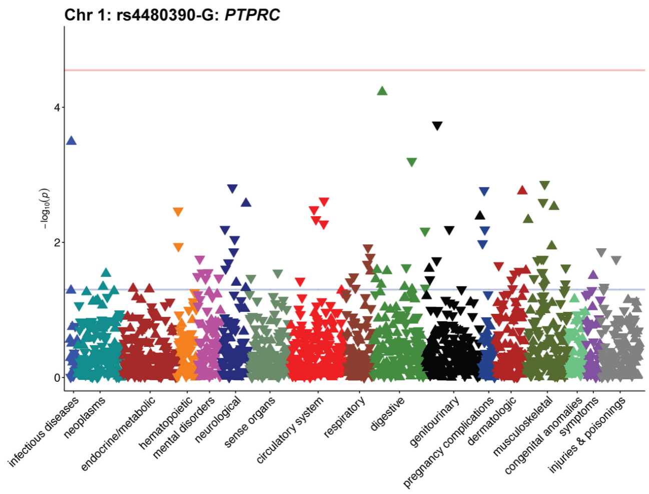


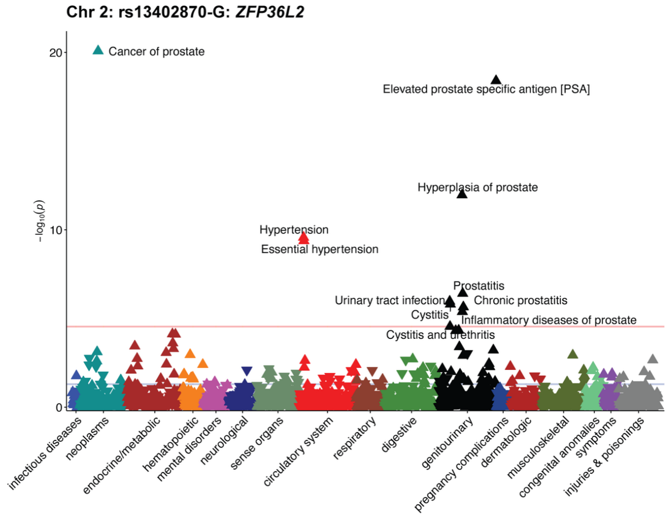


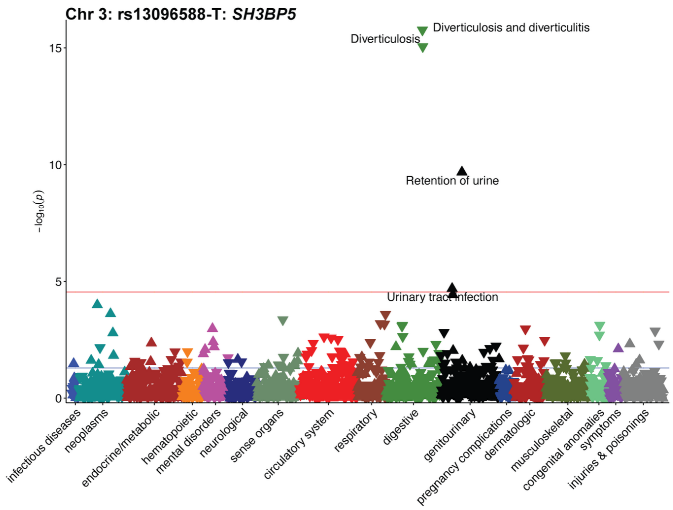

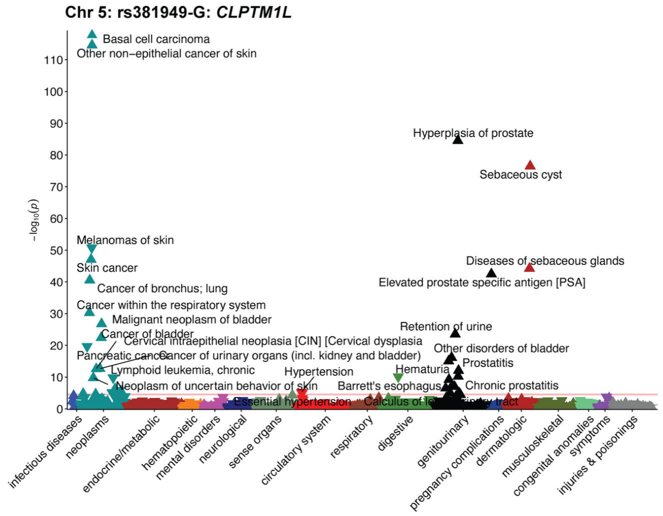
Chr 5: rs381949-G: CLPTM1L
−log10(p)
Basal cell carcinoma
Other non−epithelial cancer of skin
Hyperplasia of prostate
Sebaceous cyst
Melanomas of skin
Skin cancer
Diseases of sebaceous glands
Elevated prostate specific antigen [PSA]
Cancer of bronchus; lung
Cancer within the respiratory system
Malignant neoplasm of bladder
Cancer of bladder
Retention of urine
Cervical intraepithelial neoplasia [CIN] [Cervical dysplasia
Other disorders of bladder
Cancer of urinary organs (incl. kidney and bladder)
Prostatitis
Lymphoid leukemia, chronic
Hypertension
Hematuria
Neoplasm of uncertain behavior of skin
Barrett's esophagus
Chronic prostatitis
Essential hypertension
infectious diseases
neoplasms
endocrine/metabolic
hematopoietic
mental disorders
neurological
sense organs
circulatory system
respiratory
digestive
genitourinary
pregnancy complications
dermatologic
musculoskeletal
congenital anomalies
symptoms
injuries & poisonings

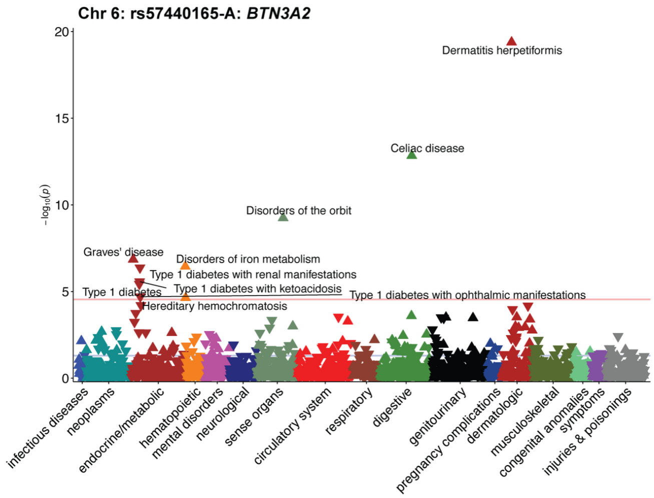
Chr 6: rs57440165-A: BTN3A2
−log10(p)
Dermatitis herpetiformis
Celiac disease
Disorders of the orbit
Graves' disease
Disorders of iron metabolism
Type 1 diabetes with renal manifestations
Type 1 diabetes with ketoacidosis
Type 1 diabetes
Type 1 diabetes with ophthalmic manifestations
Hereditary hemochromatosis
infectious diseases
neoplasms
endocrine/metabolic
hematopoietic
mental disorders
neurological
sense organs
circulatory system
respiratory
digestive
genitourinary
pregnancy complications
dermatologic
musculoskeletal
congenital anomalies
symptoms
injuries & poisonings

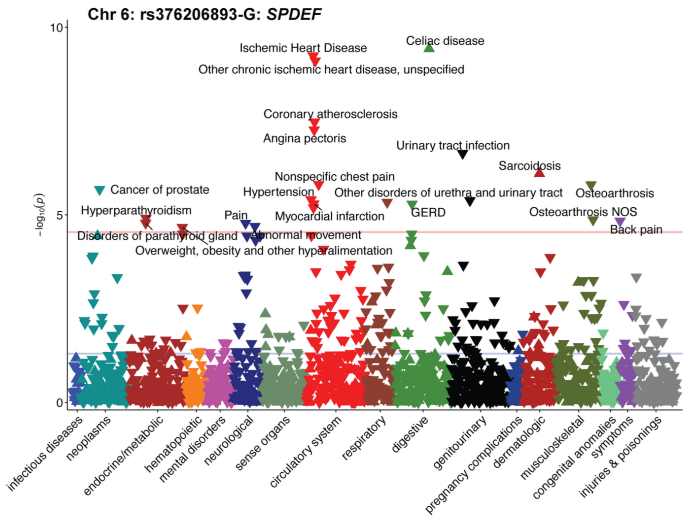
Chr 6: rs376206893-G: SPDEF
−log10(p)
Celiac disease
Ischemic Heart Disease
Other chronic ischemic heart disease, unspecified
Coronary atherosclerosis
Angina pectoris
Urinary tract infection
Sarcoidosis
Nonspecific chest pain
Cancer of prostate
Hypertension
Other disorders of urethra and urinary tract
Osteoarthrosis
Hyperparathyroidism
Pain
Myocardial infarction
GERD
Osteoarthrosis NOS
Back pain
Disorders of parathyroid gland
Abnormal movement
Overweight, obesity and other hyperalimentation
infectious diseases
neoplasms
endocrine/metabolic
hematopoietic
mental disorders
neurological
sense organs
circulatory system
respiratory
digestive
genitourinary
pregnancy complications
dermatologic
musculoskeletal
congenital anomalies
symptoms
injuries & poisonings

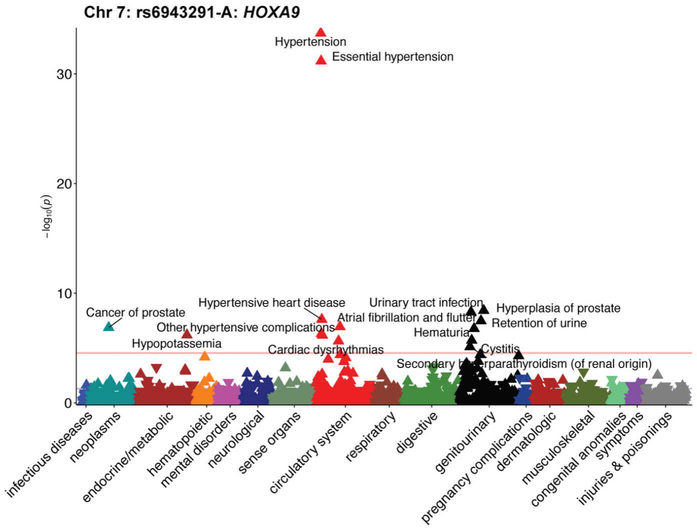
Chr 7: rs6943291-A: *HOXA9*
Hypertension
Essential hypertension
Cancer of prostate
Hypertensive heart disease
Other hypertensive complications
Hypopotassemia
Cardiac dysrhythmias
Atrial fibrillation and flutter
Urinary tract infection
Hematuria
Hyperplasia of prostate
Retention of urine
Cystitis
Secondary hyperparathyroidism (of renal origin)
$-\log_{10}(p)$
infectious diseases
neoplasms
endocrine/metabolic
hematopoietic
mental disorders
neurological
sense organs
circulatory system
respiratory
digestive
genitourinary
pregnancy complications
dermatologic
musculoskeletal
congenital anomalies
symptoms
injuries & poisonings

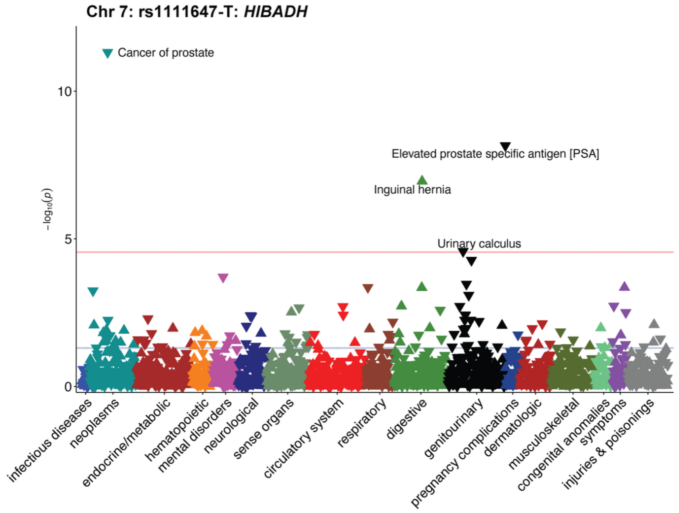
Chr 7: rs1111647-T: *HIBADH*
Cancer of prostate
Elevated prostate specific antigen [PSA]
Inguinal hernia
Urinary calculus
$-\log_{10}(p)$
infectious diseases
neoplasms
endocrine/metabolic
hematopoietic
mental disorders
neurological
sense organs
circulatory system
respiratory
digestive
genitourinary
pregnancy complications
dermatologic
musculoskeletal
congenital anomalies
symptoms
injuries & poisonings

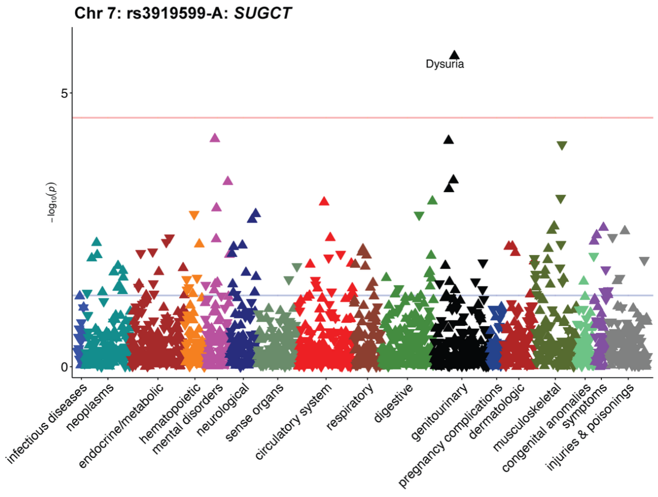
Chr 7: rs3919599-A: *SUGCT*
Dysuria
$-\log_{10}(p)$
infectious diseases
neoplasms
endocrine/metabolic
hematopoietic
mental disorders
neurological
sense organs
circulatory system
respiratory
digestive
genitourinary
pregnancy complications
dermatologic
musculoskeletal
congenital anomalies
symptoms
injuries & poisonings

**Chr 7: rs3919599-C: *HGF***

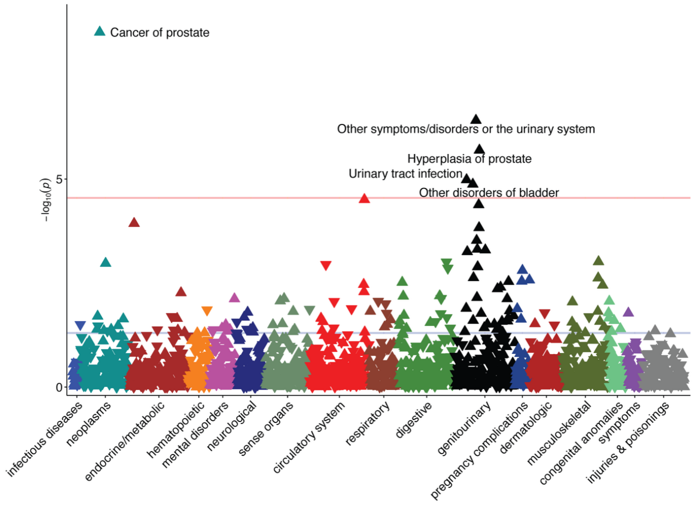


**Chr 7: rs9649395-A: *MET***

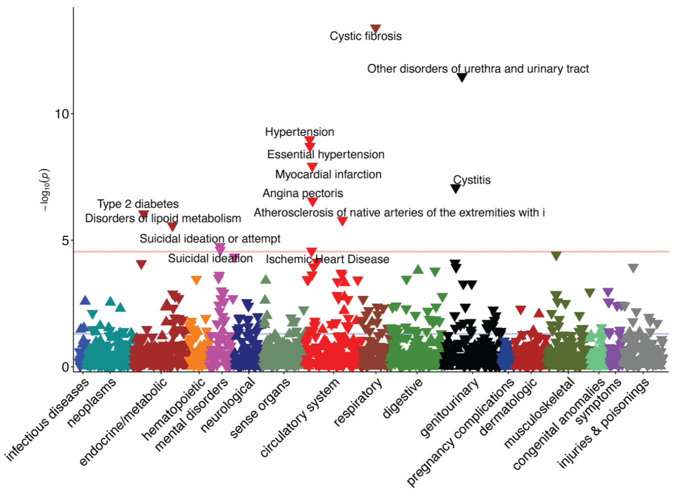


**Chr 8: rs3134175-T: *VPS13B***

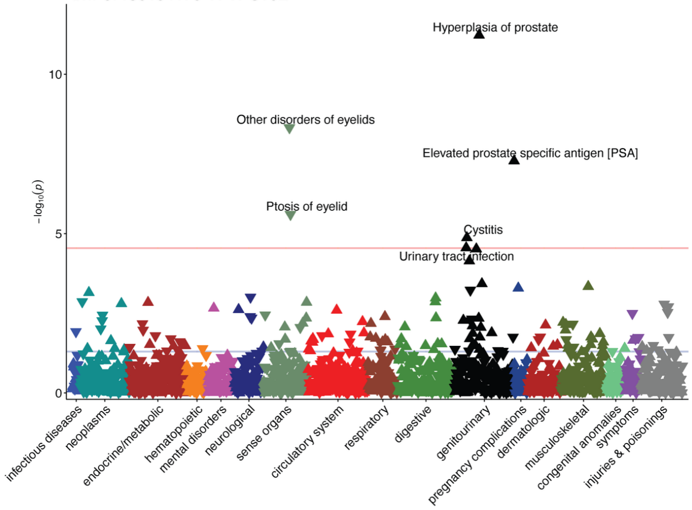

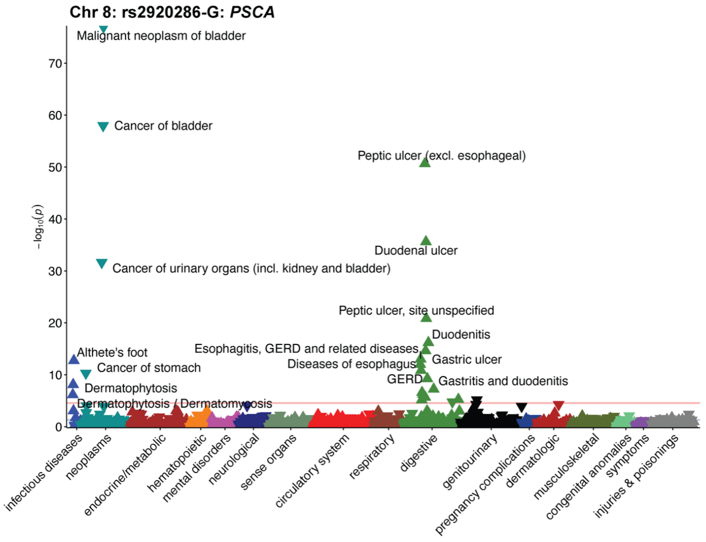
Chr 8: rs2920286-G: *PSCA*
Malignant neoplasm of bladder
Cancer of bladder
Peptic ulcer (excl. esophageal)
Duodenal ulcer
Cancer of urinary organs (incl. kidney and bladder)
Peptic ulcer, site unspecified
Duodenitis
Esophagitis, GERD and related diseases
Diseases of esophagus
Gastric ulcer
GERD
Gastritis and duodenitis
Althete's foot
Cancer of stomach
Dermatophytosis
Dermatophytosis / Dermatomycosis
−log10(p)
0
10
20
30
40
50
60
70
infectious diseases
neoplasms
endocrine/metabolic
hematopoietic
mental disorders
neurological
sense organs
circulatory system
respiratory
digestive
genitourinary
pregnancy complications
dermatologic
musculoskeletal
congenital anomalies
symptoms
injuries & poisonings

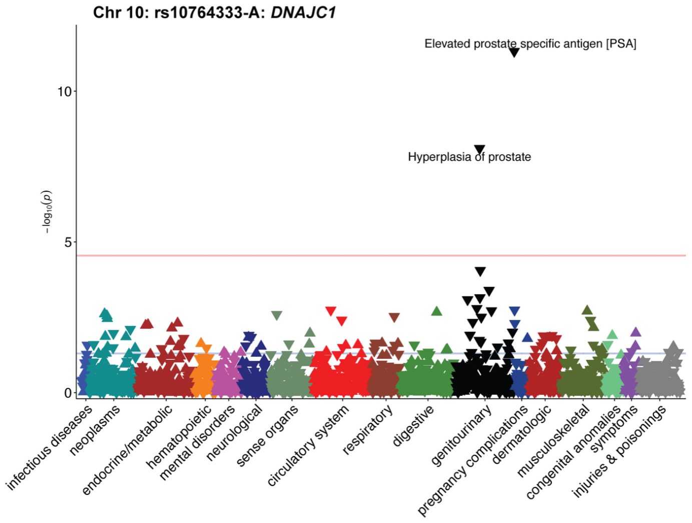
Chr 10: rs10764333-A: *DNAJC1*
Elevated prostate specific antigen [PSA]
Hyperplasia of prostate
−log10(p)
0
5
10
infectious diseases
neoplasms
endocrine/metabolic
hematopoietic
mental disorders
neurological
sense organs
circulatory system
respiratory
digestive
genitourinary
pregnancy complications
dermatologic
musculoskeletal
congenital anomalies
symptoms
injuries & poisonings

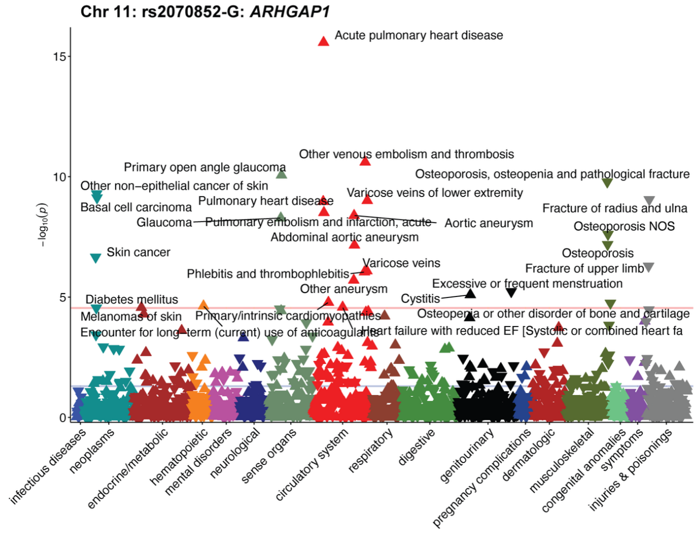
Chr 11: rs2070852-G: *ARHGAP1*
Acute pulmonary heart disease
Other venous embolism and thrombosis
Primary open angle glaucoma
Osteoporosis, osteopenia and pathological fracture
Other non−epithelial cancer of skin
Varicose veins of lower extremity
Basal cell carcinoma
Pulmonary heart disease
Fracture of radius and ulna
Glaucoma
Pulmonary embolism and infarction, acute
Aortic aneurysm
Osteoporosis NOS
Abdominal aortic aneurysm
Skin cancer
Osteoporosis
Varicose veins
Fracture of upper limb
Phlebitis and thrombophlebitis
Excessive or frequent menstruation
Other aneurysm
Diabetes mellitus
Cystitis
Melanomas of skin
Primary/intrinsic cardiomyopathies
Osteopenia or other disorder of bone and cartilage
Encounter for long−term (current) use of anticoagulants
Heart failure with reduced EF [Systolic or combined heart fa
−log10(p)
0
5
10
15
infectious diseases
neoplasms
endocrine/metabolic
hematopoietic
mental disorders
neurological
sense organs
circulatory system
respiratory
digestive
genitourinary
pregnancy complications
dermatologic
musculoskeletal
congenital anomalies
symptoms
injuries & poisonings

**Chr 12: rs35656976-T: *SLC11A2***

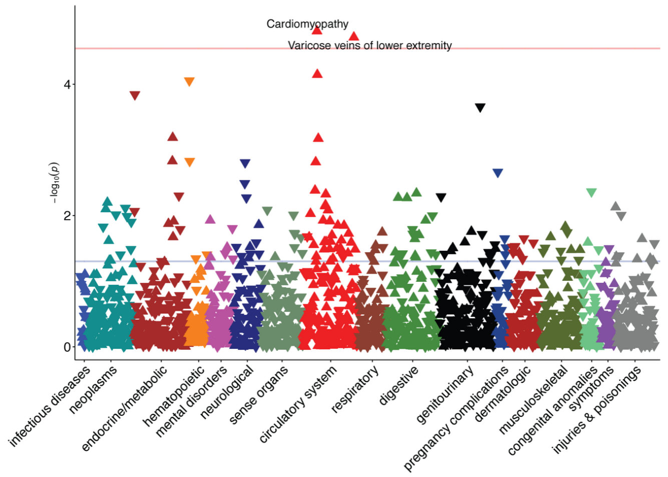


**Chr 12: rs3847783-T: *HOTAIR***

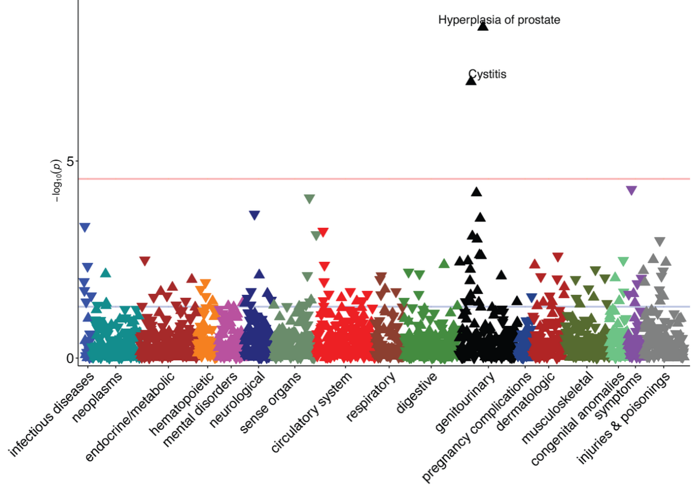


**Chr 12: rs7972674-G: *ALDH2***

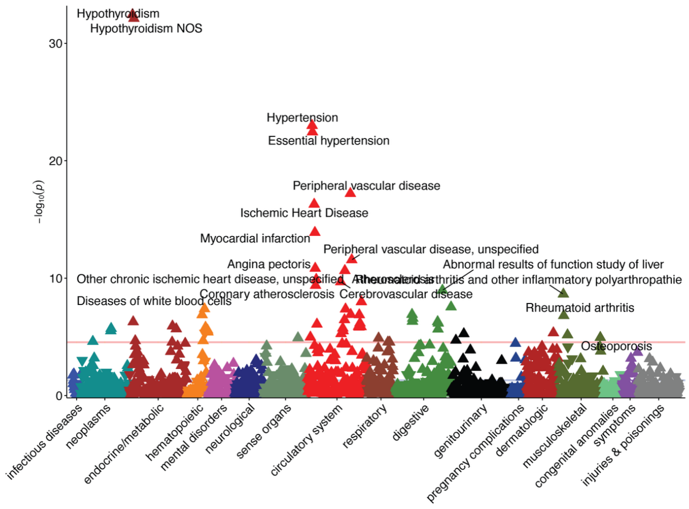

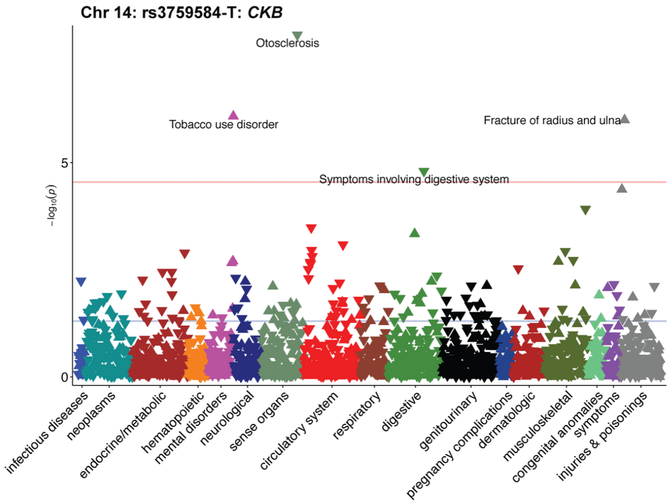

Chr 14: rs3759584-T: CKB
Otosclerosis
Tobacco use disorder
Fracture of radius and ulna
Symptoms involving digestive system
−log10(p)
0
5
infectious diseases
neoplasms
endocrine/metabolic
hematopoietic
mental disorders
neurological
sense organs
circulatory system
respiratory
digestive
genitourinary
pregnancy complications
dermatologic
musculoskeletal
congenital anomalies
symptoms
injuries & poisonings


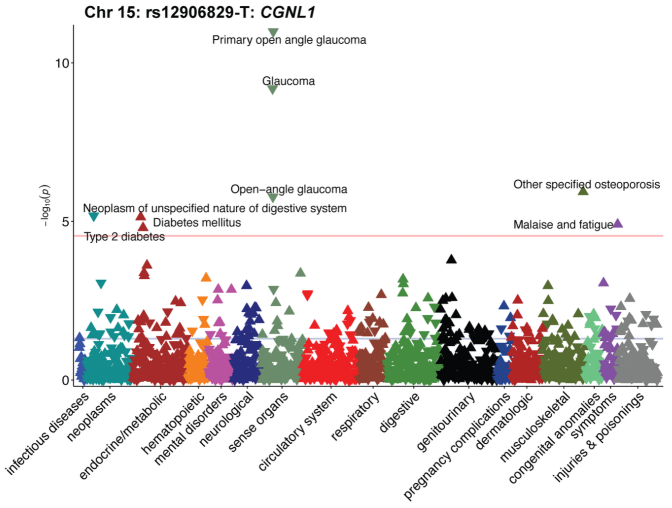

Chr 15: rs12906829-T: CGNL1
Primary open angle glaucoma
Glaucoma
Open−angle glaucoma
Other specified osteoporosis
Neoplasm of unspecified nature of digestive system
Diabetes mellitus
Type 2 diabetes
Malaise and fatigue
−log10(p)
0
5
10
infectious diseases
neoplasms
endocrine/metabolic
hematopoietic
mental disorders
neurological
sense organs
circulatory system
respiratory
digestive
genitourinary
pregnancy complications
dermatologic
musculoskeletal
congenital anomalies
symptoms
injuries & poisonings


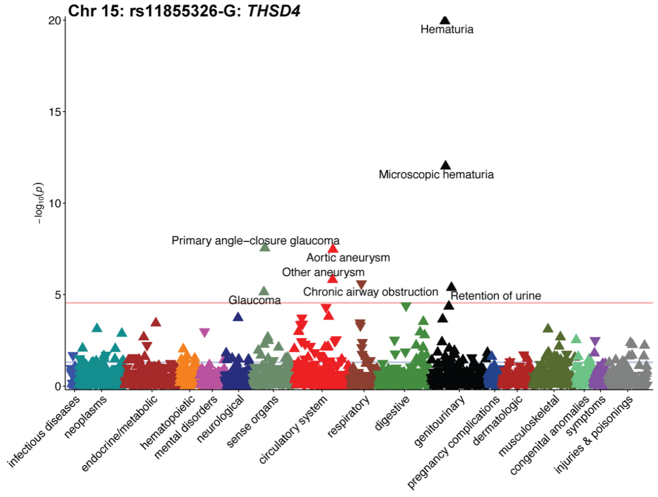

Chr 15: rs11855326-G: THSD4
Hematuria
Microscopic hematuria
Primary angle−closure glaucoma
Aortic aneurysm
Other aneurysm
Chronic airway obstruction
Glaucoma
Retention of urine
−log10(p)
0
5
10
15
20
infectious diseases
neoplasms
endocrine/metabolic
hematopoietic
mental disorders
neurological
sense organs
circulatory system
respiratory
digestive
genitourinary
pregnancy complications
dermatologic
musculoskeletal
congenital anomalies
symptoms
injuries & poisonings

**Chr 15: rs57515981-A: *FES***

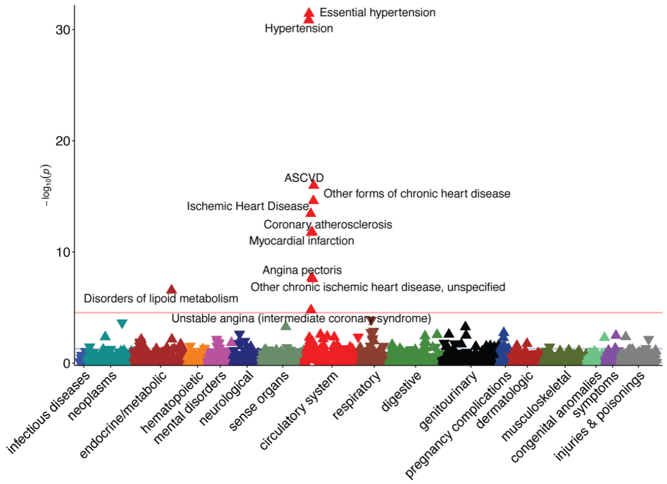


**Chr 16: rs4786481-T: *GLIS2***

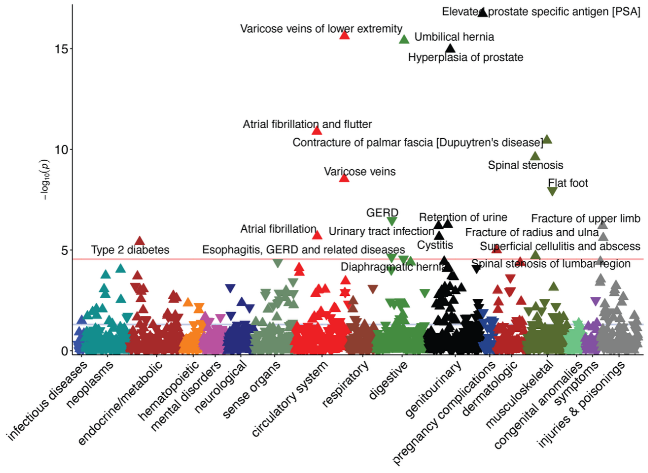


**Chr 16: rs12917707-T: *UMOD***

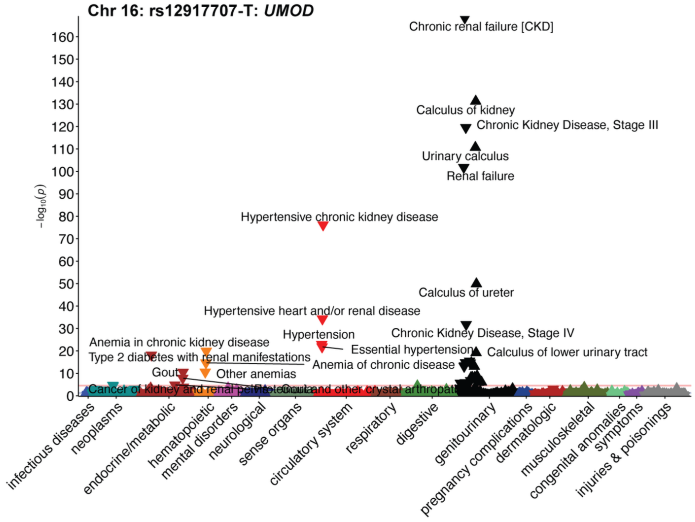

**Chr 16: rs1858800-C: *ZFHX3***

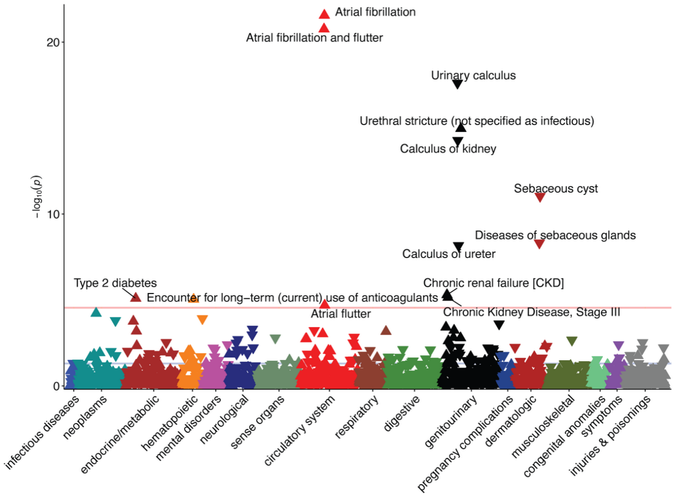


**Chr 17: rs78552540-A: *MAP2K4***

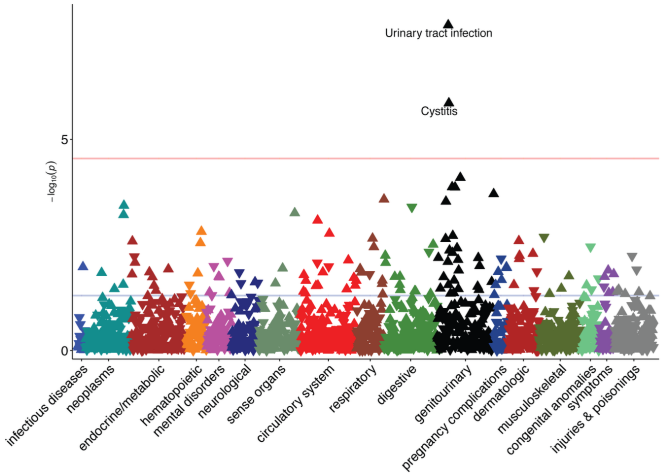


**Chr 18: NFATC1-A: *NFATC1***

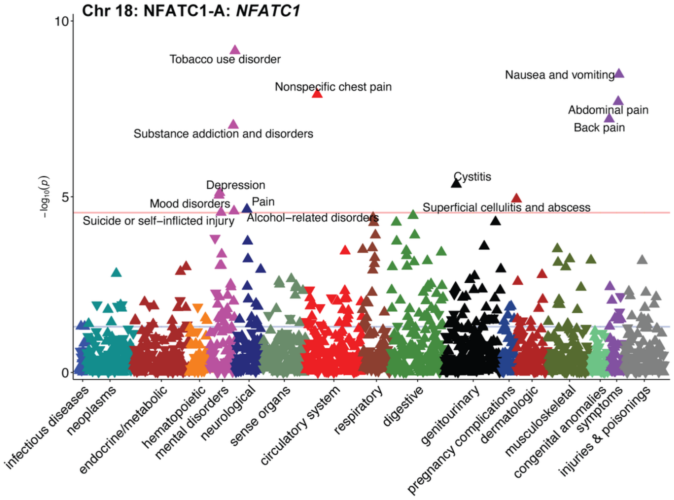

**Chr 2: rs34030812-C: *ARHGAP15***

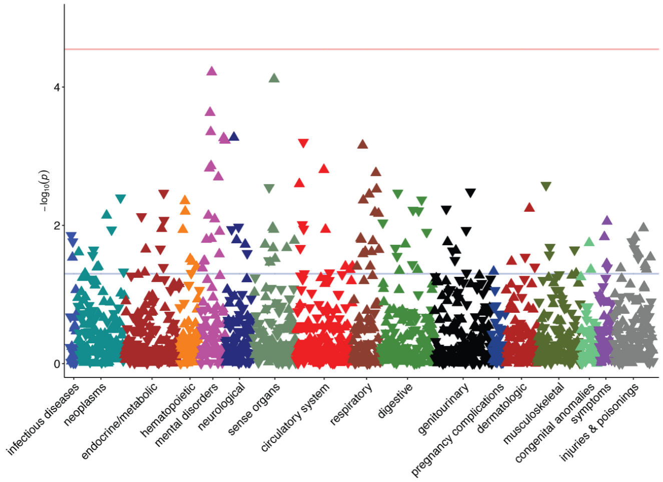


**Chr 10: rs471447-C: *CASP7***

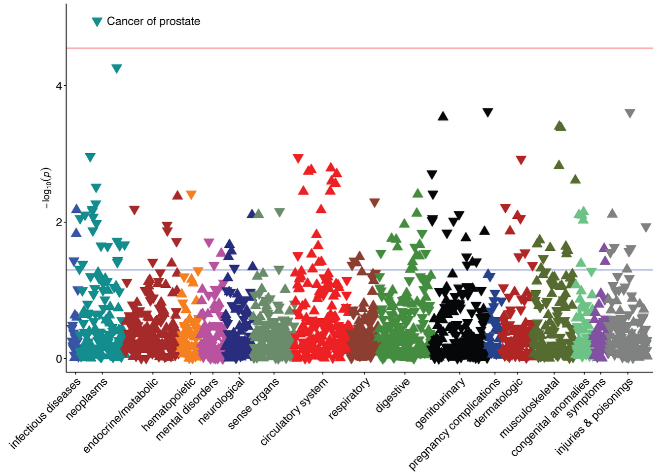


**Chr 13: rs9584326-G: *CLDN10***

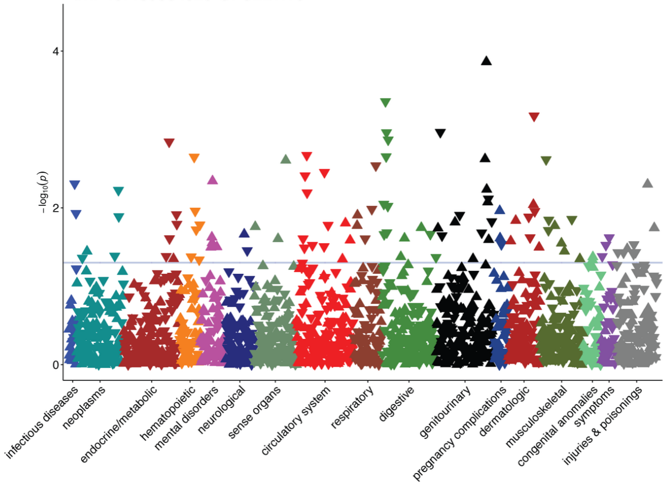

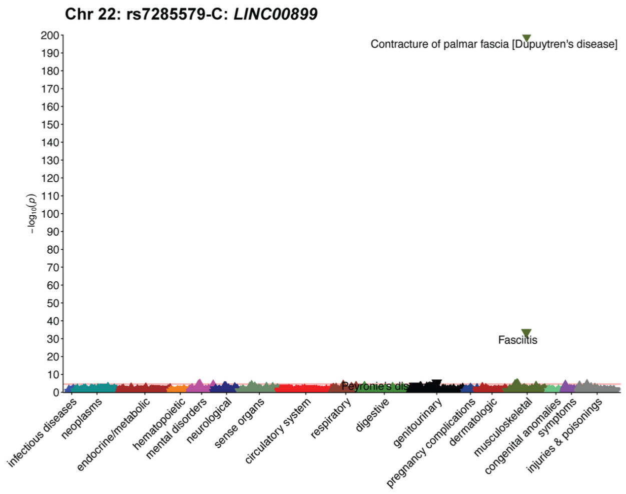

Chr 22: rs7285579-C: LINC00899
Contracture of palmar fascia [Dupuytren's disease]
Fasciitis
Peyronie's disease
$-\log_{10}(p)$
0
10
20
30
40
50
60
70
80
90
100
110
120
130
140
150
160
170
180
190
200
infectious diseases
neoplasms
endocrine/metabolic
hematopoietic
mental disorders
neurological
sense organs
circulatory system
respiratory
digestive
genitourinary
pregnancy complications
dermatologic
musculoskeletal
congenital anomalies
symptoms
injuries & poisonings


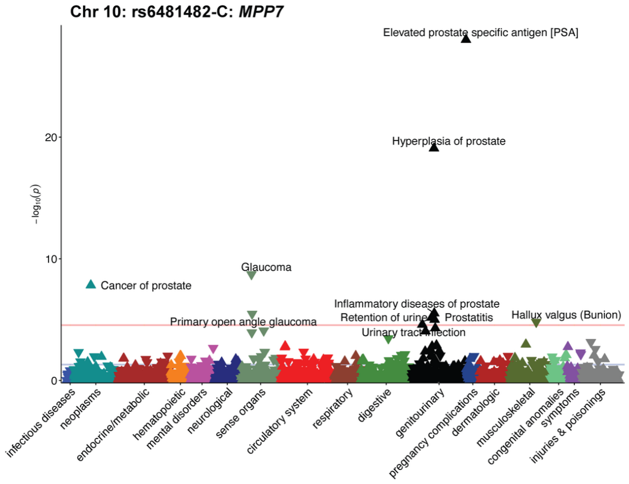

Chr 10: rs6481482-C: MPP7
Elevated prostate specific antigen [PSA]
Hyperplasia of prostate
Cancer of prostate
Glaucoma
Inflammatory diseases of prostate
Retention of urine
Prostatitis
Hallux valgus (Bunion)
Primary open angle glaucoma
Urinary tract infection
$-\log_{10}(p)$
0
10
20
infectious diseases
neoplasms
endocrine/metabolic
hematopoietic
mental disorders
neurological
sense organs
circulatory system
respiratory
digestive
genitourinary
pregnancy complications
dermatologic
musculoskeletal
congenital anomalies
symptoms
injuries & poisonings


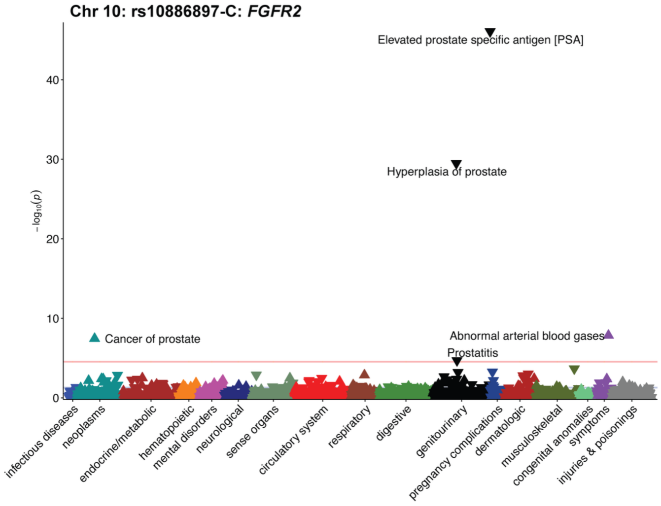

Chr 10: rs10886897-C: FGFR2
Elevated prostate specific antigen [PSA]
Hyperplasia of prostate
Cancer of prostate
Abnormal arterial blood gases
Prostatitis
$-\log_{10}(p)$
0
10
20
30
40
infectious diseases
neoplasms
endocrine/metabolic
hematopoietic
mental disorders
neurological
sense organs
circulatory system
respiratory
digestive
genitourinary
pregnancy complications
dermatologic
musculoskeletal
congenital anomalies
symptoms
injuries & poisonings

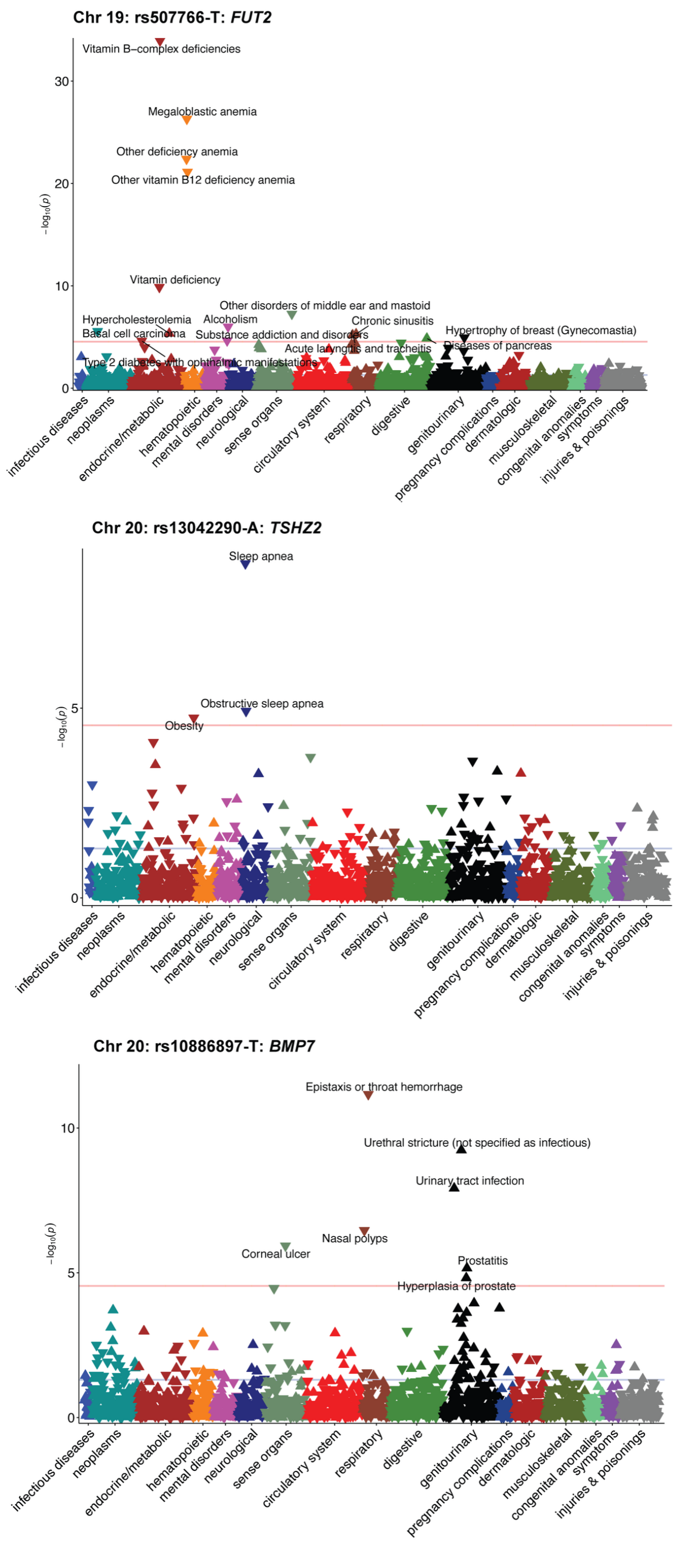
Chr 19: rs507766-T: FUT2
Vitamin B−complex deficiencies
Megaloblastic anemia
Other deficiency anemia
Other vitamin B12 deficiency anemia
Vitamin deficiency
Other disorders of middle ear and mastoid
Hypercholesterolemia
Alcoholism
Chronic sinusitis
Basal cell carcinoma
Substance addiction and disorders
Hypertrophy of breast (Gynecomastia)
Diseases of pancreas
Acute laryngitis and tracheitis
Type 2 diabetes with ophthalmic manifestations
Chr 20: rs13042290-A: TSHZ2
Sleep apnea
Obstructive sleep apnea
Obesity
Chr 20: rs10886897-T: BMP7
Epistaxis or throat hemorrhage
Urethral stricture (not specified as infectious)
Urinary tract infection
Nasal polyps
Corneal ulcer
Prostatitis
Hyperplasia of prostate
$-\log_{10}(p)$
infectious diseases
neoplasms
endocrine/metabolic
hematopoietic
mental disorders
neurological
sense organs
circulatory system
respiratory
digestive
genitourinary
pregnancy complications
dermatologic
musculoskeletal
congenital anomalies
symptoms
injuries & poisonings

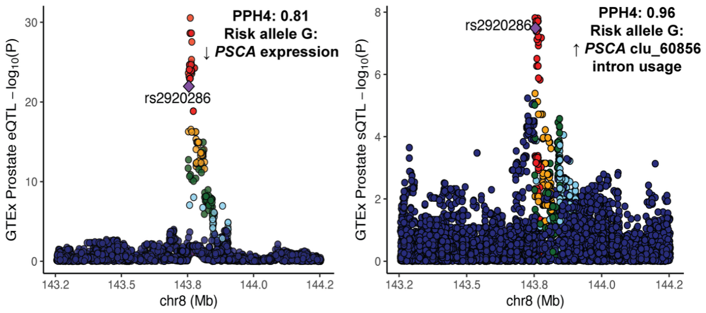


**Fig. S13. Colocalization of UTI GWAS chr8: rs2920286 locus** and prostate-specific *PSCA* eQTL and sQTL signals.

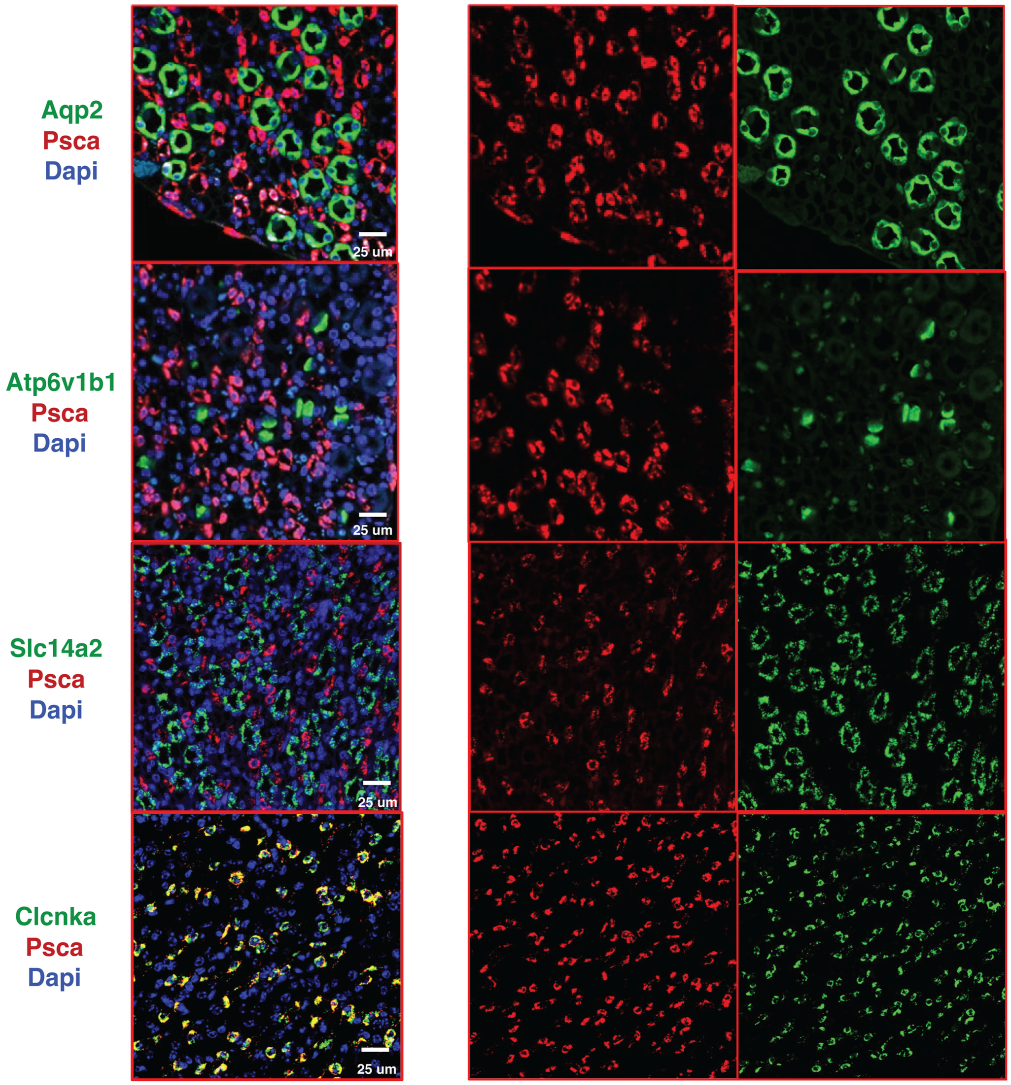


**Fig. S14. *Psca* RNA is exclusively expressed in the ascending thin limb of the kidney.** Imunofluorescence images of mouse kidney papilla co-stained with DAPI (blue) and *Psca* (red) alongside cell-type specific markers: Aqp2 (green, principal cells), Atp6v1b1 (green, intercalated cells), Slc14a2 (green, descending thin limb cells), and Clcnka (green, ascending thin limb cells). Left panels show merged images and right panels show individual channels. *Psca* expression colocalizes exclusively with Clcnka$^{+}$ ascending thin limb cells and is absent from Aqp2$^{+}$ principal cells, Atp6v1b1$^{+}$ intercalated cells, and Slc14a2$^{+}$ descending thin limb cells. Scale bars 25µm.

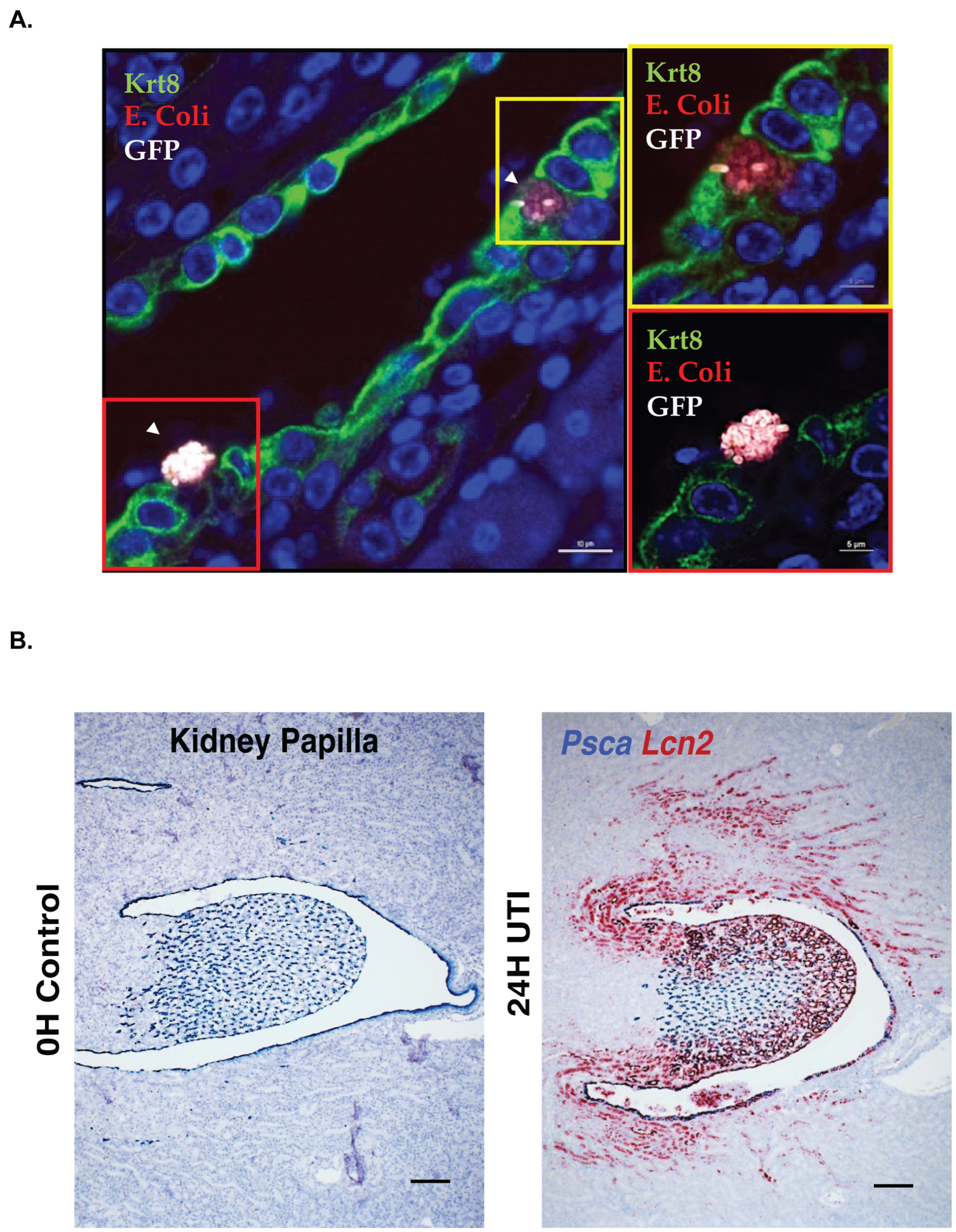


**Fig. S15.** ***E. coli*** **preferentially adheres to mouse kidney papillary epithelium and induces the kidney injury and antimicrobial marker Lcn2 in the papilla during UTI.** (A) Immunofluorescence of mouse kidney papilla showing preferential adhesion of GFP-expressing *E. coli* (red) to Krt8$^{+}$ papillary epithelial cells (green), with insets highlighting individual bacterial adhesion events at higher magnification. Scale bars shown. (B) RNAscope of mouse kidney papilla at 0 hours (control, left) and 24 hours post-transurethral *E. coli* inoculation (right), showing induction of *Lcn2* (red) in the papilla at 24 hours while *Psca* expression (blue) remains stable, demonstrating that *Psca* is constitutively expressed and does not overlap with the acute injury response marker *Lcn2*, which is restricted to collecting ducts. Scale bars = 200µM.

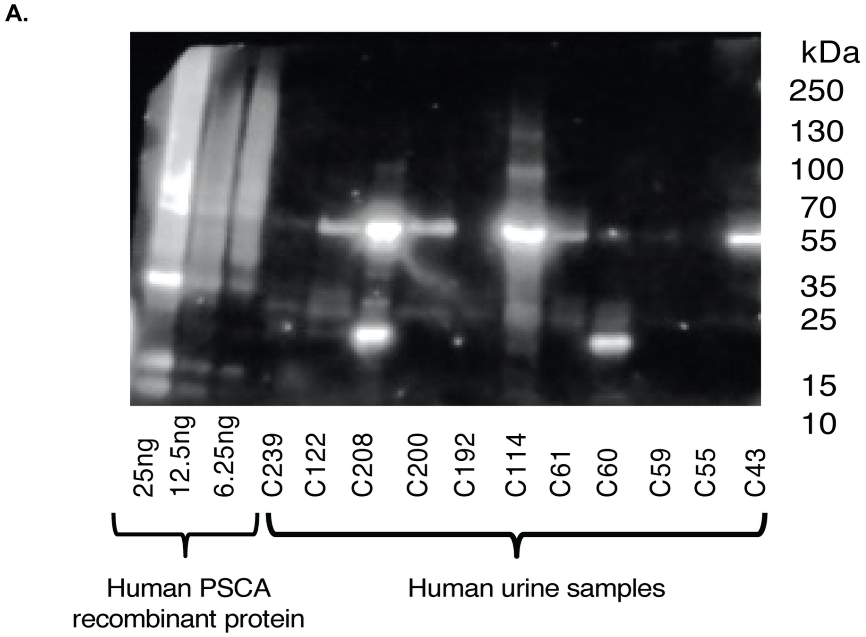


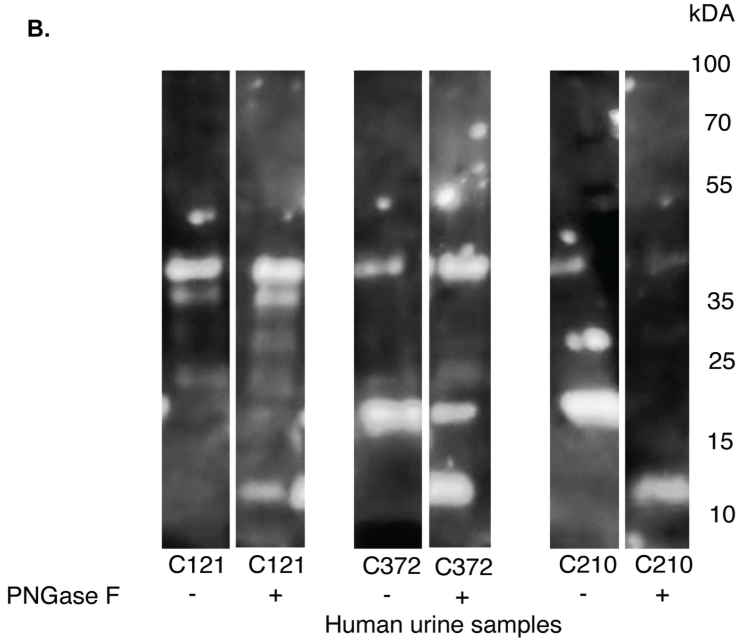


**Fig. S16. PSCA is detected in human urine samples and is heavily N-glycosylated.** (A) Western blot of human urinary PSCA across UTI-positive and UTI-negative urine samples alongside human recombinant PSCA protein standards. PSCA is detected as multiple higher molecular weight glycoforms (~20-55kDa) compared to the native ~12kDa protein, consistent with heavy glycosylation. (B) Western blot of human urine samples treated with PNGase F (+) or untreated (−), showing collapse of higher molecular weight PSCA glycoforms to lower molecular weight forms following enzymatic removal of N-linked glycans, confirming that urinary PSCA is heavily N-glycosylated.

**SUPPLEMENTARY NOTES:**

***UTI Case Inclusion and Exclusion Criteria.***

**1. UTI Inclusion**

Set of billing codes that indicate infectious or inflammatory conditions of the bladder, ureter, or kidneys caused by, or possibly caused by, a primary bacterial infection.

Specifications

(1) include codes that may be non-specific about site or organism (e.g. "urinary tract infection, unspecified")

(2) include codes related to UTI during pregnancy or postpartum

(3) do not include codes that specify viral, parasitic or fungal infection

(4) do not include codes that specify urethritis

(5) do not include infections that likely come from another organ system

   a. STI: e.g. gonococcal, chlamydial, syphilis
   b. Respiratory: e.g. TB, mycoplasma, actinomycosis
   c. GI: e.g. salmonella
   d. sepsis, systemic infection

(6) do not include codes that specify infection due to (or associated with) complication of devices, procedures, or other conditions

   a. catheter-associated UTI
   b. device-associated UTI
   c. UTI following abortion (spontaneous or induced)

(7) do not include conditions that only indicate only the "genitourinary tract" – *Non-specific; Low yield with many false positives*

(8) do not include inflammatory conditions almost always due to non-infectious processes (e.g. acute interstitial nephritis, chronic obstructive pyelonephritis, hemorrhagic cystitis) – *Low yield with many false positives*

**2. UTI Exclusion lists**

**A. Secondary UTI Exclusions**

1. Urethritis: Set of billing codes for infection of the urethra.

2. Secondary infection: Set of billing codes indicating that urinary tract infection may be secondary to an infection of a primary site, or secondary to fungal infection.
    a. STI: e.g. gonococcal, chlamydial, syphilis
    b. Respiratory: e.g. TB, mycoplasma, actinomycosis, diphtheria
    c. GI: e.g. salmonella
    d. Candiasis
3. Secondary to aborted pregnancy: Set of billing codes for UTI following miscarriages, abortions, and ectopic pregnancies.

**B. Acute Predisposing Factors Exclusions:**

1. Obstruction: Set of billing codes for potentially obstructive diseases in the urinary tract.
    (1) Urinary calculi
    (2) Chronic pyelonephritis
    (3) Foreign bodies in the urethra
    (4) Benign hypertrophy of prostate
2. Nosocomial Factor: Set of billing codes for nosocomial causes of infection, or nosocomial risk factors for UTI. This includes:
    (1) catheter or implanted device in the urinary tract,
    (2) cystotomy,
    (3) graft of urinary organ,
    (4) intra- or post-procedural complication of genitourinary system procedure

    This includes billing codes for complications of devices (e.g. "leakage of urinary catheter") because they indicate that the device was used.

**C. Chronic Predisposing Factors Exclusions:**

1. CAKUT: Set of billing codes for congenital abnormalities of the urinary tract and kidney, including VUR.
2. Immunocompromised: These are billing codes that indicate possible immunocompromised status.
3. Transplant: Billing codes that indicate the patient has received organ or tissue transplant. This includes codes for:
    (1) transplant status itself,
    (2) complications due to transplant,

(3) encounter for aftercare following transplant

4. Immunosuppressant therapy: Billing codes that indicate the patient is on, or was on, immunosuppressant therapy or chemotherapy. This includes codes for:
   (1) encounter for immunosuppressant therapy
   (2) personal history,
   (3) long-term (current) use
5. Immunodeficiency disorders: Billing codes that indicate the patient has an immunodeficiency disorder.
6. HIV: Billing codes for HIV. This includes asymptomatic HIV status.
7. Cancer: Billing codes for malignant neoplasms. Benign neoplasms are not included.
8. Splenic disorders: Billing codes for diseases of the spleen.

### 3. UTI Case Types

An individual can be assigned to one of three groups:

(1) **Case**: An individual who has had at least two instances of bacterial UTI which is the result of a community-acquired (as opposed to nosocomial) and primary process (i.e. not secondary to another disease, condition, or procedure).

(2) **Control**: An individual who has never had any kind of UTI, primary or secondary.

(3) **Exclude**: Individuals who only has a single instance of UTI as a primary process or whose status as case or control is not clear; they are excluded from further study.

***Descriptions of Biobank Cohorts***

**Electronic Medical Records and Genomics-III (eMERGE-III)** (*37*)**:** The eMERGE network consortium consists of 12 medical centers with electronic health records (EHRs) linked to genome-wide genotype data for 102,138 individuals. The genotyping and imputation of the eMERGE cohort have been recently described in detail (*38*). Briefly, we implemented the mimimac3 missing variant imputation model with genome-wide imputation using the HRC1.1 reference (Michigan Imputation Server) in genome build 37 (hg19) for each genotyping platform in a separate batch. After imputation, we merged all the 81 imputed batches based on position using bcftools (http://researchcomputing.syr.edu/bcftools/). The quality control filters required a marker to have $R^2 \geq 0.8$ in ≥75% of 81 imputation batches and Minor Allele Frequency MAF ≥0.01. We applied a principal component analysis (PCA) using FlashPCA (*39*) using 35,226 common (MAF >1%) and independent variants after LD pruning with PLINK (--indep-pairwise 500 50 0.05). We used KING to identify cryptically related subjects and removed one individual per related pair with second-degree or higher relatedness (*40*). We removed ancestry outliers, re-ran PCA, and adjusted GWAS analysis for significant principal components to reduce any potential bias from population stratification. We defined genetic ancestry using principal component analysis and a random forest algorithm trained on the 1000 Genomes Project. The eMERGE cohort included 7,631 cases and 42,265 controls of European ancestry, 1,152 cases and 8,525 controls of African ancestry, and 311 cases and 1,399 controls of East Asian ancestry.

**UK Biobank (UKBB)** (*41*)**:** UKBB is a large prospective cohort based in the United Kingdom that enrolled individuals aged 40-69 for the purpose of genetic studies (*42*). This cohort comprised 488,377 individuals recruited since 2006, genotyped with high-density SNP arrays, and linked to electronic health record data. All individuals underwent genome-wide genotyping with UK Biobank Axiom array from Affymetrix and UK BiLEVE Axiom arrays (~825,000 markers). Genotype imputation was carried out using a 1000 Genomes reference panel with IMPUTE4 software (*41*, *43*, *44*). We then applied QC filters similar to eMERGE-III, retaining 9,233,643 common (MAF≥0.01) variants imputed with high confidence ($R^2 \geq 0.8$). For principal component analysis by FlashPCA (*39*) we selected common variants (MAF >1%) after LD pruning with PLINK (--indep-pairwise 500 50 0.05). We removed ancestry outliers, re-ran PCA, and adjusted

GWAS analysis for significant principal components. Similar to eMERGE, we defined genetic ancestry using principal component analysis and a random forest algorithm trained on the 1000 Genomes Project reference populations. The final UK Biobank UTI cohort included 16,326 cases and 424,272 controls of European ancestry.

**All-of-Us (AoU)** (*45*)**:** The All of Us Research Program has been described in detail previously (*45*). We used the second data release, which included 312,944 participants with available genotype data. All analyses were conducted using the All of Us Researcher Workbench within the Google Cloud environment. The AoU genotype array data were imputed using our previously described imputation pipeline and quality control procedures (*46*). Briefly, participants were genotyped using the Illumina Global Diversity Array (GDA). Prior to imputation, variants with minor allele frequency (MAF) $\leq 0.005$ or genotype missingness $\geq 5\%$ were excluded, and genomic coordinates were lifted over from GRCh38 to hg19 where applicable. We subsequently applied the TOPMed pre-imputation quality control pipeline to harmonize allele designations and remove poorly mapping variants. Genotypes were pre-phased using EAGLE v2 and imputed with Minimac4 using the 1000 Genomes Project Phase 3 v5 reference panel. The resulting imputed genotype data were used for all downstream genetic analyses in this study. Standard genetic quality control procedures were applied before conducting association analyses. Similar to eMERGE, we defined genetic ancestry using principal component analysis and a random forest algorithm trained on the 1000 Genomes Project reference populations. The final AoU cohort included 10,388 cases and 96,597 controls of European ancestry, 3,844 cases and 40,013 controls of African ancestry, 4,286 cases and 36,528 controls of Admixed American ancestry, 287 cases and 6,116 controls of East Asian ancestry, and 112 cases and 2,359 controls of South Asian ancestry.

**BioVU** (*47*) **Vanderbilt University Medical Center Biobank (VUMC):** BioVU is a large hospital-based biobank at Vanderbilt University Medical Center (VUMC) that links DNA samples collected during clinical care with de-identified electronic health records to facilitate genomic and translational research. Genotyping was performed by the Vanderbilt Technologies for Advanced Genomics (VANTAGE) using the Illumina MEGAEX platform. Genotype quality control was performed using PLINK v1.9, and ancestry principal components were calculated using common variants (MAF >1%) with high call rates (>98%), after excluding variants that deviated from

Hardy–Weinberg equilibrium (HWE, $P < 1 \times 10^{-6}$) or were in linkage disequilibrium. We restricted analyses to individuals of European ancestry based on principal components and HapMap reference populations. Genotypes were imputed using the Michigan Imputation Server with the Haplotype Reference Consortium (HRC) v1.1 reference panel on the GRCh37/hg19 genome build. We used SAIGE (*48*) to test associations between genetic variants and UTI risk under an additive genetic model, adjusting for sex, year of first clinical visit, EHR length, and ancestry PCs to account for residual population structure (*49*). Post-GWAS quality control was performed using EASYQC (*50*) to exclude poorly imputed variants ($R^2$ <0.8), variants with MAF <0.01, and variants deviating from HWE ($P < 1 \times 10^{-6}$). The final VUMC cohort included 5,785 UTI cases and 30,880 controls of European ancestry.

**Other Datasets:** We additionally used published GWAS summary statistics from the VA Million Veteran Program (*51*), FinnGen (*52*), and 23andMe (*53*). The Million Veteran Program (MVP) is a large national biobank established by the U.S. Department of Veterans Affairs that integrates genetic data with electronic health records and survey information from U.S. veterans for genomic research. For our study, we utilized publicly available GWAS summary statistics for UTI PheCode 591 available from MVP (*51*). MVP genotype data were imputed using the TOPMed reference panel, and genetic ancestry was inferred using principal component analysis and a random forest classifier trained on reference populations from the 1000 Genomes Project. We analyzed UTI cases and controls across three ancestry groups: European (46,012 cases and 368,445 controls), African (15,423 cases and 93,003 controls), and Admixed American (5,852 cases and 54,759 controls). FinnGen is a large prospective biobank study in Finland that integrates genomic data with nationwide health registry information to investigate the genetic basis of disease. We used publicly available GWAS summary statistics released by the FinnGen consortium (*52*). We analyzed FinnGen phenotype of cystitis (N14), which included 61,460 cases and 403,328 controls. FinnGen genotype data were generated using genotyping arrays and imputed using a Finnish population-specific reference panel of 3,775 high-coverage whole-genome sequences. For our meta-analysis, we restricted variants to those with high imputation quality (INFO >0.8) and MAF >0.01. The FinnGen GWAS summary statistics were generated using the FinnGen GWAS pipeline, which accounts for population structure and relatedness. Lastly, 23andMe is a large direct-to-consumer genetic testing and research platform that has enabled genetic studies across millions of consented research participants. We used GWAS summary statistics from 23andMe generated by testing genotype data against customer survey data on the frequency of urinary tract infections (UTIs) (*53*). Genotyping was performed using the V1 and V2 23andMe platforms, variants of the Illumina HumanHap550 BeadChip. Genotypes were then imputed using the 1000 Genomes Project reference panel. We used pre-computed GWAS summary statistics from a total of 35,000 UTI cases and 33,478 controls.

**Web Resources**

eMERGE: https://emerge.mc.vanderbilt.edu/

VCFtools: http://vcftools.sourceforge.net/index.html

Bcftools: https://samtools.github.io/bcftools/bcftools.html

PLINK: https://www.cog-genomics.org/plink2

PYTHON: https://www.python.org/

R: https://www.r-project.org/

KING: http://people.virginia.edu/~wc9c/KING/

FlashPCA: https://github.com/gabraham/flashpca

Michigan Imputation Server: https://imputationserver.sph.umich.edu/index.html

Human Reference Consortium: http://www.haplotype-reference-consortium.org/site

PheWAS: https://phewascatalog.org/

43. A. Auton, G. R. Abecasis, D. M. Altshuler, R. M. Durbin, D. R. Bentley, A. Chakravarti, A. G. Clark, P. Donnelly, E. E. Eichler, P. Flicek, S. B. Gabriel, R. A. Gibbs, E. D. Green, M. E. Hurles, B. M. Knoppers, J. O. Korbel, E. S. Lander, C. Lee, H. Lehrach, E. R.

Mardis, G. T. Marth, G. A. McVean, D. A. Nickerson, J. P. Schmidt, S. T. Sherry, J. Wang, R. K. Wilson, E. Boerwinkle, H. Doddapaneni, Y. Han, V. Korchina, C. Kovar, S. Lee, D. Muzny, J. G. Reid, Y. Zhu, Y. Chang, Q. Feng, X. Fang, X. Guo, M. Jian, H. Jiang, X. Jin, T. Lan, G. Li, J. Li, Y. Li, S. Liu, X. Liu, Y. Lu, X. Ma, M. Tang, B. Wang, G. Wang, H. Wu, R. Wu, X. Xu, Y. Yin, D. Zhang, W. Zhang, J. Zhao, M. Zhao, X. Zheng, N. Gupta, N. Gharani, L. H. Toji, N. P. Gerry, A. M. Resch, J. Barker, L. Clarke, L. Gil, S. E. Hunt, G. Kelman, E. Kulesha, R. Leinonen, W. M. McLaren, R. Radhakrishnan, A. Roa, D. Smirnov, R. E. Smith, I. Streeter, A. Thormann, I. Toneva, B. Vaughan, X. Zheng-Bradley, R. Grocock, S. Humphray, T. James, Z. Kingsbury, R. Sudbrak, M. W. Albrecht, V. S. Amstislavskiy, T. A. Borodina, M. Lienhard, F. Mertes, M. Sultan, B. Timmermann, M. L. Yaspo, L. Fulton, V. Ananiev, Z. Belaia, D. Beloslyudtsev, N. Bouk, C. Chen, D. Church, R. Cohen, C. Cook, J. Garner, T. Hefferon, M. Kimelman, C. Liu, J. Lopez, P. Meric, C. O'Sullivan, Y. Ostapchuk, L. Phan, S. Ponomarov, V. Schneider, E. Shekhtman, K. Sirotkin, D. Slotta, H. Zhang, S. Balasubramaniam, J. Burton, P. Danecek, T. M. Keane, A. Kolb-Kokocinski, S. McCarthy, J. Stalker, M. Quail, C. J. Davies, J. Gollub, T. Webster, B. Wong, Y. Zhan, C. L. Campbell, Y. Kong, A. Marcketta, F. Yu, L. Antunes, M. Bainbridge, A. Sabo, Z. Huang, L. J. M. Coin, L. Fang, Q. Li, Z. Li, H. Lin, B. Liu, R. Luo, H. Shao, Y. Xie, C. Ye, C. Yu, F. Zhang, H. Zheng, H. Zhu, C. Alkan, E. Dal, F. Kahveci, E. P. Garrison, D. Kural, W. P. Lee, W. F. Leong, M. Stromberg, A. N. Ward, J. Wu, M. Zhang, M. J. Daly, M. A. DePristo, R. E. Handsaker, E. Banks, G. Bhatia, G. Del Angel, G. Genovese, H. Li, S. Kashin, S. A. McCarroll, J. C. Nemesh, R. E. Poplin, S. C. Yoon, J. Lihm, V. Makarov, S. Gottipati, A. Keinan, J. L. Rodriguez-Flores, T. Rausch, M. H. Fritz, A. M. Stütz, K. Beal, A. Datta, J. Herrero, G. R. S. Ritchie, D. Zerbino, P. C. Sabeti, I. Shlyakhter, S. F. Schaffner, J. Vitti, D. N. Cooper, E. V. Ball, P. D. Stenson, B. Barnes, M. Bauer, R. K. Cheetham, A. Cox, M. Eberle, S. Kahn, L. Murray, J. Peden, R. Shaw, E. E. Kenny, M. A. Batzer, M. K. Konkel, J. A. Walker, D. G. MacArthur, M. Lek, R. Herwig, L. Ding, D. C. Koboldt, D. Larson, K. Ye, S. Gravel, A. Swaroop, E. Chew, T. Lappalainen, Y. Erlich, M. Gymrek, T. F. Willems, J. T. Simpson, M. D. Shriver, J. A. Rosenfeld, C. D. Bustamante, S. B. Montgomery, F. M. De La Vega, J. K. Byrnes, A. W. Carroll, M. K. DeGorter, P. Lacroute, B. K. Maples, A. R. Martin, A. Moreno-Estrada, S. S.

Shringarpure, F. Zakharia, E. Halperin, Y. Baran, E. Cerveira, J. Hwang, A. Malhotra, D. Plewczynski, K. Radew, M. Romanovitch, C. Zhang, F. C. L. Hyland, D. W. Craig, A. Christoforides, N. Homer, T. Izatt, A. A. Kurdoglu, S. A. Sinari, K. Squire, C. Xiao, J. Sebat, D. Antaki, M. Gujral, A. Noor, K. Ye, E. G. Burchard, R. D. Hernandez, C. R. Gignoux, D. Haussler, S. J. Katzman, W. J. Kent, B. Howie, A. Ruiz-Linares, E. T. Dermitzakis, S. E. Devine, H. M. Kang, J. M. Kidd, T. Blackwell, S. Caron, W. Chen, S. Emery, L. Fritsche, C. Fuchsberger, G. Jun, B. Li, R. Lyons, C. Scheller, C. Sidore, S. Song, E. Sliwerska, D. Taliun, A. Tan, R. Welch, M. K. Wing, X. Zhan, P. Awadalla, A. Hodgkinson, Y. Li, X. Shi, A. Quitadamo, G. Lunter, J. L. Marchini, S. Myers, C. Churchhouse, O. Delaneau, A. Gupta-Hinch, W. Kretzschmar, Z. Iqbal, I. Mathieson, A. Menelaou, A. Rimmer, D. K. Xifara, T. K. Oleksyk, Y. Fu, X. Liu, M. Xiong, L. Jorde, D. Witherspoon, J. Xing, B. L. Browning, S. R. Browning, F. Hormozdiari, P. H. Sudmant, E. Khurana, C. Tyler-Smith, C. A. Albers, Q. Ayub, Y. Chen, V. Colonna, L. Jostins, K. Walter, Y. Xue, M. B. Gerstein, A. Abyzov, S. Balasubramanian, J. Chen, D. Clarke, Y. Fu, A. O. Harmanci, M. Jin, D. Lee, J. Liu, X. J. Mu, J. Zhang, Y. Zhang, C. Hartl, K. Shakir, J. Degenhardt, S. Meiers, B. Raeder, F. P. Casale, O. Stegle, E. W. Lameijer, I. Hall, V. Bafna, J. Michaelson, E. J. Gardner, R. E. Mills, G. Dayama, K. Chen, X. Fan, Z. Chong, T. Chen, M. J. Chaisson, J. Huddleston, M. Malig, B. J. Nelson, N. F. Parrish, B. Blackburne, S. J. Lindsay, Z. Ning, Y. Zhang, H. Lam, C. Sisu, D. Challis, U. S. Evani, J. Lu, U. Nagaswamy, J. Yu, W. Li, L. Habegger, H. Yu, F. Cunningham, I. Dunham, K. Lage, J. B. Jespersen, H. Horn, D. Kim, R. Desalle, A. Narechania, M. A. W. Sayres, F. L. Mendez, G. D. Poznik, P. A. Underhill, D. Mittelman, R. Banerjee, M. Cerezo, T. W. Fitzgerald, S. Louzada, A. Massaia, F. Yang, D. Kalra, W. Hale, X. Dan, K. C. Barnes, C. Beiswanger, H. Cai, H. Cao, B. Henn, D. Jones, J. S. Kaye, A. Kent, A. Kerasidou, R. Mathias, P. N. Ossorio, M. Parker, C. N. Rotimi, C. D. Royal, K. Sandoval, Y. Su, Z. Tian, S. Tishkoff, M. Via, Y. Wang, H. Yang, L. Yang, J. Zhu, W. Bodmer, G. Bedoya, Z. Cai, Y. Gao, J. Chu, L. Peltonen, A. Garcia-Montero, A. Orfao, J. Dutil, J. C. Martinez-Cruzado, R. A. Mathias, A. Hennis, H. Watson, C. McKenzie, F. Qadri, R. LaRocque, X. Deng, D. Asogun, O. Folarin, C. Happi, O. Omoniwa, M. Stremlau, R. Tariyal, M. Jallow, F. S. Joof, T. Corrah, K. Rockett, D. Kwiatkowski, J. Kooner, T. T. Hien, S. J. Dunstan, N. ThuyHang, R. Fonnie, R. Garry, L. Kanneh, L. Moses, J. Schieffelin, D. S. Grant, C.

Gallo, G. Poletti, D. Saleheen, A. Rasheed, L. D. Brooks, A. L. Felsenfeld, J. E. McEwen, Y. Vaydylevich, A. Duncanson, M. Dunn, J. A. Schloss, A global reference for human genetic variation. *Nat. 2015 5267571* **526**, 68–74 (2015).

51. A. Verma, J. E. Huffman, A. Rodriguez, M. Conery, M. Liu, Y. L. Ho, Y. Kim, D. A. Heise, L. Guare, V. A. Panickan, H. Garcon, F. Linares, L. Costa, I. Goethert, R. Tipton, J. Honerlaw, L. Davies, S. Whitbourne, J. Cohen, D. C. Posner, R. Sangar, M. Murray, X. Wang, D. R. Dochtermann, P. Devineni, Y. Shi, T. N. Nandi, T. L. Assimes, C. A. Brunette, R. J. Carroll, R. Clifford, S. Duvall, J. Gelernter, A. Hung, S. K. Iyengar, J. Joseph, R. Kember, H. Kranzler, C. M. Kripke, D. Levey, S. W. Luoh, V. C. Merritt, C. Overstreet, J. D. Deak, S. F. A. Grant, R. Polimanti, P. Roussos, G. Shakt, Y. V. Sun, N.

Tsao, S. Venkatesh, G. Voloudakis, A. Justice, E. Begoli, R. Ramoni, G. Tourassi, S. Pyarajan, P. Tsao, C. J. O'Donnell, S. Muralidhar, J. Moser, J. P. Casas, A. G. Bick, W. Zhou, T. Cai, B. F. Voight, K. Cho, J. M. Gaziano, R. K. Madduri, S. Damrauer, K. P. Liao, Diversity and scale: Genetic architecture of 2068 traits in the VA Million Veteran Program. *Science* **385**, eadj1182 (2024).

52. M. I. Kurki, J. Karjalainen, P. Palta, T. P. Sipilä, K. Kristiansson, K. M. Donner, M. P. Reeve, H. Laivuori, M. Aavikko, M. A. Kaunisto, A. Loukola, E. Lahtela, H. Mattsson, P. Laiho, P. Della Briotta Parolo, A. A. Lehisto, M. Kanai, N. Mars, J. Rämö, T. Kiiskinen, H. O. Heyne, K. Veerapen, S. Rüeger, S. Lemmelä, W. Zhou, S. Ruotsalainen, K. Pärn, T. Hiekkalinna, S. Koskelainen, T. Paajanen, V. Llorens, J. Gracia-Tabuenca, H. Siirtola, K. Reis, A. G. Elnahas, B. Sun, C. N. Foley, K. Aalto-Setälä, K. Alasoo, M. Arvas, K. Auro, S. Biswas, A. Bizaki-Vallaskangas, O. Carpen, C. Y. Chen, O. A. Dada, Z. Ding, M. G. Ehm, K. Eklund, M. Färkkilä, H. Finucane, A. Ganna, A. Ghazal, R. R. Graham, E. M. Green, A. Hakanen, M. Hautalahti, Å. K. Hedman, M. Hiltunen, R. Hinttala, I. Hovatta, X. Hu, A. Huertas-Vazquez, L. Huilaja, J. Hunkapiller, H. Jacob, J. N. Jensen, H. Joensuu, S. John, V. Julkunen, M. Jung, J. Junttila, K. Kaarniranta, M. Kähönen, R. Kajanne, L. Kallio, R. Kälviäinen, J. Kaprio, N. Kerimov, J. Kettunen, E. Kilpeläinen, T. Kilpi, K. Klinger, V. M. Kosma, T. Kuopio, V. Kurra, T. Laisk, J. Laukkanen, N. Lawless, A. Liu, S. Longerich, R. Mägi, J. Mäkelä, A. Mäkitie, A. Malarstig, A. Mannermaa, J. Maranville, A. Matakidou, T. Meretoja, S. V. Mozaffari, M. E. K. Niemi, M. Niemi, T. Niiranen, C. J. O'Donnell, M. Obeidat, G. Okafo, H. M. Ollila, A. Palomäki, T. Palotie, J. Partanen, D. S. Paul, M. Pelkonen, R. K. Pendergrass, S. Petrovski, A. Pitkäranta, A. Platt, D. Pulford, E. Punkka, P. Pussinen, N. Raghavan, F. Rahimov, D. Rajpal, N. A. Renaud, B. Riley-Gillis, R. Rodosthenous, E. Saarentaus, A. Salminen, E. Salminen, V. Salomaa, J. Schleutker, R. Serpi, H. yi Shen, R. Siegel, K. Silander, S. Siltanen, S. Soini, H. Soininen, J. H. Sul, I. Tachmazidou, K. Tasanen, P. Tienari, S. Toppila-Salmi, T. Tukiainen, T. Tuomi, J. A. Turunen, J. C. Ulirsch, F. Vaura, P. Virolainen, J. Waring, D. Waterworth, R. Yang, M. Nelis, A. Reigo, A. Metspalu, L. Milani, T. Esko, C. Fox, A. S. Havulinna, M. Perola, S. Ripatti, A. Jalanko, T. Laitinen, T. P. Mäkelä, R. Plenge, M. McCarthy, H. Runz, M. J. Daly, A. Palotie, FinnGen provides genetic insights from a well-phenotyped isolated population. *Nat. 2023 6137944* **613**, 508–518 (2023).

53. C. Tian, B. S. Hromatka, A. K. Kiefer, N. Eriksson, S. M. Noble, J. Y. Tung, D. A. Hinds, Genome-wide association and HLA region fine-mapping studies identify susceptibility loci for multiple common infections. *Nat. Commun.* **8**, 599 (2017).
